# Valuation system connectivity is correlated with poly-drug use in young adults

**DOI:** 10.1101/2020.11.17.20233601

**Authors:** Kavinash Loganathan, Jinglei Lv, Vanessa Cropley, Andrew Zalesky, Eric Tatt Wei Ho

## Abstract

Poly-drug consumption contributes to fatal overdose in more than half of all poly-drug users. Analyzing decision-making networks may give insight into the motivations behind poly-drug use. We correlated average functional connectivity of the valuation system (VS), executive control system (ECS) and valuation-control complex (VCC) in a large population sample (n=992) with drug use behaviour. VS connectivity is correlated with sedative use, ECS connectivity is separately correlated with hallucinogens and opiates. Network connectivity is also correlated with drug use via two-way interactions with other substances including alcohol and tobacco. These preliminary findings can contribute to our understanding of the common combinations of substance co-use and associated neural patterns.

## Main Text

The pursuit of a desired internal state is known to drive drug-seeking and -taking behaviour (Hellberg et al., 2019). For example, an individual in pain may consume sedating substances such as marijuana or sedatives for relief (Choi et al., 2017; Shurman et al., 2010), while individuals looking to experience enhanced levels of euphoria may choose stimulants or cocaine (Breiter et al., 1997; Klega and Keehbauch, 2018). Poly-drug users mix-and-match different combinations in an attempt to increase the efficacy (Scott et al., 2007) or attenuate drawbacks (Fisk et al., 2011) of a particular substance. This tendency to co-use multiple drugs can cause a fatal overdose (Su et al., 2019). Many rehabilitating patients having a history of co-using more than one drug (Su et al., 2019; Wang et al., 2018) thereby requiring different treatment strategies (Hiebler-Ragger and Unterrainer, 2019; Weigl et al., 2017) due to higher levels of risky behaviour and psychopathology, decreased cognitive functioning as well as poorer treatment outcome and engagement (Wang et al., 2017).

Addictive drugs are thought to disrupt the brains’ reward system (Koob and Le Moal, 2008). These substances are consumed to achieve desired internal states (e.g. euphoria or pain relief), with regions of the Valuation and Executive Control System responsible for weighing drug value in relation to these states (Loganathan and Ho, 2021). Given that valuation of drugs changes based on internal and external factors as well as the increased complexity of the addiction phenotype among polyu-drug users, this study will focus on the interaction between Valuation and Executive Control System connectivity and poly-drug use in young adults. Value, the currency of choice, fluctuates based on internal states (e.g. satiety, deprivation) as well as external circumstances (e.g. socioeconomic factors, health status) (Compton et al., 2007; Davis and Johnson, 2008; Helie et al., 2017). The process of assigning value to different options may be regulated by the valuation and executive control systems of the brain (van den Bos and McClure, 2013). The valuation system (VS, comprised of the ventromedial prefrontal cortex, posterior cingulate cortex, amygdala and nucleus accumbens) may be involved in estimating the incentive value of different options (van den Bos and McClure, 2013) and is hypothesized to interface between pleasure and pain (Kringelbach, 2005). The executive control system (ECS, consisting of the lateral prefrontal cortex, dorsal anterior cingulate cortex and posterior parietal cortex), guides behaviour in pursuit of long-term goals (van den Bos and McClure, 2013). The ECS may assist in refining VS-encoded values by incorporating elements of future-thinking and careful deliberation (van den Bos and McClure, 2013). Addiction is thought to be associated with an imbalance in valuation, as drug-related stimuli are attributed greater salience compared to non-drug stimuli (Helie et al., 2017). The VS and ECS are linked with substance use behaviour, both as a network (Zhai et al., 2015) and as individual regions (Bedi et al., 2015). Normally, the interaction between VS and ECS is balanced, modulating reward processing by assigning values to various reward and non-reward options to facilitate optimal decision-making (Dalley et al., 2011; Xie et al., 2014; Zhai et al., 2015). During drug dependence the valuation system activation appears not to be counteracted by the control system which has since surrendered its regulatory abilities, tilting this once-mutually-reinforcing system towards the instrumental pursuit of drugs (Xie et al., 2014; Zhai et al., 2015).

Addictive drugs operate via different pathways in the brain (Leurquin-Sterk et al., 2018; Solinas et al., 2019). Taken together with the tendency to exclude poly-drug users from individual substance use neuroimaging studies (Zou et al., 2015), these limitations have restricted the development of a unifying concept underpinning drug use, further complicating efforts to develop focused treatment plans targeted at poly-drug users (for a review, see Wang *et al* (2017)). Given that poly-drug use contributes to more than 50% of fatal overdoses (Coffin et al., 2003), interrogating the interactions between different drugs is a necessary first step in unravelling the complex psychopathology underpinning drug addiction and what is required to treat it. This study aims to provide preliminary evidence linking functional connectivity of the VS and ECS with poly-substance use to infer the common combinations of substance co-use and associated neural patterns. This is done by correlating the connectivity of the VS, ECS and the valuation-control complex (VCC, a hybrid network comprised of VS and ECS regions) with poly-drug use in the Human Connectome Project Healthy Young Adult dataset using multivariate generalized linear models. We hypothesize that functional connectivity within the VS, ECS and VCC would all be correlated with alcohol, tobacco, opiates, cocaine, stimulants, sedatives and marijuana use. We further hypothesize that in some cases, connectivity may be correlated with substance use via a paired interaction with another drug (eg VS connectivity paired with sedative use may be correlated with cocaine use).

Functional MRI data from the Human Connectome Project (HCP, Smith *et al*., 2013) was sourced for healthy adults of both genders (n = 992, 463 males, 529 females; age range = 22-37). Recruitment procedures, inclusion/exclusion criteria and pre-processing steps are described elsewhere (Glasser et al., 2013; Smith et al., 2013a). Additional steps such as global signal regression are deemed too aggressive for use with HCP imaging data and are not implemented as it has been known to introduce artifactual changes in correlations patterns, fundamentally altering interregional correlations. These changes are thought to be dependent on the underlying true interregional correlation pattern (Anderson et al., 2011; Glasser et al., 2013; Saad et al., 2012; Smith et al., 2013b). The functional scans used in this study have already been cleaned of structured noise via joint use of independent component analysis (ICA) and FMRIB’s ICA-based X-noisifier (FIX) to remove non-neural spatio-temporal components from each highpass-filtered fMRI scan. Initial head motion correction is performed by registering fMRI data to a distorted gradient echo EPI single-band reference image. For each subject, rigid body head motion was estimated using FSL’s MCFLIRT routine to derive a motion transformation matrix for each time point. Each transform is described by 6 motion parameters consisting of three translations and three rotations, which is then condensed to form a vector of framewise displacement by summing the absolute values of the differentials of the 6 parameters (Power et al., 2012). As a part of the artefactual cleanup process, 24-counfound timeseries derived from the motion estimation (comprised of the 6 rigid body-parameter timeseries, their backwards-looking temporal derivatives, plus all 12 resulting regressors squared) had temporal highpass filtering applied to them and then regressed out of the data aggressively (WU-Minn, 2017). The HCP pipeline ultimately resulted in the removal of spatial and temporal artefacts without removing potentially useful data, particularly relevant for resting-state fMRI data that is particularly sensitive to artefactual corruption across voxels (Smith et al., 2013a).

Only subjects with all four repeated resting-state fMRI sessions (first and second scan sessions with left-right and right-left phase encoding directions), were used for this study. Preparation of masks used in this study has been detailed elsewhere (Loganathan et al., 2020). Briefly, the VS, ECS and VCC were delineated using the binary masks that combined regions of interest (ROIs) from both the Desikan-Killiany (Desikan et al., 2006) and Destrieux (Destrieux et al., 2010) parcellations. All anatomical labels were extracted and merged using the FMRIB Software Library (Smith *et al*., 2004, https://fsl.fmrib.ox.ac.uk/fsl/). The complete list of all anatomical ROIs used to delineate the VS and ECS are shown in Table Supplementary 1, along with their central coordinates given in Montreal Neurological Institute (MNI) space.

The following substance use measures were obtained from the Human Connectome Project Restricted Access Data (Table 1). These substance use and frequency measures were collected using two approaches: a detailed questionnaire prepared specifically for the Human Connectome Project (for short-term alcohol and tobacco use), followed by the Semi-Structured Assessment for the Genetics of Alcoholism (SSAGA) for long-term alcohol, tobacco and illicit drug use. Additionally, parental use history, age, gender and framewise displacement (FD) measures were included as demographic variable.

**Table 1:**
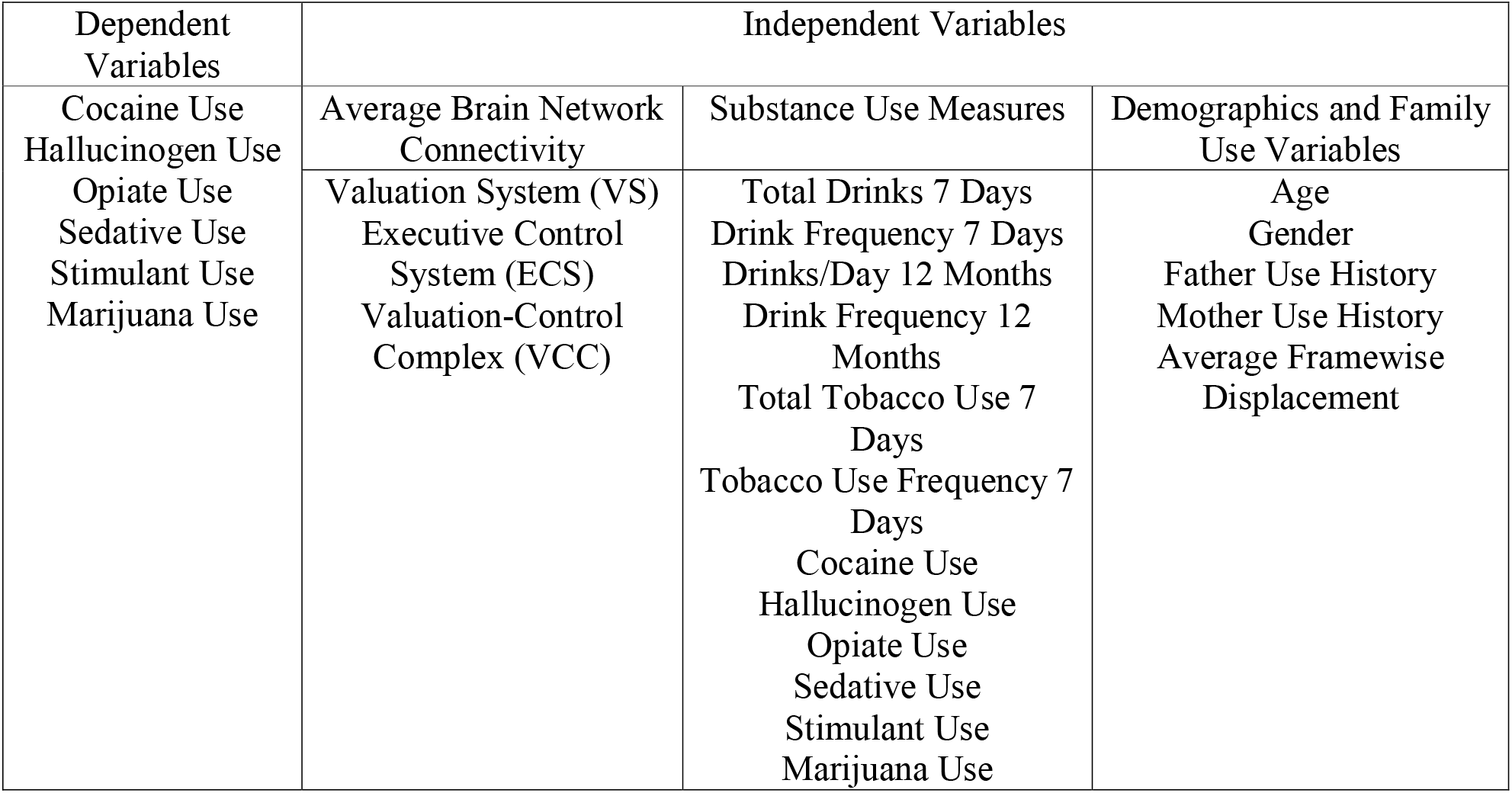
List of drug-based dependent variables and independent variables comprised of both substance use measures, as well as demographic and family use history variables.

| Dependent Variables | Independent Variables |  |  |
| --- | --- | --- | --- |
| Cocaine Use<br>Hallucinogen Use<br>Opiate Use<br>Sedative Use<br>Stimulant Use<br>Marijuana Use | Average Brain Network Connectivity | Substance Use Measures | Demographics and Family Use Variables |
|  | Valuation System (VS)<br>Executive Control System (ECS)<br>Valuation-Control Complex (VCC) | Total Drinks 7 Days<br>Drink Frequency 7 Days<br>Drinks/Day 12 Months<br>Drink Frequency 12 Months<br>Total Tobacco Use 7 Days<br>Tobacco Use Frequency 7 Days<br>Cocaine Use<br>Hallucinogen Use<br>Opiate Use<br>Sedative Use<br>Stimulant Use<br>Marijuana Use | Age<br>Gender<br>Father Use History<br>Mother Use History<br>Average Framewise Displacement |

The rsfMRI signal was averaged over all voxels comprising each ROI (node) of the VS (12 nodes), ECS (20 nodes) and VCC (32 nodes), yielding a regionally averaged signal for each node. The Pearson correlation coefficient in the regionally averaged signals were then computed between all pairs of nodes, resulting in a 12 x 12 (VS), 20 x 20 (ECS) and 32 x 32 (VCC) connectivity matrix for each of the four scan runs (Rest 1-LR, Rest 1-RL, Rest 2-LR, Rest 2-RL). This computation was repeated for each subject. The four connectivity matrices computed from the four rsfMRI sessions for each subject were then averaged to improve the signal-to-noise ratio, yielding a single connectivity matrix for each of the three systems for every subject (see Supplementary Results). These connectivity matrices were then averaged per subject, such that for each of the VS, ECS and VCC, each subject’s connectivity would be represented by a single value. Generalized linear models (GLM) were then computed to correlate the average connectivity of VS, ECS and VCC with each subject’s substance use profile. All dependent variables, independent variables and connectivity measures are standardized to z-scores with mean of 0 and standard deviation of 1. Regression coefficients are therefore representative of SD change in outcome per unit SD change in predictor and may be interpreted as partial correlation coefficients (Bring, 1994). Throughout this study, a total of 288 tests were performed (16 independent variables X 3 brain networks X 6 dependent variables). Each dependent variable was tested against 15 independent variables for all 3 networks (VS, ECS, VCC) producing a single model per dependent variable per network. For the purposes of illustration, let us assume one wishes to model Cocaine use (dependent variable) and its correlation with Valuation System average connectivity (brain network). The 16 independent variables would therefore comprise of Total Drinks 7 Days, Drink Frequency 7 Days, Drinks/Day 12 Months, Drink Frequency 12 Months, Total Tobacco Use 7 Days, Tobacco Use Frequency 7 Days, Hallucinogen, Opiate, Sedatives, Stimulants, Marijuana, Age, Gender, Fathers’ Use History, Mothers’ Use History, Average Framewise Displacement). Multiple comparisons across all models were controlled using the False Discovery Rate using the p-values attributed to each component of each model. p-values < 0.05 post-correction are deemed significant.

Correlation between substance use measures is shown in Table 2. The most highly correlated pairing of substances are opiates and sedatives (r^2^ = 0.725), followed by cocaine and hallucinogens (r^2^ = 0.614). Marijuana use was not strongly correlated with other substances, except with hallucinogens (r^2^ = 0.559). Alcohol (Total Drinks 7 Days, Drink Frequency 7 Days, Drinks/Day 12 Months and Drink Frequency 12 Months) and tobacco measures (Total Tobacco 7 Days, Tobacco Frequency 7 Days) did not show strong correlation with illicit drug use measures. Stronger correlations were reported between alcohol and Marijuana measures as well as tobacco and Marijuana measures, but even these r^2^ values were no higher than 0.326 and 0.336 respectively. Correlations between resting-state functional connectivity and substance use measures are shown in Table 3. Network connectivity of all three systems is weakly correlated with substance use measures, with no r^2^ value higher than 0.1.

**Table 2:** Correlation matrix of substance use measures. Intercepts highlighted in grey.

|  | Total<br>Drinks 7<br>Days | Drink<br>Frequency 7<br>Days | Drinks/Day<br>12 Months | Drink<br>Frequency 12<br>Months | Total<br>Tobacco<br>7 Days | Tobacco<br>Frequency<br>7 Days | Cocaine | Hallucinogens | Opiates | Sedatives | Stimulants | Marijuana |
| --- | --- | --- | --- | --- | --- | --- | --- | --- | --- | --- | --- | --- |
| Total Drinks<br>7 Days | 1.000 |  |  |  |  |  |  |  |  |  |  |  |
| Drink<br>Frequency 7<br>Days | 0.999 | 1.000 |  |  |  |  |  |  |  |  |  |  |
| Drinks/Day<br>12 Months | 0.282 | 0.282 | 1.000 |  |  |  |  |  |  |  |  |  |
| Drink<br>Frequency 12<br>Months | -0.410 | -0.410 | -0.157 | 1.000 |  |  |  |  |  |  |  |  |
| Total<br>Tobacco 7<br>Days | 0.018 | 0.018 | 0.195 | -0.034 | 1.000 |  |  |  |  |  |  |  |
| Tobacco<br>Frequency 7<br>Days | 0.077 | 0.077 | 0.208 | -0.051 | 0.803 | 1.000 |  |  |  |  |  |  |
| Cocaine | 0.119 | 0.119 | 0.197 | -0.051 | 0.181 | 0.189 | 1.000 |  |  |  |  |  |
| Hallucinogens | 0.148 | 0.148 | 0.209 | -0.088 | 0.142 | 0.167 | 0.614 | 1.000 |  |  |  |  |
| Opiates | 0.103 | 0.103 | 0.151 | -0.028 | 0.193 | 0.220 | 0.545 | 0.578 | 1.000 |  |  |  |
| Sedatives | 0.048 | 0.048 | 0.169 | 0.020 | 0.194 | 0.227 | 0.581 | 0.536 | 0.725 | 1.000 |  |  |
| Stimulants | 0.029 | 0.029 | 0.220 | 0.000 | 0.097 | 0.165 | 0.544 | 0.480 | 0.535 | 0.570 | 1.000 |  |
| Marijuana | 0.241 | 0.241 | 0.326 | -0.131 | 0.270 | 0.336 | 0.436 | 0.559 | 0.444 | 0.411 | 0.401 | 1.000 |

**Table 3:**
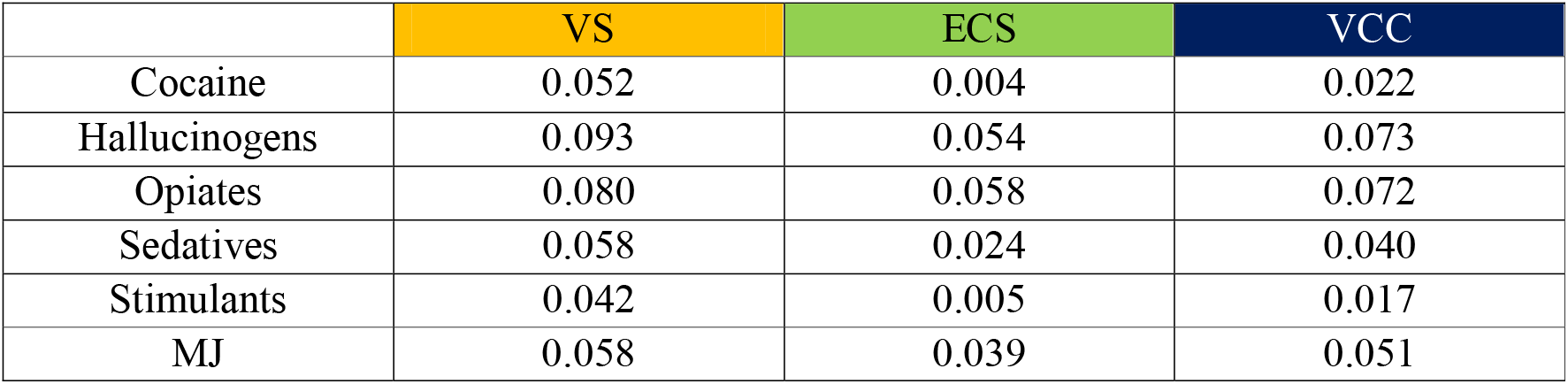
Correlation between VS, ECS and VCC connectivity and substance use measures

|  | VS | ECS | VCC |
| --- | --- | --- | --- |
| Cocaine | 0.052 | 0.004 | 0.022 |
| Hallucinogens | 0.093 | 0.054 | 0.073 |
| Opiates | 0.080 | 0.058 | 0.072 |
| Sedatives | 0.058 | 0.024 | 0.040 |
| Stimulants | 0.042 | 0.005 | 0.017 |
| MJ | 0.058 | 0.039 | 0.051 |

Average connectivity of VS, ECS and VCC is correlated, either independently or via a two-way interaction with substance use (Table 4). VS connectivity is correlated with Sedative use (Estimate=0.04, t=2.3, p=0.03) while ECS connectivity is correlated with Hallucinogen (Estimate=0.08, t=2.4, p=0.025) and Opiate (Estimate=0.05, t=2.3, p=0.03) use. Cocaine use is correlated with VS, ECS and VCC connectivity via two-way interactions with Total Tobacco 7 Days, Hallucinogen use, Opiate use, Sedative use and Fathers’ Use History. Opiate use is correlated with VS (Estimate=0.07, t=3.3, p=0.002), ECS (Estimate=0.08, t=3.6, p=0.0008) and VCC (Estimate=0.1, t=4.4, p<0.00001) connectivity via two-way interaction with Total Tobacco 7 Days. Sedative use is correlated with VS (Estimate=0.04, t=2.6, p=0.017), ECS (Estimate=0.04, t=2.2, p=0.04) and VCC (Estimate=0.04, t=2.3, p=0.03) connectivity via two-way interaction with Drinks/Day 12 Months. Stimulant use is correlated with VS (Estimate=-0.07, t=-2.9 p=0.007), ECS (Estimate=-0.25, t=-7.2, p<0.00001) and VCC (Estimate=-0.16, t=-4.8 p<0.00001) connectivity via two-way interaction with Opiate use. Hallucinogen use is correlated with VS (Estimate=-0.06, t=-2.6, p=0.015) and VCC (Estimate=-0.05, t=7.8, p=0.04) connectivity via two-way interaction with Total Tobacco 7 Days. Opiate use is correlated with ECS (Estimate=0.19, t=5.8, p<0.00001) and VCC (Estimate=0.09, t=3.6, p=0.0009) connectivity via two-way interaction with Hallucinogen use. Sedative use is correlated with ECS (Estimate=0.05, t=2.45, p=0.02) and VCC (Estimate=0.08, t=3.6, p=0.0006) connectivity via two-way interaction with Tobacco Frequency 7 Days. Sedative use is also correlated with ECS (Estimate=0.19, t=7.96 p<0.00001) and VCC (Estimate=0.18, t=8.3, p<0.00001) connectivity via two-way interaction with Opiate use. Sedative use is correlated with VS (Estimate=0.09, t=3.17 p=0.003) and ECS (Estimate=-0.08, t=-2.86, p=0.008) use via two-way interaction with Cocaine use.

**Table 4:** VS, ECS and VCC connectivity is correlated with drug use, both independently (highlighted) and as part of a two-way interaction with other substances. Effect sizes of each GLM are represented by r^2^ values. Estimates displayed are standardized partial coefficients for each component of the GLM. *p*-values shown here are FDR-corrected. Exact p-values are displayed where possible. If the p-value is too small (less than 0.00001), then it is displayed as p<0.00001. Father, Fathers’ Use History; Mother, Mothers’ Use History; E, Estimate

| Dependents | VS | ECS | VCC |
| --- | --- | --- | --- |
| Cocaine | $r^2 = 0.7227$<br>VS – Total Tobacco 7 Days ($E=0.07$, $t=2.92$ $p=0.007$)<br>VS – Tobacco Frequency 7 Days ($E=0.08$, $t=2.53$ $p=0.02$)<br>VS – Hallucinogens ($E=0.12$, $t=3.79$ $p=0.00038$)<br>VS – Opiates ($E=-0.2$, $t=-6.1$, $p<0.00001$)<br>VS – Sedatives ($E=-0.06$, $t=-2.83$ $p=0.0085$)<br>VS – Father ($E=-0.07$, $t=-3.3$ $p=0.0021$) | $r^2 = 0.7208$<br>ECS – Total Tobacco 7 Days ($E=0.06$, $t=2.84$ $p=0.08$)<br>ECS – Hallucinogens ($E=0.17$, $t=4.65$ $p=0.00001$)<br>ECS – Opiates ($E=-0.21$, $t=-5.82$, $p<0.00001$)<br>ECS – Sedatives ($E=-0.06$, $t=-2.66$ $p=0.01$)<br>ECS – Father ($E=-0.06$, $t=-3.3$, $p=0.002$) | $r^2 = 0.7198$<br>VCC – Total Tobacco 7 Days ($E=0.07$, $t=3.0$ $p=0.005$)<br>VCC – Hallucinogens ($E=0.18$, $t=5.4$ $p<0.00001$)<br>VCC – Opiates ($E=-0.20$, $t=-5.7$, $p<0.00001$)<br>VCC – Sedatives ($E=-0.05$, $t=-2.22$, $p=0.04$)<br>VCC – Father ($E=-0.06$, $t=-3.0$, $p=0.006$) |
| Hallucinogens | $r^2 = 0.6707$<br>VS – Total Tobacco 7 Days ($E=-0.06$, $t=-2.6$, $p=0.015$)<br>VS – Tobacco Frequency 7 Days ($E=0.14$, $t=4.8$ $p<0.00001$)<br>VS – Cocaine ($E=-0.21$, $t=-6.5$ $p<0.00001$)<br>VS – Opiates ($E=0.22$, $t=7.15$, $p<0.00001$)<br>VS – Stimulants ($E=0.006$, $t=2.4$, $p<0.02$) | $r^2 = 0.7856$<br>ECS ($E=0.08$, $t=2.4$, $p=0.025$)<br>ECS – Tobacco Frequency 7 Days ($E=0.03$, $t=2.37$ $p=0.03$)<br>ECS – Gender ($E=-0.1$, $t=-2.24$, $p=0.03$) | $r^2 = 0.6612$<br>VCC – Total Tobacco 7 Days ($E=-0.05$, $t=-2.2$ $p=0.04$)<br>VCC – Opiates ($E=0.19$, $t=7.8$ $p<0.00001$) |
| Opiates | $r^2 = 0.7056$<br>VS – Drink Frequency 12 Months ($E=-0.05$, $t=0.19$, $p=0.03$)<br>VS – Total Tobacco 7 Days ($E=0.07$, $t=3.3$, $p=0.002$)<br>VS – Tobacco Frequency 7 Days ($E=0.1$, $t=5.9$, $p<0.00001$) | $r^2 = 0.7137$<br>ECS ($E=0.05$, $t=2.3$, $p=0.03$)<br>ECS – Total Tobacco 7 Days ($E=0.08$, $t=3.6$, $p=0.0008$)<br>ECS – Hallucinogens ($E=0.19$, $t=5.8$, $p<0.00001$)<br>ECS – Sedatives ($E=-0.07$, $t=-3.1$, $p=0.005$) | $r^2 = 0.6996$<br>VCC – Drinks/Day 12 Months ($E=-0.04$, $t=-2.3$ $p=0.024$)<br>VCC – Total Tobacco 7 Days ($E=0.1$, $t=4.4$, $p<0.00001$)<br>VCC – Hallucinogens ($E=0.09$, $t=3.6$, $p=0.0009$) |
| Sedatives | $r^2 = 0.7797$<br>VS ($E=0.04$, $t=2.3$, $p=0.03$)<br>VS – Drinks/Day 12 Months ($E=0.04$, $t=2.6$, $p=0.017$)<br>VS – Cocaine ($E=0.09$, $t=3.17$ $p=0.003$)<br>VS – Hallucinogens ($E=-0.11$, $t=-4.42$ $p<0.00001$)<br>VS – Opiates ($E=0.11$, $t=5.7$ $p<0.00001$) | $r^2 = 0.806$<br>ECS – Drinks/Day 12 Months ($E=0.04$, $t=2.2$, $p=0.04$)<br>ECS – Tobacco Frequency 7 Days ($E=0.05$, $t=2.45$, $p=0.02$)<br>ECS – Cocaine ($E=-0.08$, $t=-2.86$, $p=0.008$)<br>ECS – Opiates ($E=0.19$, $t=7.96$ $p<0.00001$) | $r^2 = 0.8083$<br>VCC – Drinks/Day 12 Months ($E=0.04$, $t=2.3$, $p=0.03$)<br>VCC – Tobacco Frequency 7 Days ($E=0.08$, $t=3.6$, $p=0.0006$)<br>VCC – Hallucinogens ($E=-0.14$, $t=-4.7$, $p<0.00001$)<br>VCC – Opiates ($E=0.18$, $t=8.3$, $p<0.00001$) |
| Stimulants | $r^2 = 0.6265$<br>VS – Drink Frequency 12 Months ($E=-0.05$, $t=-2.5$ $p=0.022$)<br>VS – Opiates ($E=-0.07$, $t=-2.9$ $p=0.007$)<br>VS – Father ($E=-0.07$, $t=-2.4$ $p=0.025$) | $r^2 = 0.6392$<br>ECS – Tobacco Frequency 7 Days ($E=0.08$, $t=3.0$ $p=0.005$)<br>ECS – Hallucinogens ($E=-0.1$, $t=-3.2$ $p=0.0033$)<br>ECS – Opiates ($E=-0.25$, $t=-7.2$, $p<0.00001$) | $r^2 = 0.6288$<br>VCC – Tobacco Frequency 7 Days ($E=0.06$, $t=2.9$, $p=0.04$)<br>VCC – Hallucinogens ($E=-0.09$, $t=-3.1$ $p=0.0035$)<br>VCC – Opiates ($E=-0.16$, $t=-4.8$, $p<0.00001$) |
| Marijuana | $r^2 = 0.4715$<br>VS – Tobacco Frequency 7 Days ($E=-0.5$, $t=-2.5$, $p=0.019$)<br>VS – Stimulants ($E=0.7$, $t=2.62$, $p=0.015$) | $r^2 = 0.4703$<br>ECS – Drink Frequency 12 Months ($E=0.73$, $t=3.5$ $p=0.00094$) | $r^2 = 0.4732$<br>VCC – Drink Frequency 12 Months ($E=0.7$, $t=3.4$, $p=0.0018$)<br>VCC – Opiates ($E=-0.8$, $t=-3.0$, $p=0.005$)<br>VCC – Sedatives ($E=0.6$, $t=2.5$, $p=0.018$) |

Research suggests that substance use may stem in part from a disrupted decision-making system stemming from aberrant valuation of drugs (Zhai et al., 2015). The value of drugs varies between individuals, driven by hedonic sensations in some or pain relief in others (Hogarth, 2020). These underlying motivations increase the value of drugs and bias choice towards their use (Hogarth and Field, 2020). Additionally, connections both within and between the Valuation and Executive Control System act as dependency risk and resilience markers (Ersche et al., 2020). Here, we correlated the average network connectivity of the Valuation and Executive Control Systems, as well as the Valuation-Control Complex with drug use in the brains of healthy young adults. Common co-use patterns and their network correlations can thus be inferred based on trends observed in this study. Understanding the needs of one who takes addictive substances with respect to a desired internal state (e.g., euphoria or numbness) represents the first step to improving existing forms of interventions, with greater focus on individual needs that are being met via substance use.

Correlations are observed between combinations of addictive substances, for example Sedatives and Cocaine, Opiates and Sedatives. Highly correlated pairs of drugs reflect popular combinations used to achieve a particular state (euphoria, numbness etc) (Floyd et al., 2010; Laursen et al., 2016). Thus, two-way interactions involving substance use measures and average network connectivity of choice networks may signify the high value placed on these combinations to achieve said state. Stimulating and sedating substances are commonly co-used due to their synergistic interactions. Stimulant-type substances (cocaine, stimulants like amphetamine) energize the user, producing longer-lasting euphoria while off-setting sedating effects (Hernández-López et al., 2002). Sedative-type substances (alcohol, tobacco, marijuana, pain-killing sedatives, heroin) calm the user, ameliorating unpleasant side-effects of stimulant use such as anxiety and aggression (Fisk et al., 2011).

Tobacco and alcohol (Drink Frequency, Total Drinks and Drinks/Day) use measures form two-way interactions with VS, ECS and VCC average network connectivity for all six substances tested as dependent variables (Cocaine, Hallucinogens, Opiates, Sedatives, Stimulants, Marijuana). Tobacco and alcohol both possess sedating qualities (Fultz et al., 2021; Kishore, 2014; Walker, 1980) and are considered gateway substances that pave the way towards the development of illicit drug consumption (Kandel and Kandel, 2015), underpinning poly-substance use (Anderson et al., 2018; Jongenelis et al., 2019; Moss et al., 2014). Stimulants complement alcohol to produce longer-lasting euphoria while off-setting the sedating qualities of alcohol (Hernández-López et al., 2002), reducing the anxiety and agitation (Fisk et al., 2011; Knackstedt and Ettenberg, 2005). Alcohol and sedating substances (opiates, sedatives, marijuana) are often co-used for pain management (Downey et al., 2013; Hood et al., 2020; Ronen et al., 2010; Wilson et al., 2020), albeit at the expense of cognitive control over reward-driven behaviour (Crean et al., 2011; Müller-Oehring et al., 2013). Similarly, tobacco use sensitizes the reward centres of the brain making one susceptible to other drugs (Kandel and Kandel, 2015). Tobacco is thought to encourage cocaine and opiate use (Cross et al., 2017; Mello et al., 1980; Stark and Campbell, 1993) by stimulating the reward centres of the brain (Cross et al., 2017; Nuechterlein et al., 2016). The findings of the current study support existing evidence regarding the involvement of alcohol and tobacco in poly-drug use, as these two substances form two-way interactions with average connectivity of VS, ECS and VCC for all illicit drugs (Cocaine, Hallucinogens, Opiates, Sedatives, Stimulants, Marijuana). Given the “gateway” nature of these two substances (Kandel and Kandel, 2015), particularly among young adults (Laursen et al., 2016; McKetin et al., 2014), it is theorized that these participants may have started using alcohol and/or tobacco to achieve an internal state of euphoria or numbness, but began to incorporate illicit drugs to boost the strength of the desired sensation.

Two-way interactions between all three networks and Opiates in Cocaine use could be indicative of “speedball”, a cocaine-heroin mixture that induces enhanced feelings of euphoria (Lacy et al., 2014), while reducing the aggression and anxiety (Fisk et al., 2011). A similar effect may underpin other two-way interactions between all three networks and sedating substances (Opiates, Sedatives) with Stimulants and vice versa. Studies revealed ECS involvement in regulation of attentional processing of external cues (Sarter et al., 2001) including auditory (Napoli et al., 2021; Plakke et al., 2013) and visual stimuli (Geliebter et al., 2016). Both stimulant and cocaine use are capable of enhancing the senses (Devonshire et al., 2007; Oken et al., 2006) and inducing euphoria (Bellone et al., 2020; Dlugos et al., 2011) by increasing activation within the ECS (Jan et al., 2014) and VS (Crane et al., 2018) respectively.

Co-use of sedating substances are commonly used for pain management (Peele, 2016) especially when a single substance is found to be inadequate for the purpose (Wilson et al., 2020). The VS acts as the middle ground for processing both pleasure and pain (Leknes and Tracey, 2008). Connections between the VS and ECS regions is observed in patients experiencing chronic pain (Jiang et al., 2016). In the current study, VS, ECS and VCC each form a two-way interaction with alcohol in Sedatives use, suggesting a pain-killing combination. Two-way interactions involving VS, ECS and VCC connectivity and sedating substances (Marijuana, Sedatives) when paired with other sedating substances (such as alcohol) could indicate these combinations of substances are being used for pain management.

This study is not without limitations. There are various types of stimulants, sedatives and hallucinogens, some of which have been manufactured to have pharmacological properties of other abusable substances (Klega and Keehbauch, 2018). Given that each drug has its own mechanism of action that targets specific receptors or pathways, information regarding the specific types of drugs used may help shed additional light on the relationships between network connectivity and poly-drug use measures. Additionally, global signal regression was not performed in this study. The fMRI scans used in this study have been minimally pre-processed by the HCP team to ensure minimal information loss (Glasser et al., 2013). Global signal regression is deemed too aggressive for use with HCP imaging data and never implemented. This method may introduce artifactual changes in correlations patterns, fundamentally altering interregional correlations. These changes are thought to be dependent on the underlying true interregional correlation pattern (Anderson et al., 2011; Saad et al., 2012).

In conclusion, we correlated the average network connectivity of the Valuation and Executive Control Systems, as well as the Valuation-Control Complex with drug use in the brains of healthy young adults. Common co-use patterns and their network correlations can thus be inferred based on trends observed in this study. Two-way interactions involving substance use measures and average network connectivity of choice networks may signify the high value placed on these combinations to achieve said state. Particularly, tobacco and alcohol use measures form interactions with VS, ECS and VCC average network connectivity across all forms of illicit substances (Cocaine, Hallucinogens, Opiates, Sedatives, Stimulants and Marijuana), suggesting that participants may have started using alcohol and/or tobacco initially but began to incorporate illicit drugs to enhance their experience. Additionally, two-way interactions involving sedating substances (e.g. Opiates) with connectivity of all 3 networks is observed in stimulant-type substances (e.g. Cocaine) and vice-versa, suggesting a combination aimed at increasing feelings of euphoria while dampening anxiety and aggression. Finally, connectivity of all three networks each form a two-way interaction with alcohol among sedating substances, suggesting a combination for pain management. These results provide early evidence linking functional connectivity of the VS and ECS with poly-substance use, enabling us to infer common combinations of substance co-use and associated neural patterns.

## Data Availability

The date referred to in the manuscript was derived from the Human Connectome Project Healthy Young Adult Study

## Acknowledgements

Kavinash Loganathan performed analysis, interpreted results and prepared this manuscript, Jinglei Lv was involved in experimental design, assisted in analysis and interpretation of results, Eric Tatt Wei Ho performed analysis and interpreted results, Vanessa Cropley assisted with brain mask generation and provided technical feedback, Andrew Zalesky was involved in experimental design, provided technical feedback and assisted with result interpretation.

## Supplementary Methods

The following table shows the list of regions of interest (ROIs) and their center of gravity (in mm) used in the construction of the valuation, executive control and valuation-control complex masks.

**Supplementary Table 1:** List of ROIs and their corresponding center of gravity (in mm) used to assemble the Valuation and Control system brain masks. LH, left hemisphere; RH, right hemisphere; OFC, orbitofrontal cortex; PC, posterior cingulate; FG, frontal gyrus; MC, middle cingulate; lat, lateral; medial, med; vent, ventral; dors, dorsal; sup, superior; inf, inferior; ros, rostral; caud, caudal; mid, middle; ant, anterior; pos, posterior; opercularis, pars opercularis; triangularis, pars triangularis; orbitalis, pars orbitalis

| System | Functional region | ROI | x | y | z |
| --- | --- | --- | --- | --- | --- |
| Valuation | vmPFC | LH lat OFC | -23.83 | 30.67 | -17.77 |
|  |  | RH lat OFC | 23.00 | 31.98 | -18.46 |
|  |  | LH med OFC | -6.80 | 33.49 | -16.02 |
|  |  | RH med OFC | 6.32 | 37.38 | -16.73 |
|  | PCC | LH dors PC | -4.07 | -39.89 | 29.19 |
|  |  | RH dors PC | 5.33 | -36.50 | 32.92 |
|  |  | LH vent PC | -7.19 | -47.48 | 8.64 |
|  |  | RH vent PC | 8.55 | -49.22 | 7.49 |
|  | Amygdala | LH amygdala | -23.18 | -4.93 | -19.56 |
|  |  | RH amygdala | 24.20 | -3.75 | -19.71 |
|  | Accumbens | LH accumbens | -8.79 | 11.65 | -6.46 |
|  |  | RH accumbens | 9.53 | 12.09 | -7.23 |
| Control | PPC | LH inf parietal | -41.27 | -68.97 | 34.93 |
|  |  | RH inf parietal | 47.51 | -60.80 | 33.32 |
|  |  | LH sup parietal | -20.92 | -66.90 | 48.70 |
|  |  | RH sup parietal | 24.23 | -61.60 | 52.12 |
|  | dlPFC | LH sup FG | -9.79 | 27.31 | 43.89 |
|  |  | RH sup FG | 12.23 | 30.84 | 43.45 |
|  |  | LH ros mid FG | -31.16 | 48.05 | 17.58 |
|  |  | RH ros mid FG | 34.05 | 46.79 | 17.17 |
|  |  | LH caud mid FG | -35.85 | 11.95 | 47.34 |
|  |  | RH cau mid FG | 38.06 | 11.21 | 48.12 |
|  | vlPFC | LH opercularis | -47.86 | 18.01 | 13.92 |
|  |  | RH opercularis | 48.58 | 15.55 | 11.08 |
|  |  | LH orbitalis | -42.11 | 38.11 | -13.59 |
|  |  | RH orbitalis | 44.08 | 39.89 | -12.09 |
|  |  | LH triangularis | -46.47 | 35.21 | 1.73 |
|  |  | RH triangularis | 49.08 | 33.38 | 6.56 |
|  | dACC | LH ant MC | -7.19 | 12.19 | 34.99 |
|  |  | RH ant MC | 7.96 | 15.22 | 36.09 |
|  |  | LH pos MC | -7.50 | -14.65 | 40.42 |
|  |  | RH pos MC | 7.98 | -8.77 | 42.24 |

## Supplementary Results

**Supplementary Table 2:** Descriptive statistics of the variables featured in this study.

| Variable | Type | Mean | Standard Deviation | Range |
| --- | --- | --- | --- | --- |
| Age | Continuous | 28.73 | 3.71 | 15 |
| Cognition scores | Continuous | 114.08 | 20.25 | 93.32 |
| Total Drinks 7 Days | Continuous | 4.89 | 7.08 | 55 |
| Number of Days Drank 7 Days | Continuous | 1.68 | 1.78 | 7 |
| Total Tobacco 7 Days | Continuous | 7.4 | 23.06 | 175 |
| Number of Days Any Tobacco 7 Days | Continuous | 0.98 | 2.28 | 7 |
| SSAGA Alcohol 12 Drinks/Day | Ordinal | 2.28 | 1.57 |  |
| SSAGA Alcohol 12 Drink Frequency | Ordinal | 4.29 | 1.54 |  |
| Cocaine | Ordinal | 0.22 | 1.16 |  |
| Hallucinogens | Ordinal | 0.27 | 0.9 |  |
| Opiates | Ordinal | 0.23 | 0.92 |  |
| Sedatives | Ordinal | 0.17 | 0.82 |  |
| Stimulants | Ordinal | 0.21 | 0.88 |  |
| MJ | Ordinal | 1.41 | 1.69 |  |
| Father's History of Drug Use | Binary | 0.13 | 0.34 |  |
| Mother's History of Drug Use | Binary | 0.03 | 0.18 |  |

A multicollinearity test was performed among all substance use measures (Supplementary Table 3). Results do not indicate the presence of multicollinearity among these variables. Among the substance use measures, no condition index scored more than 15, while no variance inflation factor (VIF) readings are above 10.

**Supplementary Table 3:**
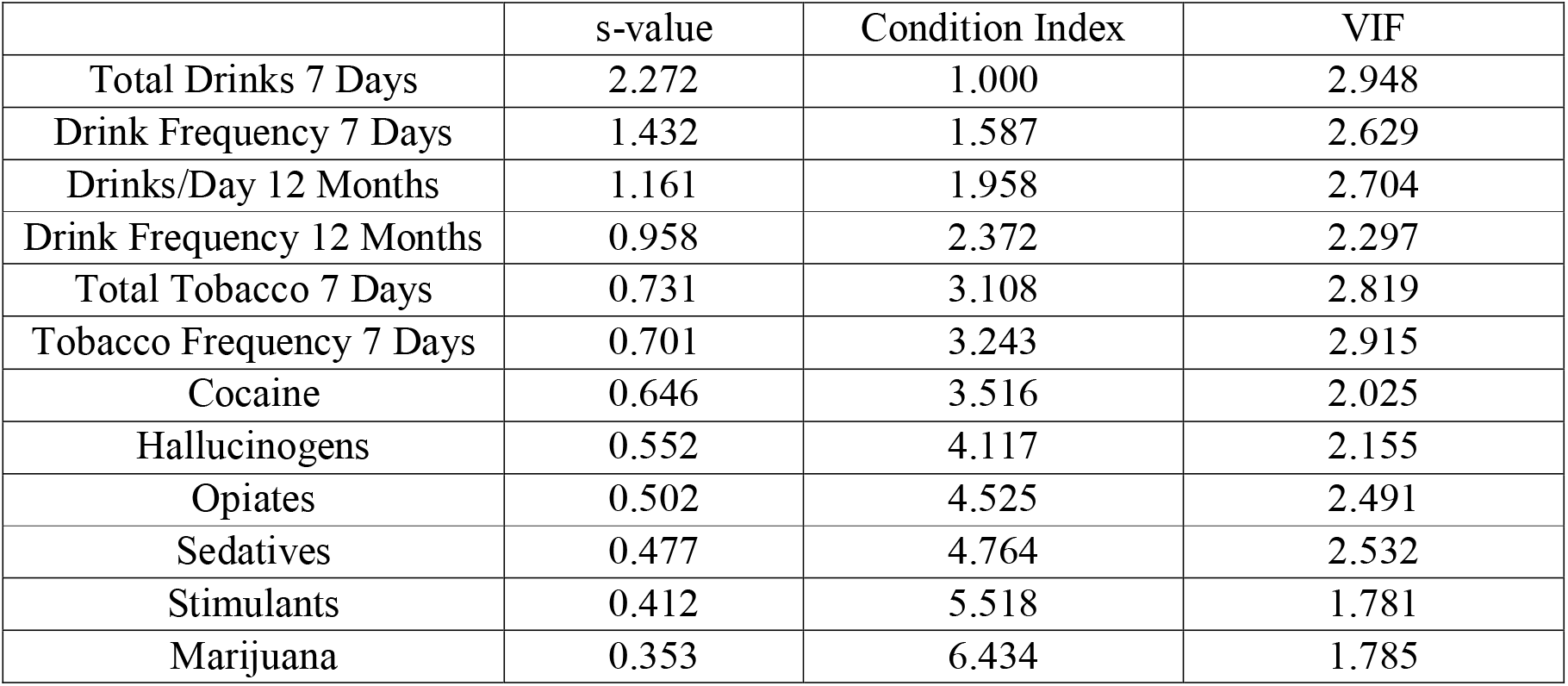
Multicollinearity test between substance use measures

The following figures show average VS, ECS and VCC connectivity in the brains of Human Connectome Project Healthy Young Adult participants used in this study. Connectivity within the VS was higher between the regions of the vmPFC and PCC (Supplementary Figure 1A). Connectivity within the ECS was higher between regions of the PPC, DLPFC and VLPFC (Supplementary Figure 1B). In the VCC, connectivity between the ECS and VS regions was higher between the PPC-DLPFC and the vmPFC-PCC as well as between the lateral OFC and VLPFC (Supplementary Figure 2).

**Supplementary Figure 1:**
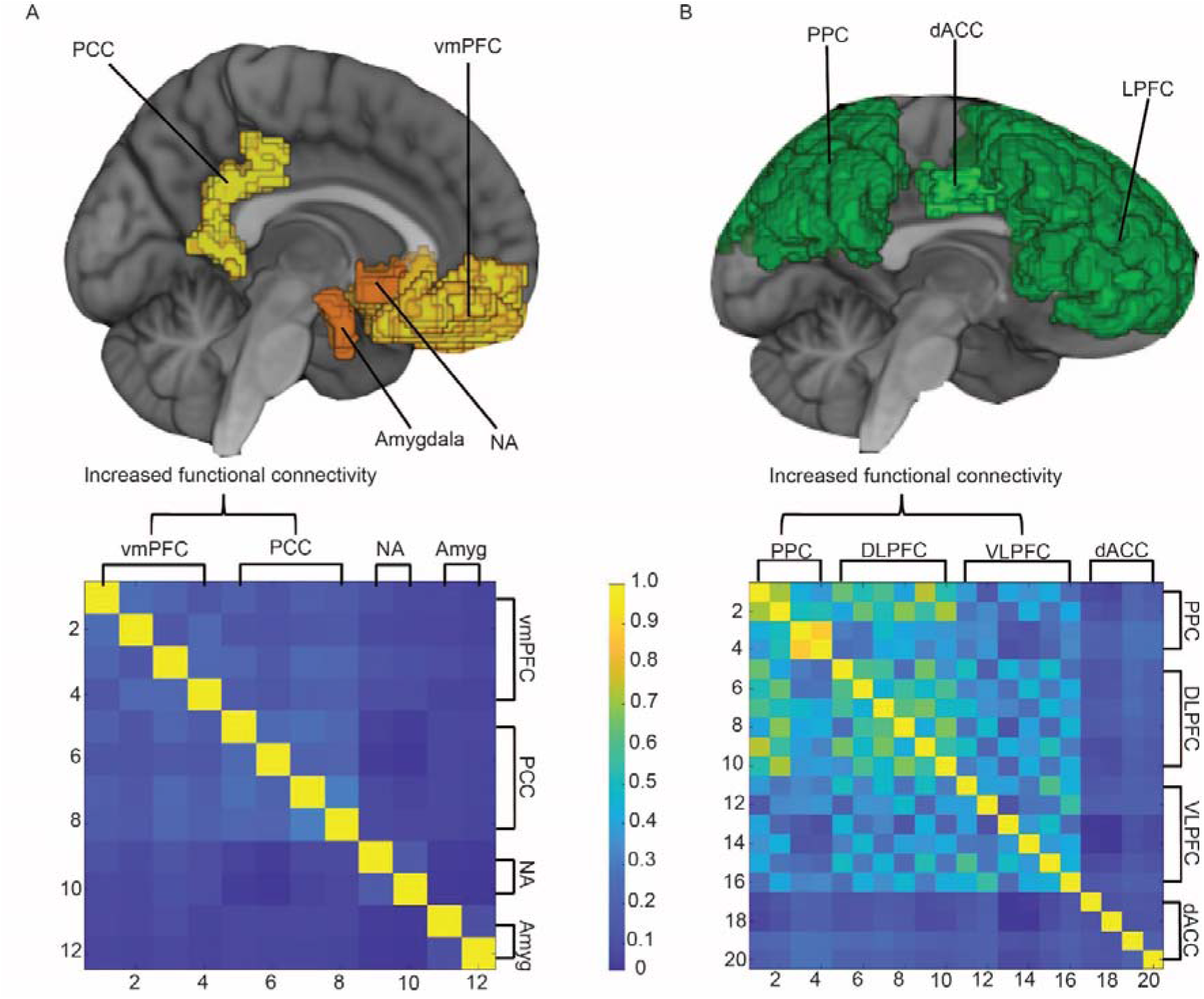
Valuation system component regions (upper) and resting-state functional connectivity matrix (lower) in the human brain (A). Control System component regions (upper) and resting-state functional connectivity matrix (lower) in the human brain (B).

**Supplementary Figure 2:**
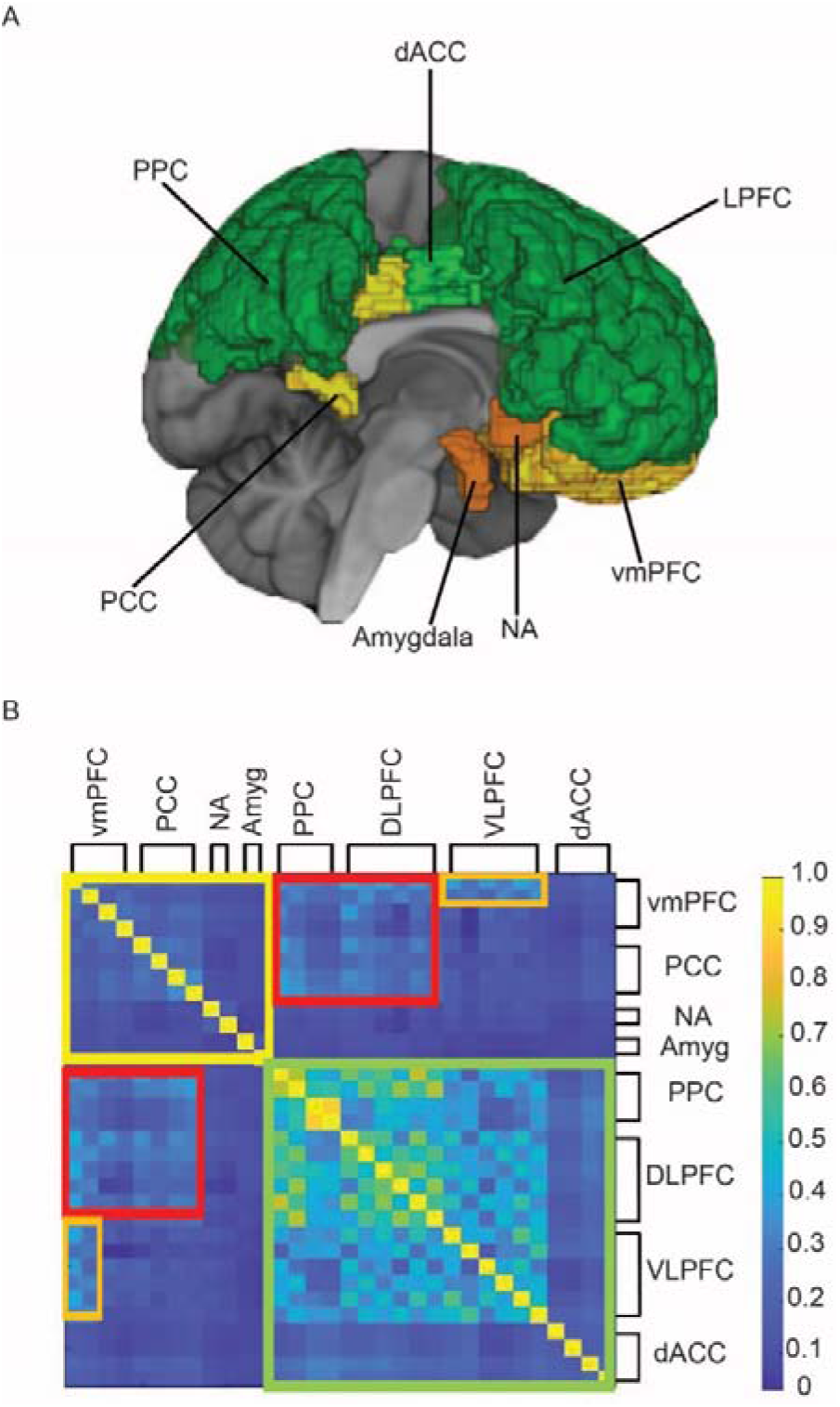
Valuation system (yellow-orange) and control system (green) component regions in the human brain. The cortical regions (vmPFC, PCC) of the valuation system are labelled yellow, while the subcortical regions (NA, amygdala) are labelled orange). The valuation-control complex is a combination of the valuation and control systems (A). Resting-state connectivity matrix of the valuation-control complex. Connections within the valuation system are shown in yellow, the control system in green. PPC-DLFPC connections with vmPFC-PCC are highlighted in red, VLPFC-lateral OFC connections are highlighted in orange (B).

Given that head motion both affects functional connectivity measures (Ciric et al., 2018) and is affected by substance use (Givens, 2016), average framewise displacement values were correlated with both average functional connectivity of VS, ECS and VCC (Supplementary Table 4) as well as substance use measures (Supplementary Table 5). Strong correlations were not observed between average framewise displacement with either average connectivity or substance use measures with only tobacco measures (Total Tobacco 7 Days and Tobacco Frequency 7 Days) registering r values higher than 0.1.

**Supplementary Table 4:** Correlations between average framewise displacement for each subject and average functional connectivity of the VS, ECS and VCC

| Network | Correlation (r) | p-value |
| --- | --- | --- |
| Valuation System | -0.010 | 0.3765 |
| Executive Control System | 0.090 | 0.0023 |
| Valuation-Control Complex | 0.080 | 0.0059 |

**Supplementary Table 5:** Correlations between average framewise displacement for each subject and substance use measures.

| Substance Use Measure | Correlation (r) | p-value |
| --- | --- | --- |
| Total Drinks 7 Days | -0.019 | 0.275 |
| Drink Frequency 7 Days | -0.019 | 0.275 |
| Drinks/Day 12 Months | 0.090 | 0.0023 |
| Drink Frequency 12 Months | 0.053 | 0.0476 |
| Total Tobacco 7 Days | 0.187 | <0.0001 |
| Tobacco Frequency 7 Days | 0.150 | <0.0001 |
| Cocaine | 0.040 | 0.1041 |
| Hallucinogens | 0.029 | 0.1808 |
| Opiates | 0.053 | 0.0476 |
| Sedatives | 0.062 | 0.0255 |
| Stimulants | -0.0039 | 0.4512 |
| Marijuana | 0.0798 | 0.006 |

**Supplementary Table 6:** GLM equations and their corresponding r^2^- and p-values (Valuation System). To enable comparison of effect sizes across relationships, all predictor and outcome variables were standardized to *Z* scores with a mean of 0 and an SD of 1. Regression coefficients are therefore representative of SD change in outcome per unit SD change in predictor and may be interpreted as partial correlation coefficients. “7” (e.g. Total Tobacco 7, Drink Frequency 7) indicates 7 days; “12” (e.g. Drinks/Day 12) indicates 12 months.

| Equation | $r^2$ | p-value |
| --- | --- | --- |
| cocaine ~ 0.19443 + 0.068862(VS – Total Tobacco 7) + 0.075595(VS – Tobacco Frequency 7) + 0.11683(VS – Hallucinogens) - 0.19901(VS – Opiates) – 0.05976(VS – Sedatives) - 0.07448(VS – Father) + 0.059324(Age – Total Drinks 7) + 0.16187(Age – Tobacco Frequency 7) + 0.14899(Age – Hallucinogens) + 0.06162(Total Drinks 7 – Drinks/Day 12) + 0.10987(Total Drinks 7 – Tobacco Frequency 7) + 0.11095(Total Drinks 7 – Hallucinogen) – 0.31816(Total Drinks 7 – Hallucinogens) – 0.31816(Total Drinks 7 – Stimulants) + 0.092877(Total Drinks 7 – Marijuana) - 0.1899(Drink Frequency 7 – Tobacco Frequency 7) – 0.1584(Drink Frequency 7 – Hallucinogens) + 0.36753(Drink Frequency 7 – Sedatives) + 0.16412(Drinks/Day 12 – Total Tobacco 7) + 0.035364(Drink Frequency 7 – Total Tobacco 7) + 0.093752(Drink Frequency 7 – Tobacco Frequency 7) - 0.19016 (Drink Frequency 7 – Sedatives) + 0.10236(Total Tobacco 7 – Tobacco Frequency 7) – 0.078794(Total Tobacco 7 – Hallucinogens) + 0.090749(Total Tobacco 7 – Sedatives) – 0.51708(Total Tobacco 7 – Mother) + 0.083325(Tobacco Frequency 7 – Sedatives) + 0.21259(Tobacco Frequency 7 – Father) - 0.6422(Tobacco Frequency 7 – Mother) - 0.030834(Hallucinogens – Opiates) + 0.71706(Hallucinogens – Sedatives) – 0.32437(Hallucinogens – Father) - 2.2652(Hallucinogens – Mother) + 0.28473(Hallucinogens – Gender) – 0.10987(Opiates – Sedatives) – 0.1689(Opiates – Marijuana) + 1.2938(Opiates – Mother) + 0.24486(Opiates – Gender) – 0.054253(Sedatives – Marijuana) – 0.59964(Sedatives – Gender) + 0.11409(Stimulants – Mother) | 0.7227 | 2.96e-230 |
| hallucinogens ~ -1.6301 – 0.06369(VS – Total Tobacco 7) + 0.13897(VS – Tobacco Frequency 7) - 0.22626 (VS – Cocaine) + 0.21878(VS – Opiates) + 0.0062508(VS – Stimulants) + 0.05203(Age – Drinks/Day 12) + 0.20312(Age – Cocaine) – 0.12568(Age – Opiates) + 0.10987(Total Drinks 7 – Cocaine) – 0.065655(Total Drinks 7 – Marijuana) – 0.1457(Drink Frequency 7 – Cocaine) + 0.0087609(Drink Frequency 7 – Sedatives) + 0.141(Drinks/Day 12 – Opiates) – 0.094957(Drinks/Day 12 – Sedatives) + 0.10969(Drink Frequency 12 – Tobacco Frequency 7) + 0.070751(Drink frequency 12 – Opiates) – 0.012169(Drink Frequency 12 – Stimulants) – 0.091478(Total Tobacco 7 – Opiates) – 0.066949(Total Tobacco 7 – Sedatives) + 0.062218(Tobacco Frequency 7 – Marijuana) – 0.081708(Cocaine – Sedatives) + 0.11838(Cocaine – Marijuana) + 0.4122(Cocaine – Father) + 0.42785(Cocaine – Gender) – 0.15891(Opiates – Father) + 4.9378(Opiates – Mother) + 0.013022(Sedatives – Stimulants) – 3.465(Sedatives – Mother) – 0.26538(Sedatives – Gender) + 0.014303(Stimulants – Gender) | 0.6707 | 1.98e-208 |
| opiates ~ -0.077541 – 0.047223 (VS – Drink Frequency 7) + 0.07354(VS – Total Tobacco 7) + 0.11462(VS – Tobacco Frequency 7) – 0.036665(VS – Father) + 0.065478(Age – Hallucinogens) – 0.17539(Age – Sedatives) + 0.042794(Total Drinks 7 – Cocaine) + 0.19854(Total Drinks – Cocaine) – 0.19573(Drink Frequency 7 – Cocaine) + 0.065957(Drink Frequency 7 – Hallucinogens) – 0.061074(Drinks/Day 12 – Total Tobacco 7) – 0.063834(Drinks/Day 12 – Cocaine) + 0.083089(Drinks/Day 12 – Hallucinogens) – 0.14065(Drinks/Day 12 – Hallucinogens) – 0.071936(Drink Frequency 2 – Cocaine) – 0.12058(Drink Frequency 12 – Hallucinogens) + 0.16286(Drink Frequency 12 – Hallucinogens) – 0.047018(Total Tobacco 7 – Tobacco Frequency 7) + 0.14016(Total Tobacco 7 – Cocaine) – 0.082762(Tobacco Frequency 12 – Hallucinogens) + 0.04842(Tobacco Frequency 12 – Sedatives) – 2.2096(Cocaine – Mother) – 0.19427(Cocaine – Gender) + 0.0052223(Hallucinogens – stimulants) – 0.13244(Hallucinogens – Marijuana) + 3.216(Sedatives – Mother) + 0.31369(Stimulants – Gender) | 0.7056 | 1.65e-228 |
| sedatives ~ 0.022372 + 0.044131(VS – Drinks/Day 12) + 0.05633(VS – Tobacco Frequency 7) + 0.087239(VS – Cocaine) – 0.11234(VS – Hallucinogens) + 0.1094(VS – Opiates) + 0.035095(Age – Drink Frequency 7) – 0.13456(Age – Cocaine) + 0.18204 (Age – Opiates) – 0.038749(Total Drinks 7 – Drink Frequency 7) – 0.29372(Total Drinks 7 – Hallucinogens) + 0.37067(Total Drinks 7 – Opiates) – 0.95341(Total Drinks 7 – Mother) – 0.062641(Drink Frequency 7 – Tobacco Frequency 7) + 0.089163(Tobacco Frequency 7 – Hallucinogens) – 0.40739(Drink Frequency 7 – Opiate) + 0.034425(Drinks/Day 12 – Drink Frequency 12) + 0.040748(Drinks/Day 12 – Total Tobacco 7) – 0.1544(Drinks/Day 12 – Tobacco Frequency 7) – 0.095024(Drinks/Day 12 – Cocaine) + 0.41269(Drinks/Day 12 – Hallucinogens) – 0.21047(Drinks/Day 12 – Sedatives) – 0.16845(Drink Frequency 12 – Tobacco Frequency 7) + 0.1052(Total Tobacco 7 – Tobacco Frequency 7) – 0.078702(Total Tobacco 7 – Sedatives) – 0.030747(Total Tobacco 7 – Father) + 0.050822(Tobacco Frequency 7 – Cocaine) – 0.12095(Tobacco Frequency 7 – Hallucinogens) + 0.058513(Tobacco frequency 7 – Opiates) + 5.4137(Tobacco Frequency 7 – Mother) – 0.24899(Tobacco Frequency 7 – Gender) + 0.10356(Cocaine – Hallucinogens) – 0.097371(Cocaine – Opiates) – 0.012427(Cocaine – stimulants) + 0.1893(Cocaine – Gender) + 0.031576(Hallucinogens – Opiates) + 0.01372(Hallucinogens – Stimulants) + 0.26187(Hallucinogens – Marijuana) + 0.25412(Hallucinogens – Gender) – 0.0076042(Opiates – Stimulants) – 0.12105(Opiates – Marijuana) – 3.9793(Opiates – Mother) – 0.58812(Marijuana – Mother) | 0.7797 | 1.24e-293 |
| stimulants ~ 0.088602 – 0.050192(VS – Drink Frequency 12) – 0.0724(VS – Opiates) – 0.060044(VS – Father) + 0.13787(Age – Opiates) + 0.078607(Total Drinks 7 – Drinks/Day 12) – 0.22526(Total Drinks 7 – Drink Frequency 12) + 0.3232(Total Drinks 7 – Opiates) + 0.0088373(Total Drinks 7 – Stimulants) – 0.8904(Total Drinks 7 – Mother) + 0.15143(Drink Frequency 7 – Tobacco Frequency 7) – 0.36282(Drink Frequency 7 – Opiates) + 0.29091(Drinks/Day 12 – Drink Frequency 7) – 0.14723(Drinks/Day 12 – Opiates) + 0.22343(Drink Frequency 12 – Hallucinogens) – 0.066402(Total Tobacco 7 – Cocaine) – 0.099145(Total Tobacco 7 – Opiates) + 0.038182(Tobacco Frequency 7 – Cocaine) – 0.065418(Tobacco Frequency 7 – Opiates) – 0.15984(Tobacco Frequency 7 – Marijuana) – 0.6715(Tobacco Frequency – Mother) – 0.3597(Tobacco Frequency – Gender) – 0.032032(Cocaine – Hallucinogens) + 0.081362(Hallucinogens – Father) + 2.3972(Hallucinogens – Mother) + 0.23292(Opiates – Gender) + 0.0064412(Stimulants – Father) – 0.81659(Father – Mother) + 0.15155(Father – Gender) | 0.6265 | 6.35e-180 |
| marijuana ~ 8.3068 – 0.50742(VS – Tobacco Frequency 7) + 0.70711(VS – Stimulants) – 0.5338(Age – Total Tobacco 7) – 0.50275(Total Drinks 7 – Cocaine) – 1.0985(Drink Frequency 7 – Drinks/Day 2) – 1.0304(Drinks/Day 12 – Drink Frequency 12) + 0.38691(Drink Frequency 12 – Cocaine) + 0.82055(Drink Frequency 12 – Gender) – 0.49909(Total Tobacco 7 – Stimulants) – 0.80167(Opiates – Father) – 1.0505(Sedatives – Gender) | 0.4715 | 4.31e-120 |

**Supplementary Table 7:**
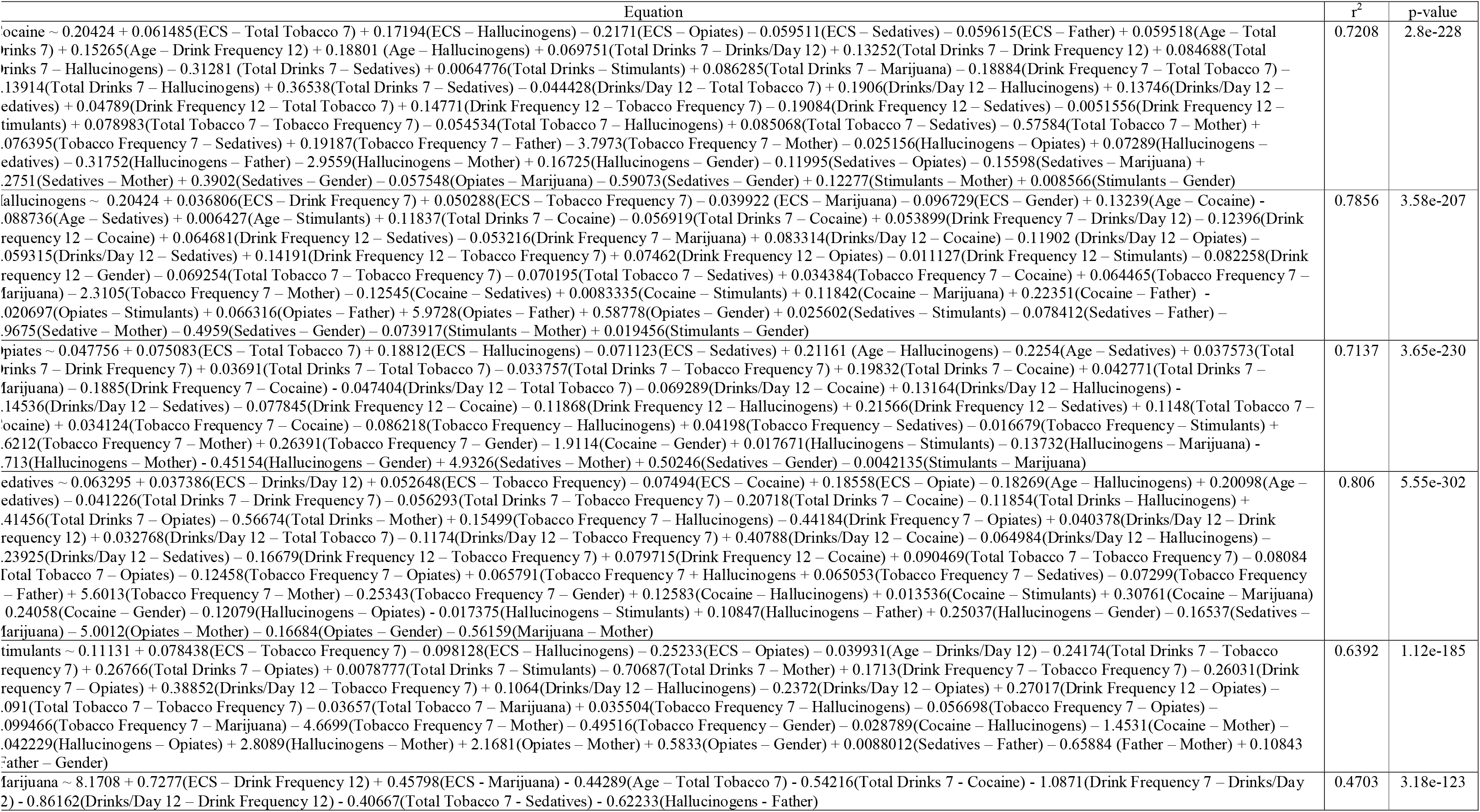
GLM equations and their corresponding r^2^- and p-values (Executive Control System). To enable comparison of effect sizes across relationships, all predictor and outcome variables were standardized to *Z* scores with a mean of 0 and an SD of 1. Regression coefficients are therefore representative of SD change in outcome per unit SD change in predictor and may be interpreted as partial correlation coefficients. “7” (e.g. Total Tobacco 7, Drink Frequency 7) indicates 7 days; “12” (e.g. Drinks/Day 12) indicates 12 months.

**Supplementary Table 8:** GLM equations and their corresponding r^2^- and p-values (Valuation-Control Complex). To enable comparison of effect sizes across relationships, all predictor and outcome variables were standardized to *Z* scores with a mean of 0 and an SD of 1. Regression coefficients are therefore representative of SD change in outcome per unit SD change in predictor and may be interpreted as partial correlation coefficients. “7” (e.g. Total Tobacco 7, Drink Frequency 7) indicates 7 days; “12” (e.g. Drinks/Day 12) indicates 12 months.

| Equation | $r^2$ | p-value |
| --- | --- | --- |
| Cocaine ~ 0.21801 + 0.070669(VCC – Total Tobacco 7) + 0.18494(VCC – Hallucinogens) – 0.19881(VCC – Opiates) – 0.048278(VCC – Sedatives) – 0.0050319(VCC – Stimulants) – 0.056556(VCC – Father) + 0.057609(Age – Total Drinks 7) + 0.14546(Age – Tobacco Frequency 7) + 0.1923(Age – Hallucinogens) + 0.074442(Total Drinks 7 – Drinks/Day 12) + 0.1322(Total Drinks 7 – Tobacco frequency 7) + 0.091828(Total Drinks – Hallucinogens) – 0.31607(Total Drinks 7 – Sedatives) + 0.0084922(Total Drinks 7 – Stimulants) + 0.09164(Total Drinks 7 – Marijuana) – 0.19438(Drink frequency 7 – Tobacco Frequency 7) – 0.15694(Tobacco Frequency 7 + Hallucinogens) + 0.38012(Tobacco Frequency 7 – Sedatives) – 0.054669(Drinks/Day 12 – Total Tobacco 7) + .1659(Drinks/Day 12 – Hallucinogens) + 0.12783(Drinks/Day 12 – Sedatives) + 0.14028(Drink Frequency 12 – Tobacco Frequency 7) – 0.19896(Drink Frequency 12 – Sedatives) + 0.090095(Total Tobacco 7 – Tobacco Frequency 7) – 0.084401(Total Tobacco 7 – Hallucinogens) + 0.088186(Total Tobacco 7 – Sedatives) + 0.066431(Tobacco Frequency 7 – Sedatives) + 0.20564(Tobacco Frequency – Father) – 3.4857(Tobacco Frequency 7 – Mother) + 0.061953(Hallucinogens – Sedatives) – 0.31718(Hallucinogens – Father) – 3.3182(Hallucinogens – Mother) + 0.21742(Hallucinogens – Gender) – .11906(Opiates – Sedatives) – 0.1508(Opiates – Marijuana) + 2.1964(Opiates – Mother) + 0.27352(Opiates – Mother) + 0.27352(Opiates – Gender) – 0.067314(Opiates – Marijuana) – 0.611(Opiates – Gender) + 0.11112(Stimulants – Mother) + 0.012633(Stimulants – Gender) | 0.7198 | 1.95e-229 |
| Hallucinogens ~ -0.2302 – 0.048987(VCC – Total Tobacco 7) + 0.18501(VCC – Opiates) + 0.054542(Age – Drinks/Day 12) + 0.18734(Age – Cocaine) + 0.047491(Total Drinks 7 – Drink Frequency 7) – 0.13593(Total Drinks 7 + Cocaine) – 0.045186(Total Drinks 7 – Opiates) – 0.058856(Total Drinks 7 – Marijuana) – 0.058856(Total Drinks 7 – Marijuana) – 0.1669(Drink Frequency 7 – Cocaine) + .060532(Drink Frequency 7 – Opiates) – 0.12282(Drinks/Day 12 – Opiates) + 0.1384(Drink Frequency 12 – Tobacco Frequency 7) – 0.010106(Drink Frequency 12 – Stimulants) – 0.056535(Total Tobacco 7 – Opiates) – 0.073854(Total Tobacco 7 – Sedatives) – 0.10552(Cocaine – Sedatives) + 0.11938(Cocaine – Marijuana) + 0.26035(Cocaine – Father) + 0.15462(Cocaine – Gender) + .017296(Sedatives – Stimulants) – 0.044036(Sedatives – Father) + 0.016868(Stimulants – Gender) | 0.6612 | 5.64e-204 |
| Opiates ~ -0.13254 – 0.045838(VCC – Drinks/Day 12) + 0.096573(VCC – Total Tobacco 7) + 0.08931(VCC – Hallucinogens) 0.063258(Age – Cocaine) – 0.21705(Age – Sedatives) + 0.037971(Total Drinks 7 – Drink Frequency 7) + 0.18642(Total Drinks 7 – Cocaine) – 0.17706(Drink Frequency 7 – Cocaine) – 0.0482(Drinks/Day 12 – Total Tobacco 7) – 0.11275(Drinks/Day 12 – Opiate) – .056763(Drink Frequency 12 – Cocaine) – 0.18749(Drink Frequency 12 – Hallucinogens) + 0.20665(Drink Frequency 12 – Sedatives) + 0.10117(Total Tobacco 7 – Cocaine) – 0.056695(Tobacco frequency 7 – Hallucinogens) + 0.047575(Tobacco Frequency 7 – Sedatives) + 0.1865(Tobacco Frequency 7 – Gender) – 2.2217(Cocaine – Mother) – 0.20784(Cocaine – Gender) – .094492(Hallucinogens – Marijuana) – 0.21276(Hallucinogens – Gender) + 3.3288(Sedatives – Mother) + 0.38016(Sedatives – Gender) | 0.6996 | 3.94e-227 |
| Sedatives ~ 0.024389 + 0.039047(VCC – Drinks/Day 12) + 0.082159(VCC – Tobacco Frequency 7) – 0.13514(VCC – Opiates) + 0.18292(VCC – Sedatives) – 0.16846(Age – Cocaine) + 0.18293(Age – Sedatives) – 0.042658(Total Drinks 7 – Drink Frequency 7) – 0.05001(Total Drinks 7 – Tobacco Frequency 7) – 0.1456(Total Drinks 7 – Cocaine) – 0.20886(Total Drinks 7 – Hallucinogens) + .43029(Total Drinks 7 – Opiate) – 0.68629(Total Drinks 7 – Mother) + 0.18042(Drink Frequency 7 – Cocaine) – 0.46683(Drink Frequency 7 – Opiates) + 0.041472(Drinks/Day 12 – Drink Frequency 2) + 0.031309(Drinks/Day 12 – Total Tobacco 7) – 0.10872(Drinks/Day 12 – Tobacco Frequency 7) – 0.062916 (Drinks/Day 12 – Cocaine) + 0.38104(Drinks/Day 12 – Hallucinogens) – 0.24011(Drink frequency 12 – Tobacco Frequency 7) + 0.07902(Drink Frequency 12 – Hallucinogens) + 0.085114(Total Tobacco 7 – Tobacco Frequency 7) – 0.087971 (Total Tobacco 7 – Opiates) + 0.06194(Tobacco frequency 7 – Cocaine) – 0.12195(Tobacco Frequency 7 – Hallucinogens) + 0.069098(Tobacco Frequency 7 – Sedatives) – 0.069737(Tobacco Frequency 7 – Father) + 5.4962(Tobacco Frequency 7 – Mother) – 0.2812(Tobacco Frequency – Gender) + 0.11883(Cocaine – Hallucinogens) – 0.11238(Cocaine – Opiates) – 0.014053(Cocaine – Stimulants) + 0.09909(Cocaine – Father) + 0.24428(Cocaine – Gender) + 0.015054(Hallucinogens – Stimulants) + 0.30791(Hallucinogens – Marijuana) + 1.3966(Hallucinogens – Mother) + 0.35179(Hallucinogens – Gender) – 0.0058883(Opiates – Stimulants) – .16606(Opiates – Marijuana) – 5.6098(Opiates – Mother) – 0.11617(Opiates – Gender) – 0.56871(Marijuana – Mother) | 0.8083 | 5.7e-303 |
| Stimulants ~ 0.12215 + 0.062193(VCC – Tobacco Frequency 7) – 0.089502(VCC – Opiates) – 0.16196(VCC – Sedatives) – 0.22405(Total Drinks 7 – Tobacco Frequency 7) + 0.25054(Total Drinks 7 – Opiates) + 0.0077219(Total Drinks 7 – Sedatives) – 1.0664(Total Drinks 7 – Mother) + 0.15457(Drink Frequency 7 – Tobacco Frequency 7) – 0.25852(Tobacco Frequency – Sedatives) + .33811(Drinks/Day 12 – Tobacco Frequency 7) – 0.13356(Drinks/Day 12 – Opiates) + 0.31323(Drink Frequency 12 – Hallucinogens) – 0.074914(Drink Frequency 12 – Hallucinogens) – 0.07229 (Total Tobacco 7 – Tobacco Frequency 7) – 0.044806(Total Tobacco 7 – Cocaine) – 0.03702(Total Tobacco 7 – Marijuana) – 0.048082(Tobacco Frequency 7 – Opiates) + 0.013621(Tobacco Frequency 7 – Sedatives) – 0.12984(Tobacco Frequency 7 – Marijuana) – 3.9026(Tobacco Frequency 7 – Mother) – 0.55796(Tobacco Frequency 7 – Gender) + 3.2924(Hallucinogens – Mother) – 0.016408(Opiates – Sedatives) + 0.45508(Opiates – Gender) + 0.0076174(Stimulants – Father) – 0.57193(Father – Mother) + 0.11931(Father – Gender) | 0.6288 | 252e-182 |
| Marijuana ~ 8.2017 + 0.69869(VCC – Drink Frequency 12) – 0.78946(VCC – Opiates) + 0.62535(VCC – Marijuana) – 0.62702(Age – Opiates) – 0.55758(Total Drinks 7 – Cocaine) – 1.0602(Drink frequency 7 – Drinks/Day 12) – 0.84237(Drinks/Day 12 – Drink Frequency 12) – 0.37699(Total Tobacco 7 – Sedatives) – 0.82425(Hallucinogens – Father) | 0.4732 | 4.32e-123 |

**Supplementary Table 9:**
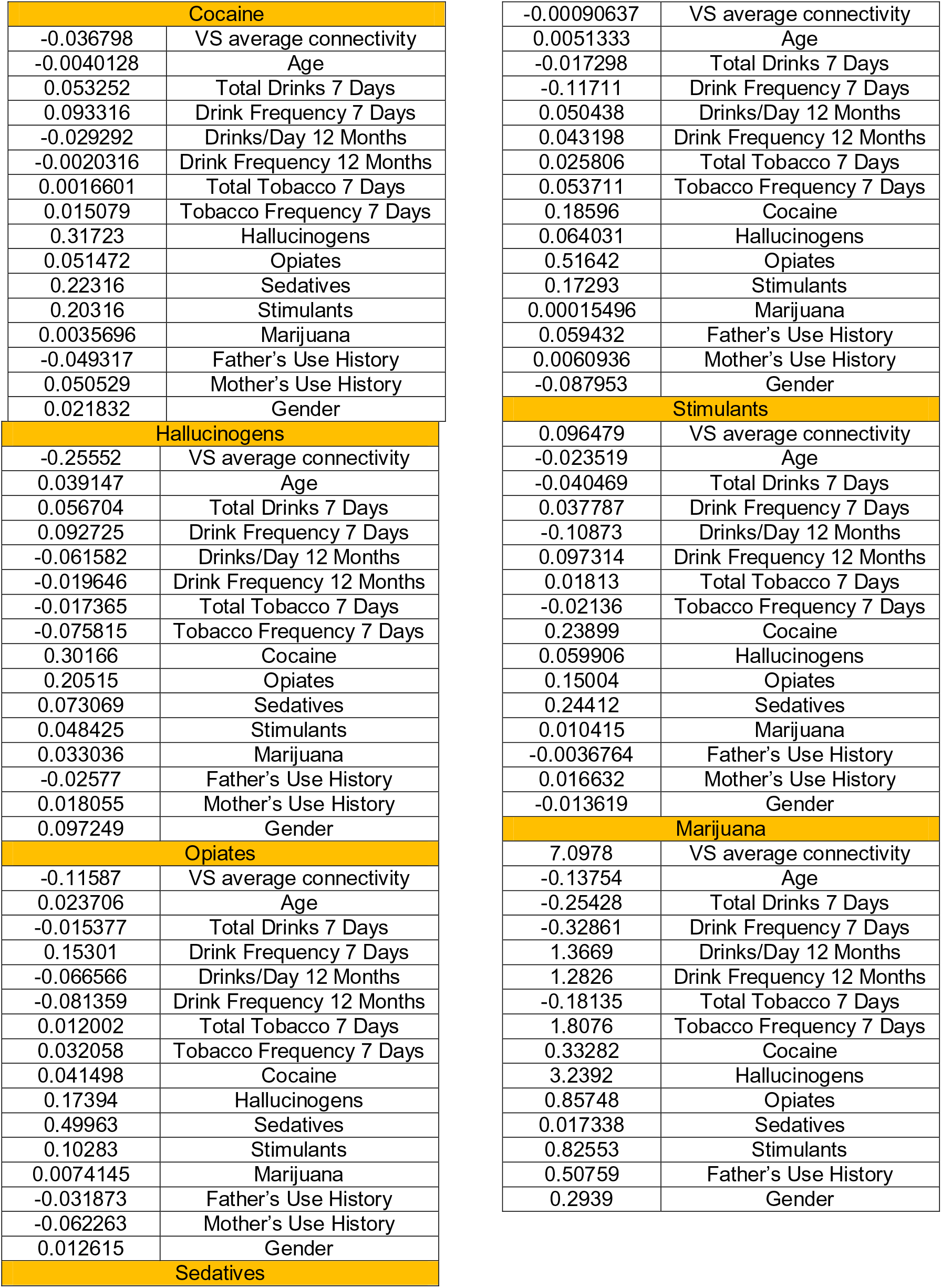
Beta coefficients for all dependent variables used in the Valuation System (VS) GLM analysis.

**Supplementary Table 10:**
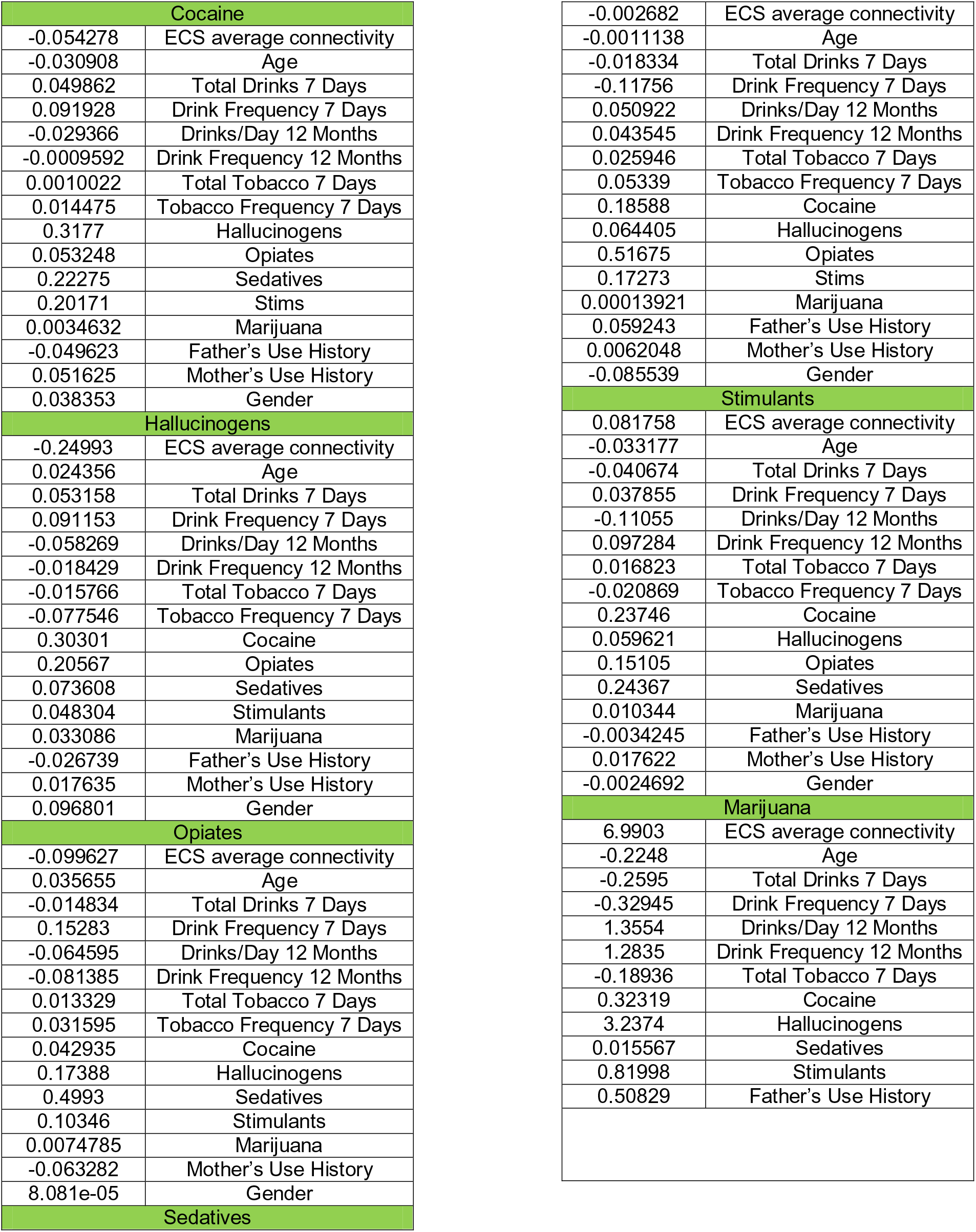
Beta coefficients for all dependent variables used in the Executive Control System (ECS) GLM analysis.

**Supplementary Table 11:** Beta coefficients for all dependent variables used in the Valuation-Control Complex (VCC) GLM analysis.

| Cocaine |  |
| --- | --- |
| -0.049962 | VCC Average Connectivity |
| -0.025465 | Age |
| 0.050458 | Total Drinks 7 Days |
| 0.091885 | Drink Frequency 7 Days |
| -0.028744 | Drinks/Day 12 Months |
| -0.0011179 | Drink Frequency 12 Months |
| 0.0014423 | Total Tobacco 7 Days |
| 0.014491 | Tobacco Frequency 7 Days |
| 0.3181 | Hallucinogens |
| 0.052813 | Opiates |
| 0.22304 | Sedatives |
| 0.20204 | Stimulants |
| 0.0034975 | Marijuana |
| -0.049828 | Father's Use History |
| 0.051446 | Mother's Use History |
| 0.033915 | Gender |
| Hallucinogens |  |
| -0.24443 | VCC Average Connectivity |
| 0.035679 | Age |
| 0.054761 | Total Drinks 7 Days |
| 0.092129 | Drink Frequency 7 Days |
| -0.059251 | Drinks/Day 12 Months |
| -0.018987 | Drink Frequency 12 Months |
| -0.016038 | Total Tobacco 7 Days |
| -0.076975 | Tobacco Frequency 7 Days |
| 0.30293 | Cocaine |
| 0.20484 | Opiates |
| 0.0733 | Sedatives |
| 0.048699 | Stimulants |
| 0.033072 | Marijuana |
| -0.02614 | Father's Use History |
| 0.091916 | Gender |
| Opiates |  |
| -0.10261 | VCC Average Connectivity |
| 0.033092 | Age |
| -0.014998 | Total Drinks 7 Days |
| 0.15322 | Drink Frequency 7 Days |
| -0.065477 | Drinks/Day 12 Months |
| -0.081394 | Drink Frequency 12 Months |
| 0.01284 | Total Tobacco 7 Days |
| 0.03172 | Tobacco Frequency 7 Days |
| 0.042579 | Cocaine |
| 0.17342 | Hallucinogens |
| 0.49921 | Sedatives |
| 0.10336 | Stimulants |
| 0.0074543 | Marijuana |
| -0.063216 | Mother's Use History |
| 0.0032496 | Gender |
| Sedatives |  |

| -0.00047632 | VCC Average Connectivity |
| --- | --- |
| 0.0027925 | Age |
| -0.017812 | Total Drinks 7 Days |
| -0.11732 | Drink Frequency 7 Days |
| 0.050815 | Drinks/Day 12 Months |
| 0.043372 | Drink Frequency 12 Months |
| 0.025975 | Total Tobacco 7 Days |
| 0.053506 | Tobacco Frequency 7 Days |
| 0.18604 | Hallucinogens |
| 0.064204 | Opiates |
| 0.5165 | Sedatives |
| 0.17289 | Stimulants |
| 0.00015173 | Marijuana |
| 0.059336 | Father's Use History |
| 0.0060493 | Mother's Use History |
| -0.087633 | Gender |
| Stimulants |  |
| 0.083756 | VCC Average Connectivity |
| -0.040712 | Total Drinks 7 Days |
| 0.037457 | Drink Frequency 7 Days |
| -0.10971 | Drinks/Day 12 Months |
| 0.097337 | Drink Frequency 12 Months |
| 0.017278 | Total Tobacco 7 Days |
| -0.021013 | Tobacco Frequency 7 Days |
| 0.23776 | Hallucinogens |
| 0.060179 | Opiates |
| 0.15087 | Sedatives |
| 0.24392 | Stimulants |
| 0.010365 | Marijuana |
| -0.0037528 | Father's Use History |
| 0.017598 | Mother's Use History |
| -0.0046949 | Gender |
| Marijuana |  |
| 7.021 | VCC Average Connectivity |
| -0.18956 | Age |
| -0.25597 | Total Drinks 7 Days |
| -0.33041 | Drink Frequency 7 Days |
| 1.3605 | Drinks/Day 12 Months |
| 1.2828 | Drink Frequency 12 Months |
| -0.18623 | Total Tobacco 7 Days |
| 0.32632 | Cocaine |
| 3.2403 | Hallucinogens |
| 0.86268 | Opiates |
| 0.016972 | Sedatives |
| 0.82179 | Stimulants |
| 0.50696 | Father's Use History |

### GLM Outputs

This section contains GLM outputs for all models analyzed in this study. The data shown here can be analyzed in conjunction with data from Table 4 in the main text as well as Supplementary Tables 6-11. FDR, False Discovery Rate

### Valuation System

#### Cocaine

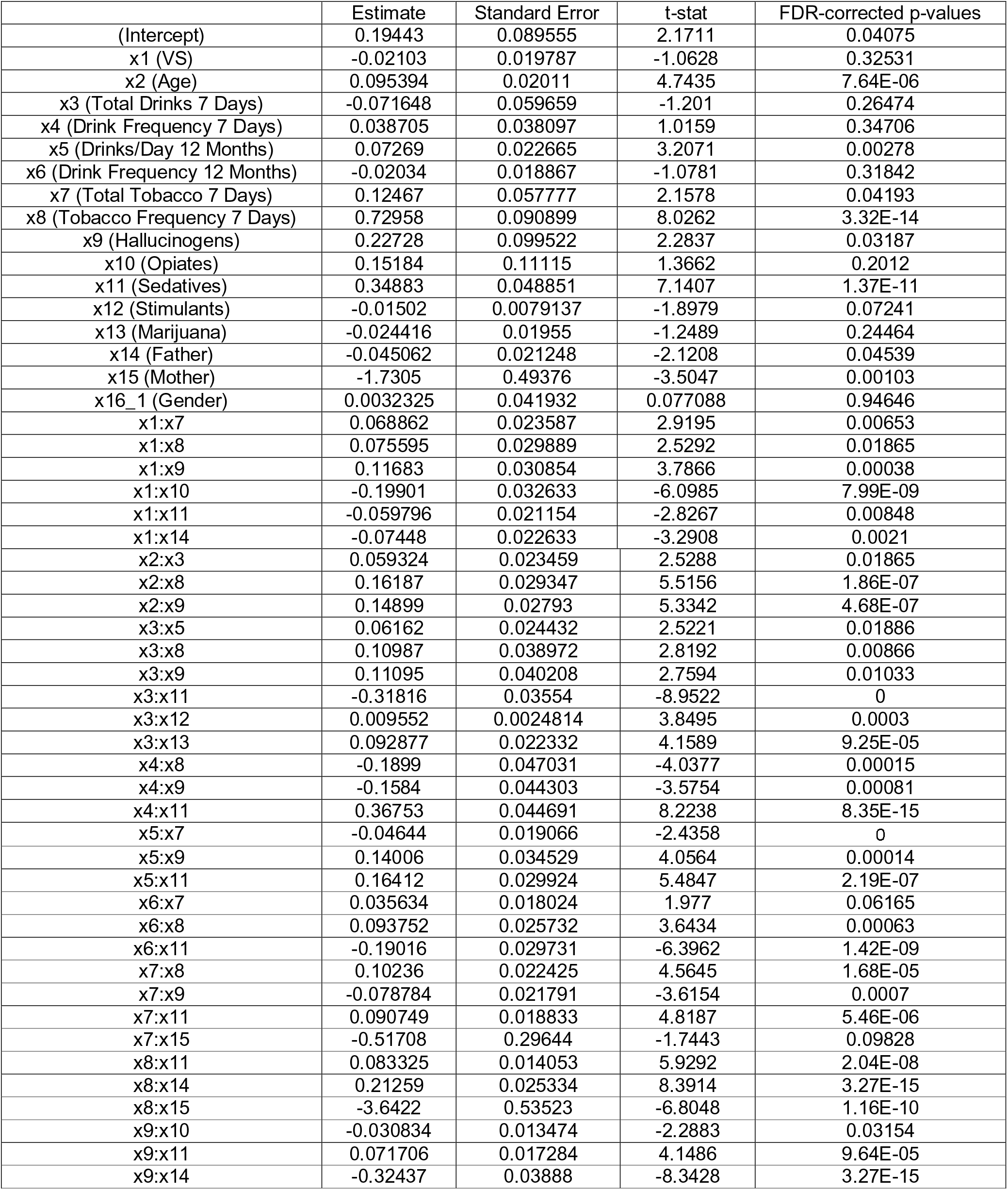
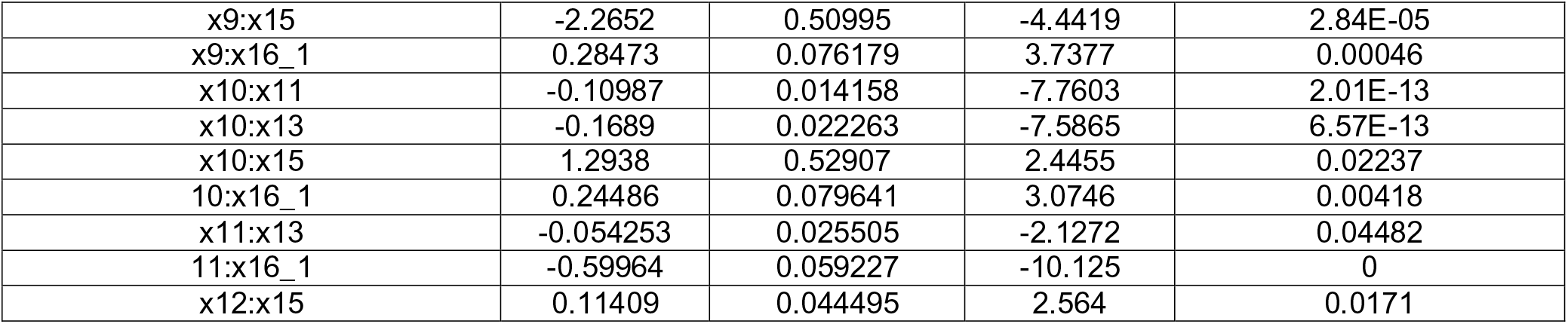

|  | Estimate | Standard Error | t-stat | FDR-corrected p-values |
| --- | --- | --- | --- | --- |
| (Intercept) | 0.19443 | 0.089555 | 2.1711 | 0.04075 |
| x1 (VS) | -0.02103 | 0.019787 | -1.0628 | 0.32531 |
| x2 (Age) | 0.095394 | 0.02011 | 4.7435 | 7.64E-06 |
| x3 (Total Drinks 7 Days) | -0.071648 | 0.059659 | -1.201 | 0.26474 |
| x4 (Drink Frequency 7 Days) | 0.038705 | 0.038097 | 1.0159 | 0.34706 |
| x5 (Drinks/Day 12 Months) | 0.07269 | 0.022665 | 3.2071 | 0.00278 |
| x6 (Drink Frequency 12 Months) | -0.02034 | 0.018867 | -1.0781 | 0.31842 |
| x7 (Total Tobacco 7 Days) | 0.12467 | 0.057777 | 2.1578 | 0.04193 |
| x8 (Tobacco Frequency 7 Days) | 0.72958 | 0.090899 | 8.0262 | 3.32E-14 |
| x9 (Hallucinogens) | 0.22728 | 0.099522 | 2.2837 | 0.03187 |
| x10 (Opiates) | 0.15184 | 0.11115 | 1.3662 | 0.2012 |
| x11 (Sedatives) | 0.34883 | 0.048851 | 7.1407 | 1.37E-11 |
| x12 (Stimulants) | -0.01502 | 0.0079137 | -1.8979 | 0.07241 |
| x13 (Marijuana) | -0.024416 | 0.01955 | -1.2489 | 0.24464 |
| x14 (Father) | -0.045062 | 0.021248 | -2.1208 | 0.04539 |
| x15 (Mother) | -1.7305 | 0.49376 | -3.5047 | 0.00103 |
| x16_1 (Gender) | 0.0032325 | 0.041932 | 0.077088 | 0.94646 |
| x1:x7 | 0.068862 | 0.023587 | 2.9195 | 0.00653 |
| x1:x8 | 0.075595 | 0.029889 | 2.5292 | 0.01865 |
| x1:x9 | 0.11683 | 0.030854 | 3.7866 | 0.00038 |
| x1:x10 | -0.19901 | 0.032633 | -6.0985 | 7.99E-09 |
| x1:x11 | -0.059796 | 0.021154 | -2.8267 | 0.00848 |
| x1:x14 | -0.07448 | 0.022633 | -3.2908 | 0.0021 |
| x2:x3 | 0.059324 | 0.023459 | 2.5288 | 0.01865 |
| x2:x8 | 0.16187 | 0.029347 | 5.5156 | 1.86E-07 |
| x2:x9 | 0.14899 | 0.02793 | 5.3342 | 4.68E-07 |
| x3:x5 | 0.06162 | 0.024432 | 2.5221 | 0.01886 |
| x3:x8 | 0.10987 | 0.038972 | 2.8192 | 0.00866 |
| x3:x9 | 0.11095 | 0.040208 | 2.7594 | 0.01033 |
| x3:x11 | -0.31816 | 0.03554 | -8.9522 | 0 |
| x3:x12 | 0.009552 | 0.0024814 | 3.8495 | 0.0003 |
| x3:x13 | 0.092877 | 0.022332 | 4.1589 | 9.25E-05 |
| x4:x8 | -0.1899 | 0.047031 | -4.0377 | 0.00015 |
| x4:x9 | -0.1584 | 0.044303 | -3.5754 | 0.00081 |
| x4:x11 | 0.36753 | 0.044691 | 8.2238 | 8.35E-15 |
| x5:x7 | -0.04644 | 0.019066 | -2.4358 | 0 |
| x5:x9 | 0.14006 | 0.034529 | 4.0564 | 0.00014 |
| x5:x11 | 0.16412 | 0.029924 | 5.4847 | 2.19E-07 |
| x6:x7 | 0.035634 | 0.018024 | 1.977 | 0.06165 |
| x6:x8 | 0.093752 | 0.025732 | 3.6434 | 0.00063 |
| x6:x11 | -0.19016 | 0.029731 | -6.3962 | 1.42E-09 |
| x7:x8 | 0.10236 | 0.022425 | 4.5645 | 1.68E-05 |
| x7:x9 | -0.078784 | 0.021791 | -3.6154 | 0.0007 |
| x7:x11 | 0.090749 | 0.018833 | 4.8187 | 5.46E-06 |
| x7:x15 | -0.51708 | 0.29644 | -1.7443 | 0.09828 |
| x8:x11 | 0.083325 | 0.014053 | 5.9292 | 2.04E-08 |
| x8:x14 | 0.21259 | 0.025334 | 8.3914 | 3.27E-15 |
| x8:x15 | -3.6422 | 0.53523 | -6.8048 | 1.16E-10 |
| x9:x10 | -0.030834 | 0.013474 | -2.2883 | 0.03154 |
| x9:x11 | 0.071706 | 0.017284 | 4.1486 | 9.64E-05 |
| x9:x14 | -0.32437 | 0.03888 | -8.3428 | 3.27E-15 |
| x9:x15 | -2.2652 | 0.50995 | -4.4419 | 2.84E-05 |
| x9:x16_1 | 0.28473 | 0.076179 | 3.7377 | 0.00046 |
| x10:x11 | -0.10987 | 0.014158 | -7.7603 | 2.01E-13 |
| x10:x13 | -0.1689 | 0.022263 | -7.5865 | 6.57E-13 |
| x10:x15 | 1.2938 | 0.52907 | 2.4455 | 0.02237 |
| 10:x16_1 | 0.24486 | 0.079641 | 3.0746 | 0.00418 |
| x11:x13 | -0.054253 | 0.025505 | -2.1272 | 0.04482 |
| 11:x16_1 | -0.59964 | 0.059227 | -10.125 | 0 |
| x12:x15 | 0.11409 | 0.044495 | 2.564 | 0.0171 |

#### Hallucinogens

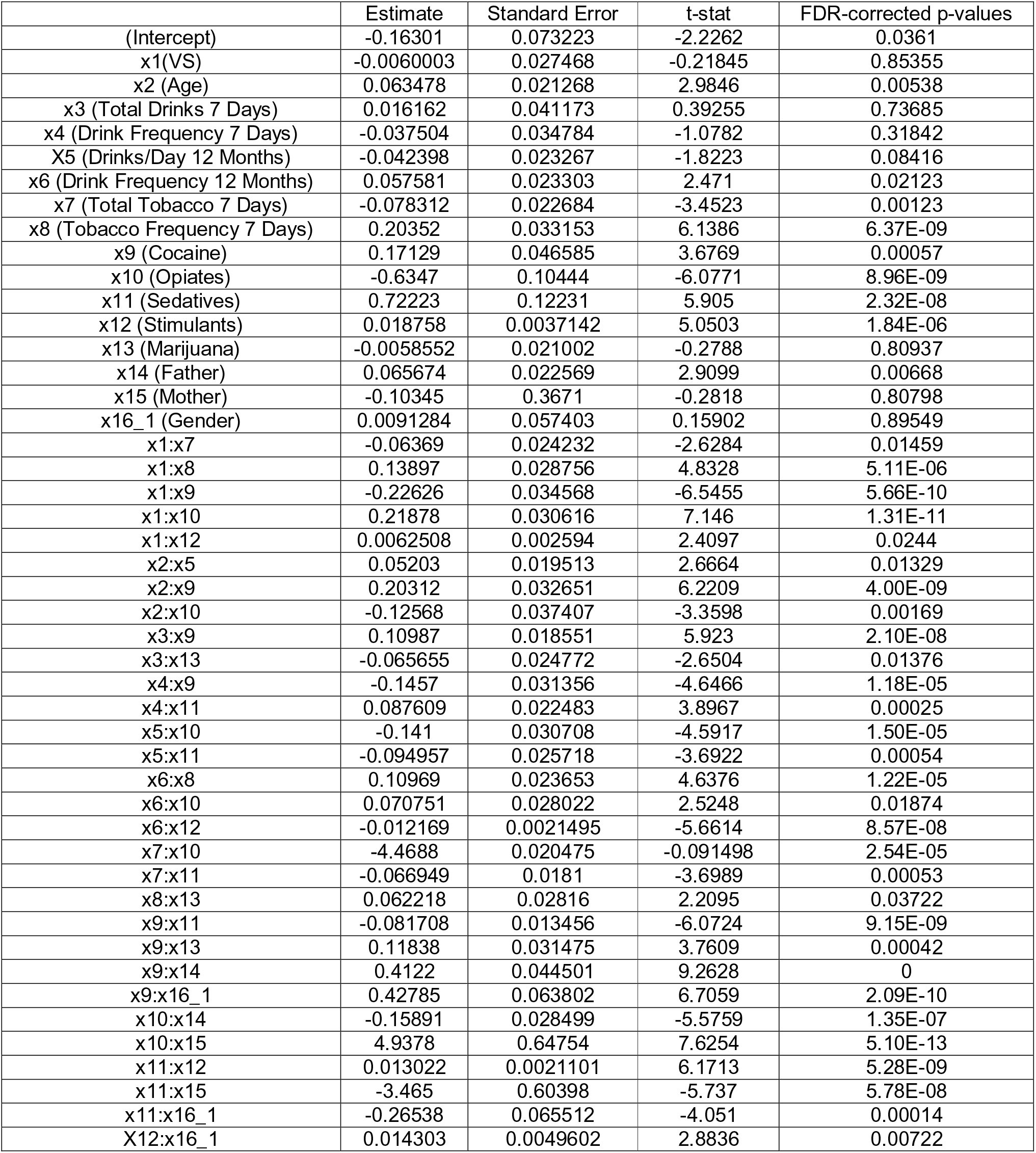

#### Opiates

|  | Estimate | Standard Error | t-stat | FDR-corrected p-values |
| --- | --- | --- | --- | --- |
| (Intercept) | -0.077541 | 0.070267 | -1.1035 | 0.30701 |
| x1 (VS) | 0.040826 | 0.020083 | 2.0329 | 0.05481 |
| x2 (Age) | -0.038028 | 0.020823 | -1.8263 | 0.08354 |
| x3 (Total Drinks 7 Days) | 0.026743 | 0.040899 | 0.65387 | 0.55642 |
| x4 (Drink Frequency 7 Days) | -0.010397 | 0.033645 | -0.30903 | 0.79001 |
| x5 (Drinks/Day 12 Months) | -0.065305 | 0.022145 | -2.9489 | 0.00599 |
| x6 (Drink Frequency 7 Days) | -0.0084335 | 0.018997 | -0.44393 | 0.69878 |
| x7 (Total Tobacco 7 Days) | 0.041498 | 0.022255 | 1.8647 | 0.07748 |
| x8 (Tobacco Frequency 7 Days) | 0.062363 | 0.033979 | 1.8354 | 0.08211 |
| x9 (Cocaine) | 0.53118 | 0.097458 | 5.4503 | 2.59E-07 |
| x10 (Hallucinogens) | 0.41164 | 0.050822 | 8.0995 | 2.00E-14 |
| x11 (Sedatives) | -0.52993 | 0.09063 | -5.8472 | 3.18E-08 |
| x12 (Stimulants) | 0.0087431 | 0.0026062 | 3.3547 | 0.00172 |
| x13 (Marijuana) | -0.046655 | 0.019967 | -2.3367 | 0.02829 |
| x14 (Father) | -0.016033 | 0.020014 | -0.80108 | 0.46662 |
| x15 (Mother) | -0.013956 | 0.35628 | -0.039172 | 0.97223 |
| x16_1 (Gender) | -0.00019884 | 0.04351 | -0.00457 | 0.99635 |
| x1:x5 | -0.047223 | 0.02008 | -2.3517 | 0.0275 |
| x1:x7 | 0.07354 | 0.022599 | 3.2541 | 0.00237 |
| x1:x8 | 0.11462 | 0.019541 | 5.8657 | 2.90E-08 |
| x1:x14 | -0.036665 | 0.021236 | -1.7265 | 0.10179 |
| x2:x10 | 0.065478 | 0.032658 | 2.005 | 0.05848 |
| x2:x11 | -0.17539 | 0.032013 | -5.4786 | 2.24E-07 |
| x3:x7 | 0.042794 | 0.018475 | 2.3163 | 0.02965 |
| x3:x9 | 0.19854 | 0.022039 | 9.0082 | 0 |
| x4:x9 | -0.19573 | 0.033408 | -5.8589 | 2.98E-08 |
| x4:x10 | 0.065957 | 0.032399 | 2.0358 | 0.05451 |
| x5:x7 | -0.061074 | 0.020429 | -2.9896 | 0.0053 |
| x5:x9 | -0.063834 | 0.028204 | -2.2633 | 0.03322 |
| x5:x10 | 0.083089 | 0.02691 | 3.0876 | 0.00404 |
| x5:x11 | -0.14065 | 0.026189 | -5.3707 | 3.86E-07 |
| x6:x9 | -0.071936 | 0.024398 | -2.9484 | 0.00599 |
| x6:x10 | -0.12058 | 0.031964 | -3.7725 | 0.0004 |
| x6:x11 | 0.16286 | 0.024471 | 6.6553 | 2.89E-10 |
| x7:x8 | -0.047018 | 0.018819 | -2.4985 | 0.02 |
| x7:x9 | 0.14016 | 0.023093 | 6.0696 | 9.25E-09 |
| x8:x10 | -0.082762 | 0.011907 | -6.9506 | 4.69E-11 |
| x8:x11 | 0.04842 | 0.010479 | 4.6206 | 1.32E-05 |
| x9:x15 | -2.2096 | 0.4736 | -4.6655 | 1.09E-05 |
| x9:x16_1 | -0.19427 | 0.063385 | -3.0649 | 0.00428 |
| x10:x12 | 0.0052223 | 0.0025444 | 2.0524 | 0.0527 |
| x10:x13 | -0.13244 | 0.020595 | -6.4307 | 1.15E-09 |
| x11:x15 | 3.216 | 0.41204 | 7.8051 | 1.48E-13 |
| x11:x16_1 | 0.31369 | 0.059312 | 5.2888 | 5.84E-07 |

#### Sedatives

|  | Estimate | Standard Error | t-stat | FDR-corrected p-values |
| --- | --- | --- | --- | --- |
| (Intercept) | 0.022372 | 0.062735 | 0.35661 | 0.75943 |
| x1 (VS) | 0.04005 | 0.01716 | 2.3339 | 0.02845 |
| x2 (Age) | 0.0014991 | 0.017759 | 0.084415 | 0.94466 |
| x3 (Total Drinks 7 Days) | 0.20616 | 0.074978 | 2.7496 | 0.0106 |
| x4 (Drink Frequency 7 Days) | -0.059464 | 0.029751 | -1.9987 | 0.05903 |
| x5 (Drinks/Day 12 Months) | 0.00073822 | 0.019948 | 0.037007 | 0.97281 |
| x6 (Drink Frequency 12 Months) | 0.0039006 | 0.018343 | 0.21265 | 0.85716 |
| x7 (Total Tobacco 7 Days) | 0.022957 | 0.019201 | 1.1956 | 0.26677 |
| x8 (Tobacco Frequency 7 Days) | -0.65301 | 0.10563 | -6.1818 | 5.03E-09 |
| x9 (Cocaine) | 0.20559 | 0.082675 | 2.4867 | 0.02052 |
| x10 (Hallucinogens) | 0.10711 | 0.056903 | 1.8824 | 0.07478 |
| x11 (Opiates) | 0.94797 | 0.093351 | 10.155 | 0 |
| x12 (Stimulants) | -0.0012394 | 0.0022654 | -0.54707 | 0.62541 |
| x13 (Marijuana) | 0.17369 | 0.044953 | 3.8638 | 0.00029 |
| x14 (Father) | 0.0062953 | 0.018357 | 0.34294 | 0.76735 |
| x15 (Mother) | 0.29914 | 0.31024 | 0.96421 | 0.37293 |
| x16_1 (Gender) | 0.0035812 | 0.037342 | 0.095903 | 0.93814 |
| x1:x5 | 0.044131 | 0.017117 | 2.5782 | 0.01658 |
| x1:x8 | 0.05633 | 0.027295 | 2.0637 | 0.05137 |
| x1:x9 | 0.087239 | 0.027482 | 3.1744 | 0.00309 |
| x1:x10 | -0.11234 | 0.025444 | -4.415 | 3.20E-05 |
| x1:x11 | 0.1094 | 0.01925 | 5.6831 | 7.78E-08 |
| x2:x4 | 0.035095 | 0.01753 | 2.0019 | 0.05882 |
| x2:x9 | -0.13456 | 0.024858 | -5.4131 | 3.14E-07 |
| x2:x11 | 0.18204 | 0.030242 | 6.0193 | 1.21E-08 |
| x3:x4 | -0.038749 | 0.018143 | -2.1358 | 0.04415 |
| x3:x10 | -0.29372 | 0.024593 | -11.943 | 0 |
| x3:x11 | 0.37067 | 0.032158 | 11.527 | 0 |
| x3:x15 | -0.95341 | 0.31225 | -3.0533 | 0.0044 |
| x4:x8 | -0.062641 | 0.025853 | -2.423 | 0.02363 |
| x4:x10 | 0.089163 | 0.029154 | 3.0583 | 0.00437 |
| x4:x11 | -0.40739 | 0.04155 | -9.8047 | 0 |
| x5:x6 | 0.034425 | 0.01492 | 2.3074 | 0.03026 |
| x5:x7 | 0.040748 | 0.016035 | 2.5413 | 0.01814 |
| x5:x8 | -0.1544 | 0.025616 | -6.0276 | 1.16E-08 |
| x5:x9 | -0.095024 | 0.025905 | -3.6681 | 0.00058 |
| x5:x10 | 0.41269 | 0.029598 | 13.943 | 0 |
| x5:x11 | -0.21047 | 0.027074 | -7.7739 | 1.84E-13 |
| x6:x8 | -0.16845 | 0.025648 | -6.5679 | 4.98E-10 |
| x7:x8 | 0.1052 | 0.017689 | 5.9472 | 1.85E-08 |
| x7:x11 | -0.078702 | 0.017486 | -4.5008 | 2.20E-05 |
| x7:x14 | -0.030747 | 0.012369 | -2.4859 | 0.02053 |
| x8:x9 | 0.050822 | 0.012278 | 4.1392 | 0.0001 |
| x8:x10 | -0.12095 | 0.013251 | -9.1279 | 0 |
| x8:x11 | 0.058513 | 0.012748 | 4.59 | 1.51E-05 |
| x8:x15 | 5.4137 | 0.50968 | 10.622 | 0 |
| x8:x16_1 | -0.24899 | 0.059585 | -4.1788 | 8.56E-05 |
| x9:x10 | 0.10356 | 0.01592 | 6.5047 | 7.31E-10 |
| x9:x11 | -0.097371 | 0.018421 | -5.2859 | 5.90E-07 |
| x9:x12 | -0.012427 | 0.0032082 | -3.8735 | 0.00028 |
| x9:x16_1 | 0.1893 | 0.060695 | 3.1188 | 0.00367 |
| x10:x11 | 0.031576 | 0.011918 | 2.6494 | 0.01377 |
| x10:x12 | 0.01372 | 0.002781 | 4.9333 | 3.18E-06 |
| x10:x13 | 0.26187 | 0.025894 | 10.113 | 0 |
| x10:x16_1 | 0.25412 | 0.054138 | 4.694 | 9.55E-06 |
| x11:x12 | -0.0076042 | 0.0033078 | -2.2989 | 0.03084 |
| x11:x13 | -0.12105 | 0.023165 | -5.2257 | 7.85E-07 |
| x11:x15 | -3.9793 | 0.48393 | -8.2228 | 8.35E-15 |
| x13:x15 | -0.58812 | 0.23422 | -2.511 | 0.01935 |

#### Stimulants

|  | Estimate | Standard Error | t-stat | FDR-corrected p-values |
| --- | --- | --- | --- | --- |
| x1 (VS) | -0.015797 | 0.02277 | -0.69377 | 0.53072 |
| x2 (Age) | -0.0021483 | 0.023112 | -0.092952 | 0.93938 |
| x3 (Total Drinks 7 Days) | 0.04537 | 0.09465 | 0.47934 | 0.6735 |
| x4 (Drink Frequency 7 Days) | -0.049917 | 0.044039 | -1.1335 | 0.29366 |
| x5 (Drinks/Day 12 Months) | 0.12911 | 0.02559 | 5.0453 | 1.88E-06 |
| x6 (Drink Frequency 12 Months) | 0.046473 | 0.021799 | 2.1318 | 0.04444 |
| x7 (Total Tobacco 7 Days) | -0.03839 | 0.024451 | -1.5701 | 0.13849 |
| x8 (Tobacco Frequency 7 Days) | 1.014 | 0.10643 | 9.5275 | 0 |
| x9 (Cocaine) | 0.088885 | 0.034089 | 2.6074 | 0.01532 |
| x10 (Hallucinogens) | -0.11112 | 0.086958 | -1.2779 | 0.23327 |
| x11 (Opiates) | 0.30046 | 0.058197 | 5.1627 | 1.07E-06 |
| x12 (Sedatives) | 0.0076386 | 0.0030082 | 2.5393 | 0.0182 |
| x13 (Marijuana) | -0.02888 | 0.022981 | -1.2567 | 0.24168 |
| x14 (Father) | 0.064528 | 0.063226 | 1.0206 | 0.3451 |
| x15 (Mother) | -0.6984 | 0.4068 | -1.7168 | 0.10376 |
| x16_1 (Gender) | 0.0095395 | 0.049015 | 0.19463 | 0.8706 |
| x1:x6 | -0.050192 | 0.020432 | -2.4566 | 0.02193 |
| x1:x11 | -0.0724 | 0.024844 | -2.9142 | 0.00662 |
| x1:x14 | -0.060044 | 0.025055 | -2.3965 | 0.02503 |
| x2:x11 | 0.13787 | 0.032584 | 4.2312 | 6.95E-05 |
| x3:x5 | 0.078607 | 0.027167 | 2.8935 | 0.00701 |
| x3:x8 | -0.22526 | 0.029655 | -7.596 | 6.15E-13 |
| x3:x11 | 0.3232 | 0.029161 | 11.083 | 0 |
| x3:x12 | 0.0088373 | 0.0026012 | 3.3974 | 0.0015 |
| x3:x15 | -0.8904 | 0.37182 | -2.3947 | 0.0251 |
| x4:x8 | 0.15143 | 0.03608 | 4.1971 | 7.97E-05 |
| x4:x11 | -0.36282 | 0.038143 | -9.5122 | 0 |
| x5:x8 | 0.29091 | 0.036896 | 7.8848 | 8.46E-14 |
| x5:x11 | -0.14723 | 0.03378 | -4.3586 | 4.06E-05 |
| x6:x10 | 0.22343 | 0.027907 | 8.0062 | 3.66E-14 |
| x7:x9 | -0.066402 | 0.022901 | -2.8995 | 0.00689 |
| x7:x11 | -0.099145 | 0.020526 | -4.8303 | 5.16E-06 |
| x8:x9 | 0.038182 | 0.014379 | 2.6554 | 0.01359 |
| x8:x11 | -0.065418 | 0.011601 | -5.6388 | 9.64E-08 |
| x8:x13 | -0.15984 | 0.029059 | -5.5007 | 2.01E-07 |
| x8:x15 | -3.6715 | 0.53941 | -6.8066 | 1.14E-10 |
| x8:x16_1 | -0.3597 | 0.069834 | -5.1508 | 1.12E-06 |
| x9:x10 | -0.032032 | 0.013357 | -2.3982 | 0.02496 |
| x10:x14 | 0.081362 | 0.029542 | 2.7541 | 0.01047 |
| x10:x15 | 2.3972 | 0.47992 | 4.9951 | 2.39E-06 |
| x11:x16_1 | 0.23292 | 0.058028 | 4.0138 | 0.00016 |
| x12:x14 | 0.0064412 | 0.0024247 | 2.6565 | 0.01359 |
| x14:x15 | -0.81659 | 0.26801 | -3.0469 | 0.00449 |
| x14:x16_1 | 0.15155 | 0.049576 | 3.0569 | 0.00437 |

#### Marijuana

|  | Estimate | Standard Error | t-stat | FDR-corrected p-values |
| --- | --- | --- | --- | --- |
| (Intercept) | 8.3068 | 0.34772 | 23.89 | 0 |
| x1 (VS) | -0.019352 | 0.24243 | -0.079828 | 0.94646 |
| x2 (Age) | -0.18552 | 0.24165 | -0.76773 | 0.48626 |
| x3 (Total Drinks 7 Days) | 0.78284 | 0.51027 | 1.5342 | 0.14825 |
| x4 (Drink Frequency 7 Days) | 0.45268 | 0.4113 | 1.1006 | 0.30801 |
| x5 (Drinks/Day 12 Months) | 0.63854 | 0.28214 | 2.2632 | 0.03322 |
| x6 (Drink Frequency 12 Months) | -1.1296 | 0.34158 | -3.307 | 0.002 |
| x7 (Total Tobacco 7 Days) | 1.9231 | 0.24825 | 7.7466 | 2.14E-13 |
| x8 (Tobacco Frequency 7 Days) | 0.52491 | 0.30152 | 1.7408 | 0.09886 |
| x9 (Cocaine) | 4.0786 | 0.34962 | 11.666 | 0 |
| x10 (Hallucinogens) | 1.1093 | 0.34118 | 3.2512 | 0.00239 |
| x11 (Opiates) | 0.47346 | 0.42039 | 1.1262 | 0.29672 |
| x12 (Sedatives) | 0.84531 | 0.29437 | 2.8716 | 0.00748 |
| x13 (Stimulants) | 0.64646 | 0.23954 | 2.6987 | 0.01217 |
| x14 (Father) | 0.25295 | 0.23984 | 1.0547 | 0.32907 |
| x16_1 (Gender) | 0.061843 | 0.51184 | 0.12082 | 0.92141 |
| x1:x8 | -0.50742 | 0.20172 | -2.5154 | 0.01917 |
| x1:x13 | 0.70711 | 0.27039 | 2.6152 | 0.01504 |
| x2:x7 | -0.5338 | 0.24006 | -2.2236 | 0.03613 |
| x3:x9 | -0.50275 | 0.14888 | -3.377 | 0.0016 |
| x4:x5 | -1.0985 | 0.27824 | -3.9481 | 0.00021 |
| x5:x6 | -1.0304 | 0.21604 | -4.7696 | 6.80E-06 |
| x6:x9 | 0.38691 | 0.18561 | 2.0846 | 0.04913 |
| x6:x16_1 | 0.82055 | 0.42794 | 1.9174 | 0.06945 |
| x7:x12 | -0.49909 | 0.1745 | -2.8601 | 0.00772 |
| x10:x14 | -0.80167 | 0.26792 | -2.9922 | 0.00528 |
| x11:x16_1 | -1.0505 | 0.44694 | -2.3505 | 0.02754 |

### Executive Control System

#### Cocaine

Executive Control System Cocaine
|  | Estimate | Standard Error | t-stat | FDR-corrected p-values |
| --- | --- | --- | --- | --- |
| x1 (ECS) | -0.031471 | 0.020638 | -1.5249 | 0.15056 |
| x2 (Age) | 0.098818 | 0.020121 | 4.9112 | 3.54E-06 |
| x3 (Total Drinks 7 Days) | -0.072847 | 0.061224 | -1.1898 | 0.269 |
| x4 (Drink Frequency 7 Days) | 0.049544 | 0.038299 | 1.2936 | 0.22726 |
| x5 (Drinks/Day 12 Months) | 0.071731 | 0.023154 | 3.098 | 0.00393 |
| x6 (Drink Frequency 12 Months) | 0.01366 | 0.02488 | 0.54902 | 0.62478 |
| x7 (Total Tobacco 7 Days) | 0.11902 | 0.058074 | 2.0494 | 0.05301 |
| x8 (Tobacco Frequency 7 Days) | 0.76604 | 0.092808 | 8.254 | 7.52E-15 |
| x9 (Hallucinogens) | 0.43617 | 0.11062 | 3.9431 | 0.00021 |
| x10 (Opiates) | -0.14972 | 0.11575 | -1.2935 | 0.22726 |
| x11 (Sedatives) | 0.37474 | 0.04997 | 7.4993 | 1.20E-12 |
| x12 (Stimulants) | -0.021634 | 0.0084403 | -2.5632 | 0.0171 |
| x13 (Marijuana) | -0.026332 | 0.019564 | -1.346 | 0.20838 |
| x14 (Father) | -0.02156 | 0.021297 | -1.0123 | 0.34812 |
| x15 (Mother) | -1.6654 | 0.49743 | -3.348 | 0.00176 |
| x16_1 (Gender) | -0.031925 | 0.055064 | -0.57979 | 0.60467 |
| x1:x7 | 0.061485 | 0.021668 | 2.8376 | 0.00824 |
| x1:x9 | 0.17194 | 0.036951 | 4.6534 | 1.15E-05 |
| x1:x10 | -0.2171 | 0.037325 | -5.8165 | 3.77E-08 |
| x1:x11 | -0.059511 | 0.022342 | -2.6637 | 0.01337 |
| x1:x14 | -0.059614 | 0.018062 | -3.3005 | 0.00204 |
| x2:x3 | 0.059518 | 0.023665 | 2.515 | 0.01917 |
| x2:x8 | 0.15265 | 0.029622 | 5.1532 | 1.12E-06 |
| x2:x9 | 0.18801 | 0.029605 | 6.3507 | 1.86E-09 |
| x3:x5 | 0.069751 | 0.024554 | 2.8407 | 0.00819 |
| x3:x8 | 0.13252 | 0.04014 | 3.3016 | 0.00203 |
| x3:x9 | 0.084688 | 0.041412 | 2.045 | 0.05349 |
| x3:x11 | -0.31281 | 0.0366 | -8.5467 | 0 |
| x3:x12 | 0.0064776 | 0.0029109 | 2.2253 | 0.0361 |
| x3:x13 | 0.086285 | 0.022464 | 3.841 | 0.00031 |
| x4:x8 | -0.18884 | 0.047314 | -3.9912 | 0.00018 |
| x4:x9 | -0.13914 | 0.045219 | -3.0769 | 0.00416 |
| x4:x11 | 0.36538 | 0.045181 | 8.087 | 2.24E-14 |
| x5:x7 | -0.044428 | 0.01896 | -2.3432 | 0.02789 |
| x5:x9 | 0.1906 | 0.037116 | 5.1354 | 1.22E-06 |
| x5:x11 | 0.13746 | 0.028916 | 4.7539 | 7.32E-06 |
| x6:x7 | 0.04789 | 0.018668 | 2.5653 | 0.0171 |
| x6:x8 | 0.14771 | 0.028293 | 5.2207 | 8.03E-07 |
| x6:x11 | -0.19084 | 0.030374 | -6.2829 | 2.78E-09 |
| x6:x12 | -0.0051556 | 0.0026829 | -1.9217 | 0.06889 |
| x7:x8 | 0.078983 | 0.022407 | 3.5248 | 0.00096 |
| x7:x9 | -0.054534 | 0.02224 | -2.452 | 0.02217 |
| x7:x11 | 0.085068 | 0.018651 | 4.5609 | 1.69E-05 |
| x7:x15 | -0.57584 | 0.29704 | -1.9386 | 0.06676 |
| x8:x11 | 0.076395 | 0.014183 | 5.3865 | 3.61E-07 |
| x8:x14 | 0.19187 | 0.023467 | 8.1763 | 1.23E-14 |
| x8:x15 | -3.7973 | 0.54146 | -7.013 | 3.19E-11 |
| x9:x10 | -0.025156 | 0.013487 | -1.8652 | 0.07748 |
| x9:x11 | 0.07289 | 0.017395 | 4.1904 | 8.19E-05 |
| x9:x14 | -0.31752 | 0.039577 | -8.023 | 3.39E-14 |
| x9:x15 | -2.9559 | 0.5444 | -5.4296 | 2.90E-07 |
| x9:x16_1 | 0.16725 | 0.086066 | 1.9433 | 0.06623 |
| x10:x11 | -0.11995 | 0.014434 | -8.3097 | 4.74E-15 |
| x10:x13 | -0.15598 | 0.022114 | -7.0536 | 2.46E-11 |
| x10:x15 | 2.2751 | 0.52492 | 4.3343 | 4.52E-05 |
| x10:x16_1 | 0.3902 | 0.091601 | 4.2598 | 6.20E-05 |
| x11:x13 | -0.057548 | 0.025445 | -2.2616 | 0.03325 |
| x11:x16_1 | -0.59073 | 0.059839 | -9.8719 | 0 |
| x12:x15 | 0.12277 | 0.04466 | 2.749 | 0.0106 |
| x12:x16_1 | 0.008566 | 0.0047225 | 1.8139 | 0.08561 |

#### Hallucinogens

|  | Estimate | Standard Error | t-stat | FDR-corrected p-values |
| --- | --- | --- | --- | --- |
| (Intercept) | -0.22681 | 0.094423 | -2.4021 | 0.02479 |
| x1 (ECS) | 0.07726 | 0.032183 | 2.4007 | 0.02484 |
| x2 (Age) | 0.0022272 | 0.028569 | 0.077959 | 0.94646 |
| x3 (Total Drinks 7 Days) | -0.032389 | 0.045157 | -0.71727 | 0.51643 |
| x4 (Drink Frequency 7 Days) | -0.0046904 | 0.036355 | -0.12902 | 0.91591 |
| x5 (Drinks/Day 12 Months) | -0.021125 | 0.024389 | -0.86614 | 0.42678 |
| x6 (Drink Frequency 12 Months) | 0.10623 | 0.029075 | 3.6538 | 0.00061 |
| x7 (Total Tobacco 7 Days) | -0.080646 | 0.022677 | -3.5563 | 0.00086 |
| x8 (Tobacco Frequency 7 Days) | 0.59539 | 0.12458 | 4.7793 | 6.54E-06 |
| x9 (Cocaine) | 0.14217 | 0.068832 | 2.0654 | 0.05131 |
| x10 (Opiates) | -0.66893 | 0.13481 | -4.962 | 2.76E-06 |
| x11 (Sedatives) | 0.70246 | 0.14078 | 4.9896 | 2.45E-06 |
| x12 (Stimulants) | 0.030857 | 0.0083792 | 3.6825 | 0.00056 |
| x13 (Marijuana) | 0.0065073 | 0.021284 | 0.30574 | 0.79094 |
| x14 (Father) | 0.06189 | 0.023545 | 2.6286 | 0.01459 |
| x15 (Mother) | 0.13255 | 0.5071 | 0.26139 | 0.82224 |
| x16_1 (Gender) | -0.02261 | 0.058349 | -0.38749 | 0.73988 |
| x1:x4 | 0.036806 | 0.020836 | 1.7665 | 0.09425 |
| x1:x8 | 0.050288 | 0.021219 | 2.37 | 0.02642 |
| x1:x13 | -0.039922 | 0.020628 | -1.9353 | 0.06717 |
| x1:x16_1 | -0.096729 | 0.043237 | -2.2372 | 0.03525 |
| x2:x9 | 0.13239 | 0.033821 | 3.9145 | 0.00024 |
| x2:x11 | -0.088736 | 0.033883 | -2.6189 | 0.01494 |
| x2:x12 | 0.006427 | 0.0024571 | 2.6157 | 0.01504 |
| x3:x9 | 0.11837 | 0.02814 | 4.2065 | 7.70E-05 |
| x3:x10 | -0.056919 | 0.029891 | -1.9042 | 0.07149 |
| x4:x5 | 0.053899 | 0.025363 | 2.1251 | 0.04498 |
| x4:x7 | -0.037749 | 0.021035 | -1.7946 | 0.08891 |
| x4:x9 | -0.12396 | 0.034019 | -3.6437 | 0.00063 |
| x4:x11 | 0.064681 | 0.026296 | 2.4598 | 0.02183 |
| x4:x13 | -0.053216 | 0.022399 | -2.3758 | 0.02624 |
| x5:x9 | 0.093314 | 0.047296 | 1.973 | 0.062 |
| x5:x10 | -0.11902 | 0.038521 | -3.0898 | 0.00402 |
| x5:x11 | -0.059315 | 0.028216 | -2.1022 | 0.04736 |
| x6:x8 | 0.14191 | 0.026411 | 5.373 | 3.86E-07 |
| x6:x10 | 0.07462 | 0.03775 | 1.9767 | 0.06165 |
| x6:x12 | -0.011127 | 0.0022261 | -4.9985 | 2.37E-06 |
| x6:x16_1 | -0.082258 | 0.036965 | -2.2253 | 0.0361 |
| x7:x8 | -0.069254 | 0.022405 | -3.091 | 0.00401 |
| x7:x11 | -0.070195 | 0.018432 | -3.8083 | 0.00035 |
| x8:x9 | 0.034384 | 0.013883 | 2.4767 | 0.02099 |
| x8:x13 | 0.064465 | 0.027173 | 2.3724 | 0.02634 |
| x8:x15 | -2.3105 | 0.67801 | -3.4078 | 0.00145 |
| x9:x11 | -0.12545 | 0.016288 | -7.7016 | 2.99E-13 |
| x9:x12 | 0.0083335 | 0.0035499 | 2.3475 | 0.02762 |
| x9:x13 | 0.11842 | 0.031964 | 3.7047 | 0.00052 |
| x9:x14 | 0.22351 | 0.04463 | 5.008 | 2.26E-06 |
| x10:x12 | -0.020697 | 0.0039431 | -5.249 | 6.99E-07 |
| x10:x14 | 0.066316 | 0.024384 | 2.7197 | 0.01148 |
| x10:x15 | 5.9728 | 0.74249 | 8.0443 | 3.02E-14 |
| x10:x16_1 | 0.58778 | 0.065169 | 9.0193 | 0 |
| x11:x12 | 0.025602 | 0.0041578 | 6.1575 | 5.78E-09 |
| x11:x14 | -0.078412 | 0.020079 | -3.9051 | 0.00025 |
| x11:x15 | -3.9675 | 0.69157 | -5.737 | 5.78E-08 |
| x11:x16_1 | -0.4959 | 0.072623 | -6.8284 | 1.01E-10 |
| x12:x15 | -0.073917 | 0.043568 | -1.6966 | 0.10813 |
| x12:x16_1 | 0.019456 | 0.0050381 | 3.8618 | 0.00029 |

#### Opiates

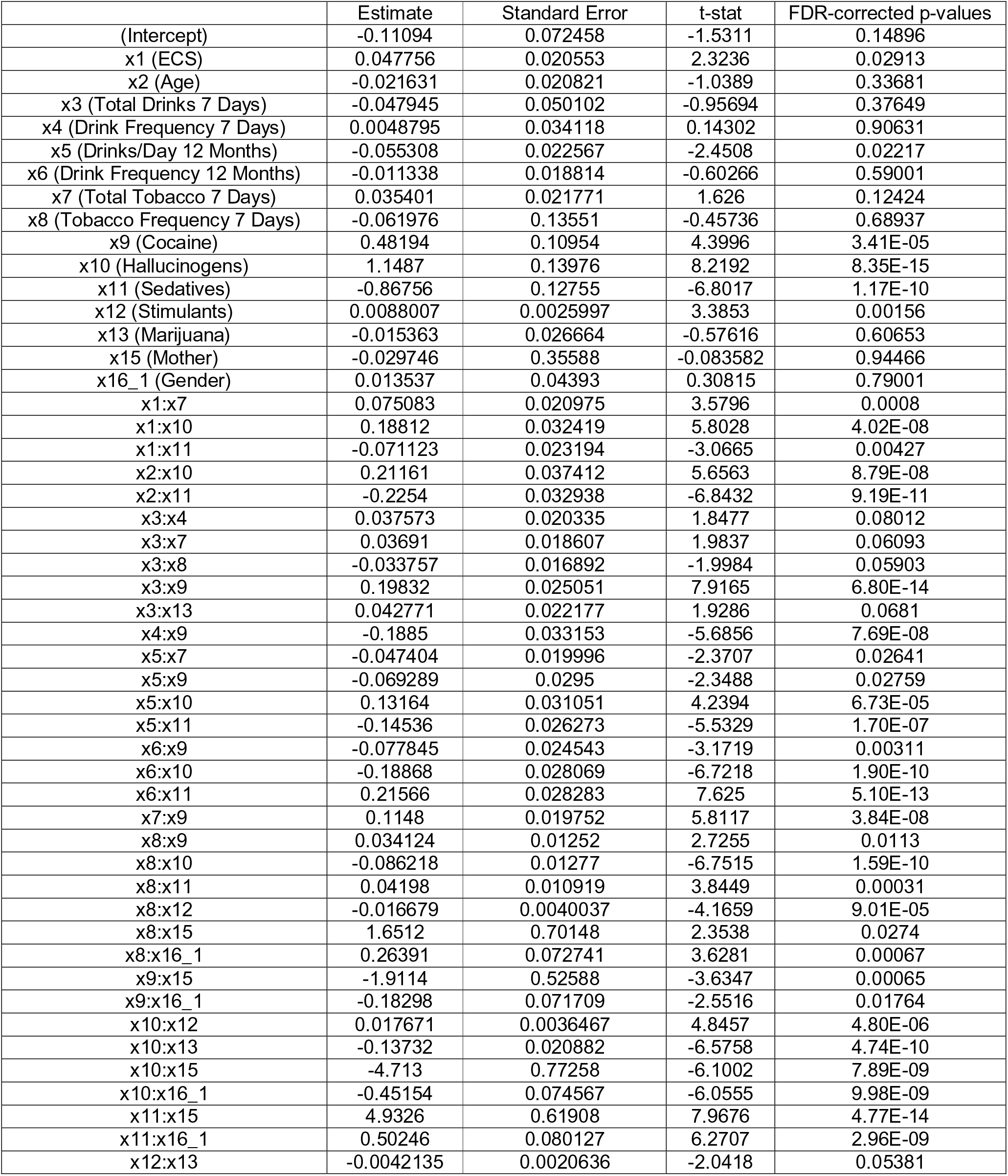

|  | Estimate | Standard Error | t-stat | FDR-corrected p-values |
| --- | --- | --- | --- | --- |
| (Intercept) | -0.11094 | 0.072458 | -1.5311 | 0.14896 |
| x1 (ECS) | 0.047756 | 0.020553 | 2.3236 | 0.02913 |
| x2 (Age) | -0.021631 | 0.020821 | -1.0389 | 0.33681 |
| x3 (Total Drinks 7 Days) | -0.047945 | 0.050102 | -0.95694 | 0.37649 |
| x4 (Drink Frequency 7 Days) | 0.0048795 | 0.034118 | 0.14302 | 0.90631 |
| x5 (Drinks/Day 12 Months) | -0.055308 | 0.022567 | -2.4508 | 0.02217 |
| x6 (Drink Frequency 12 Months) | -0.011338 | 0.018814 | -0.60266 | 0.59001 |
| x7 (Total Tobacco 7 Days) | 0.035401 | 0.021771 | 1.626 | 0.12424 |
| x8 (Tobacco Frequency 7 Days) | -0.061976 | 0.13551 | -0.45736 | 0.68937 |
| x9 (Cocaine) | 0.48194 | 0.10954 | 4.3996 | 3.41E-05 |
| x10 (Hallucinogens) | 1.1487 | 0.13976 | 8.2192 | 8.35E-15 |
| x11 (Sedatives) | -0.86756 | 0.12755 | -6.8017 | 1.17E-10 |
| x12 (Stimulants) | 0.0088007 | 0.0025997 | 3.3853 | 0.00156 |
| x13 (Marijuana) | -0.015363 | 0.026664 | -0.57616 | 0.60653 |
| x15 (Mother) | -0.029746 | 0.35588 | -0.083582 | 0.94466 |
| x16_1 (Gender) | 0.013537 | 0.04393 | 0.30815 | 0.79001 |
| x1:x7 | 0.075083 | 0.020975 | 3.5796 | 0.0008 |
| x1:x10 | 0.18812 | 0.032419 | 5.8028 | 4.02E-08 |
| x1:x11 | -0.071123 | 0.023194 | -3.0665 | 0.00427 |
| x2:x10 | 0.21161 | 0.037412 | 5.6563 | 8.79E-08 |
| x2:x11 | -0.2254 | 0.032938 | -6.8432 | 9.19E-11 |
| x3:x4 | 0.037573 | 0.020335 | 1.8477 | 0.08012 |
| x3:x7 | 0.03691 | 0.018607 | 1.9837 | 0.06093 |
| x3:x8 | -0.033757 | 0.016892 | -1.9984 | 0.05903 |
| x3:x9 | 0.19832 | 0.025051 | 7.9165 | 6.80E-14 |
| x3:x13 | 0.042771 | 0.022177 | 1.9286 | 0.0681 |
| x4:x9 | -0.1885 | 0.033153 | -5.6856 | 7.69E-08 |
| x5:x7 | -0.047404 | 0.019996 | -2.3707 | 0.02641 |
| x5:x9 | -0.069289 | 0.0295 | -2.3488 | 0.02759 |
| x5:x10 | 0.13164 | 0.031051 | 4.2394 | 6.73E-05 |
| x5:x11 | -0.14536 | 0.026273 | -5.5329 | 1.70E-07 |
| x6:x9 | -0.077845 | 0.024543 | -3.1719 | 0.00311 |
| x6:x10 | -0.18868 | 0.028069 | -6.7218 | 1.90E-10 |
| x6:x11 | 0.21566 | 0.028283 | 7.625 | 5.10E-13 |
| x7:x9 | 0.1148 | 0.019752 | 5.8117 | 3.84E-08 |
| x8:x9 | 0.034124 | 0.01252 | 2.7255 | 0.0113 |
| x8:x10 | -0.086218 | 0.01277 | -6.7515 | 1.59E-10 |
| x8:x11 | 0.04198 | 0.010919 | 3.8449 | 0.00031 |
| x8:x12 | -0.016679 | 0.0040037 | -4.1659 | 9.01E-05 |
| x8:x15 | 1.6512 | 0.70148 | 2.3538 | 0.0274 |
| x8:x16_1 | 0.26391 | 0.072741 | 3.6281 | 0.00067 |
| x9:x15 | -1.9114 | 0.52588 | -3.6347 | 0.00065 |
| x9:x16_1 | -0.18298 | 0.071709 | -2.5516 | 0.01764 |
| x10:x12 | 0.017671 | 0.0036467 | 4.8457 | 4.80E-06 |
| x10:x13 | -0.13732 | 0.020882 | -6.5758 | 4.74E-10 |
| x10:x15 | -4.713 | 0.77258 | -6.1002 | 7.89E-09 |
| x10:x16_1 | -0.45154 | 0.074567 | -6.0555 | 9.98E-09 |
| x11:x15 | 4.9326 | 0.61908 | 7.9676 | 4.77E-14 |
| x11:x16_1 | 0.50246 | 0.080127 | 6.2707 | 2.96E-09 |
| x12:x13 | -0.0042135 | 0.0020636 | -2.0418 | 0.05381 |

#### Sedatives

|  | Estimate | Standard Error | t-stat | FDR-corrected p-values |
| --- | --- | --- | --- | --- |
| (Intercept) | 0.063295 | 0.06235 | 1.0152 | 0.34706 |
| x1 (ECS) | 0.022653 | 0.017454 | 1.2979 | 0.22614 |
| x2 (Age) | -0.0063168 | 0.017389 | -0.36326 | 0.75515 |
| x3 (Total Drinks 7 Days) | 0.16392 | 0.07281 | 2.2513 | 0.03408 |
| x4 (Drink Frequency 7 Days) | -0.060771 | 0.029101 | -2.0882 | 0.04885 |
| x5 (Drinks/Day 12 Months) | 0.0063587 | 0.019491 | 0.32624 | 0.77862 |
| x6 (Drink Frequency 12 Months) | 0.026354 | 0.017924 | 1.4703 | 0.1666 |
| x7 (Total Tobacco 7 Days) | 0.012226 | 0.018608 | 0.65701 | 0.55494 |
| x8 (Tobacco Frequency 7 Days) | -0.67415 | 0.11159 | -6.0413 | 1.09E-08 |
| x9 (Cocaine) | 0.1641 | 0.05368 | 3.057 | 0.00437 |
| x10 (Hallucinogens) | 0.27159 | 0.072366 | 3.753 | 0.00043 |
| x11 (Opiates) | 1.1102 | 0.10969 | 10.121 | 0 |
| x12 (Stimulants) | -0.00077495 | 0.0022157 | -0.34976 | 0.76323 |
| x13 (Marijuana) | 0.17497 | 0.044005 | 3.9761 | 0.00019 |
| x14 (Father) | 0.0008118 | 0.017085 | 0.047516 | 0.96787 |
| x15 (Mother) | 0.18437 | 0.30614 | 0.60224 | 0.59001 |
| x16_1 (Gender) | -0.0027746 | 0.037746 | -0.073507 | 0.94819 |
| x1:x5 | 0.037386 | 0.017247 | 2.1677 | 0.04103 |
| x1:x8 | 0.052648 | 0.02148 | 2.451 | 0.02217 |
| x1:x9 | -0.07494 | 0.02616 | -2.8647 | 0.00763 |
| x1:x11 | 0.18558 | 0.023318 | 7.9586 | 5.06E-14 |
| x2:x10 | -0.18269 | 0.023965 | -7.6233 | 5.15E-13 |
| x2:x11 | 0.20098 | 0.027039 | 7.4329 | 1.88E-12 |
| x3:x4 | -0.041226 | 0.017809 | -2.3148 | 0.02972 |
| x3:x8 | -0.056293 | 0.02099 | -2.6818 | 0.01275 |
| x3:x9 | -0.20718 | 0.027782 | -7.4572 | 1.60E-12 |
| x3:x10 | -0.11854 | 0.024863 | -4.7676 | 6.87E-06 |
| x3:x11 | 0.41456 | 0.030515 | 13.585 | 0 |
| x3:x15 | -0.56774 | 0.30361 | -1.87 | 0.07679 |
| x4:x10 | 0.15499 | 0.029285 | 5.2924 | 5.79E-07 |
| x4:x11 | -0.44184 | 0.039512 | -11.182 | 0 |
| x5:x6 | 0.040378 | 0.014729 | 2.7415 | 0.01082 |
| x5:x7 | 0.032768 | 0.015676 | 2.0903 | 0.04869 |
| x5:x8 | -0.1174 | 0.030016 | -3.9111 | 0.00024 |
| x5:x9 | 0.40788 | 0.031176 | 13.083 | 0 |
| x5:x10 | -0.064984 | 0.026973 | -2.4092 | 0.0244 |
| x5:x11 | -0.23925 | 0.026356 | -9.0774 | 0 |
| x6:x8 | -0.16679 | 0.027628 | -6.0369 | 1.10E-08 |
| x6:x9 | 0.079715 | 0.030611 | 2.6042 | 0.01544 |
| x7:x8 | 0.090469 | 0.01705 | 5.3061 | 5.39E-07 |
| x7:x11 | -0.08084 | 0.017622 | -4.5875 | 1.52E-05 |
| x8:x9 | -0.12458 | 0.013819 | -9.0147 | 0 |
| x8:x10 | 0.065791 | 0.012247 | 5.372 | 3.86E-07 |
| x8:x11 | 0.065053 | 0.012399 | 5.2466 | 7.07E-07 |
| x8:x14 | -0.07299 | 0.018513 | -3.9426 | 0.00021 |
| x8:x15 | 5.6013 | 0.53161 | 10.536 | 0 |
| x8:x16_1 | -0.25341 | 0.062201 | -4.0741 | 0.00013 |
| x9:x10 | 0.12583 | 0.015716 | 8.0068 | 3.66E-14 |
| x9:x12 | 0.013536 | 0.0025597 | 5.2882 | 5.85E-07 |
| x9:x13 | 0.30761 | 0.024737 | 12.435 | 0 |
| x9:x16_1 | 0.24058 | 0.060539 | 3.974 | 0.00019 |
| x10:x11 | -0.12079 | 0.016654 | -7.2531 | 6.41E-12 |
| x10:x12 | -0.017375 | 0.0027485 | -6.3217 | 2.20E-09 |
| x10:x14 | 0.10847 | 0.022908 | 4.735 | 7.90E-06 |
| x10:x16_1 | 0.25037 | 0.061208 | 4.0905 | 0.00012 |
| x11:x13 | -0.16537 | 0.023859 | -6.9314 | 5.31E-11 |
| x11:x15 | -5.0012 | 0.54716 | -9.1402 | 0 |
| x11:x16_1 | -0.16684 | 0.063851 | -2.613 | 0.01511 |
| x13:x15 | -0.56159 | 0.22925 | -2.4497 | 0.0222 |

#### Stimulants

|  | Estimate | Standard Error | t-stat | FDR-corrected p-values |
| --- | --- | --- | --- | --- |
| (Intercept) | 0.11131 | 0.078811 | 1.4124 | 0.18556 |
| x1 (ECS) | -0.038318 | 0.023064 | -1.6614 | 0.11569 |
| x2 (Age) | -0.023192 | 0.02256 | -1.028 | 0.34208 |
| x3 (Total Drinks 7 Days) | 0.12014 | 0.089307 | 1.3453 | 0.20838 |
| x4 (Drink Frequency 7 Days) | -0.085533 | 0.037819 | -2.2616 | 0.03325 |
| x5 (Drinks/Day 12 Months) | 0.12023 | 0.025063 | 4.7973 | 6.02E-06 |
| x6 (Drink Frequency 12 Months) | 0.064702 | 0.02136 | 3.0291 | 0.00473 |
| x7 (Total Tobacco 7 Days) | -0.01706 | 0.024613 | -0.69314 | 0.53072 |
| x8 (Tobacco Frequency 7 Days) | 1.3016 | 0.14539 | 8.952 | 0 |
| x9 (Cocaine) | 0.32042 | 0.089353 | 3.586 | 0.00078 |
| x10 (Hallucinogens) | -0.12336 | 0.10733 | -1.1493 | 0.28653 |
| x11 (Opiates) | -0.29382 | 0.1409 | -2.0853 | 0.04913 |
| x12 (Sedatives) | 0.0051447 | 0.0029203 | 1.7617 | 0.09494 |
| x13 (Marijuana) | -0.014103 | 0.022828 | -0.61777 | 0.58116 |
| x14 (Father) | 0.021006 | 0.060127 | 0.34936 | 0.76323 |
| x15 (Mother) | -0.64329 | 0.4018 | -1.601 | 0.13053 |
| x16_1 (Gender) | 0.050649 | 0.049595 | 1.0213 | 0.3451 |
| x1:x8 | 0.078438 | 0.026103 | 3.0049 | 0.00509 |
| x1:x10 | -0.098128 | 0.031071 | -3.1582 | 0.00325 |
| x1:x11 | -0.25233 | 0.03497 | -7.2156 | 8.18E-12 |
| x2:x5 | -0.039931 | 0.020773 | -1.9223 | 0.06889 |
| x3:x8 | -0.24174 | 0.030043 | -8.0463 | 2.93E-14 |
| x3:x11 | 0.26766 | 0.028124 | 9.5172 | 0 |
| x3:x12 | 0.0078777 | 0.0025577 | 3.0799 | 0.00412 |
| x3:x15 | -0.70687 | 0.3854 | -1.8341 | 0.08222 |
| x4:x8 | 0.1713 | 0.037185 | 4.6068 | 1.41E-05 |
| x4:x11 | -0.26031 | 0.035715 | -7.2886 | 5.00E-12 |
| x5:x8 | 0.38852 | 0.035747 | 10.868 | 0 |
| x5:x10 | 0.1064 | 0.046633 | 2.2815 | 0.03199 |
| x5:x11 | -0.2372 | 0.042345 | -5.6015 | 1.18E-07 |
| x6:x10 | 0.27017 | 0.029604 | 9.1263 | 0 |
| x7:x8 | -0.091 | 0.02046 | -4.4478 | 2.77E-05 |
| x7:x13 | -0.03657 | 0.01851 | -1.9757 | 0.0617 |
| x8:x10 | 0.035504 | 0.018747 | 1.8938 | 0.07297 |
| x8:x11 | -0.056698 | 0.01254 | -4.5213 | 2.01E-05 |
| x8:x13 | -0.099466 | 0.028304 | -3.5141 | 0.001 |
| x8:x15 | -4.6699 | 0.72641 | -6.4287 | 1.16E-09 |
| x8:x16_1 | -0.49516 | 0.07509 | -6.5942 | 4.24E-10 |
| x9:x10 | -0.028789 | 0.016335 | -1.7624 | 0.09494 |
| x9:x15 | -1.4531 | 0.54717 | -2.6556 | 0.01359 |
| x10:x11 | -0.042229 | 0.014902 | -2.8339 | 0.00831 |
| x10:x15 | 2.8089 | 0.61931 | 4.5356 | 1.89E-05 |
| x11:x15 | 2.1681 | 0.7847 | 2.7629 | 0.01024 |
| x11:x16_1 | 0.5833 | 0.065393 | 8.9199 | 0 |
| x12:x14 | 0.0088012 | 0.0023364 | 3.7669 | 0.00041 |
| x14:x15 | -0.65884 | 0.25482 | -2.5856 | 0.01626 |
| x14:x16_1 | 0.10843 | 0.047345 | 2.2902 | 0.03144 |

#### Marijuana

|  | Estimate | Standard Error | t-stat | FDR-corrected p-values |
| --- | --- | --- | --- | --- |
| (Intercept) | 8.1708 | 0.2524 | 32.372 | 0 |
| x1 (ECS) | -0.0025895 | 0.23706 | -0.010923 | 0.99247 |
| x2 (Age) | -0.17051 | 0.23635 | -0.72146 | 0.51427 |
| x3 (Total Drinks 7 Days) | 0.84217 | 0.50579 | 1.6651 | 0.11514 |
| x4 (Drink Frequency 7 Days) | 0.3732 | 0.40998 | 0.91028 | 0.40216 |
| x5 (Drinks/Day 12 Months) | 0.6603 | 0.27926 | 2.3645 | 0.02676 |
| x6 (Drink Frequency 12 Months) | -0.77861 | 0.25241 | -3.0847 | 0.00406 |
| x7 (Total Tobacco 7 Days) | 1.9331 | 0.24641 | 7.8453 | 1.09E-13 |
| x9 (Cocaine) | 3.7293 | 0.29585 | 12.605 | 0 |
| x10 (Hallucinogens) | 0.98857 | 0.28357 | 3.4862 | 0.00109 |
| x12 (Sedatives) | 0.97543 | 0.27819 | 3.5064 | 0.00102 |
| x13 (Stimulants) | 0.55419 | 0.23598 | 2.3485 | 0.02759 |
| x14 (Father) | 0.35927 | 0.23685 | 1.5168 | 0.15272 |
| x1:x6 | 0.7277 | 0.2061 | 3.5308 | 0.00094 |
| x1:x13 | 0.45798 | 0.23166 | 1.9769 | 0.06165 |
| x2:x7 | -0.44289 | 0.23867 | -1.8557 | 0.07881 |
| x3:x9 | -0.54216 | 0.13978 | -3.8788 | 0.00027 |
| x4:x5 | -1.0871 | 0.27811 | -3.9088 | 0.00024 |
| x5:x6 | -0.86162 | 0.20531 | -4.1967 | 7.97E-05 |
| x7:x12 | -0.40667 | 0.16553 | -2.4568 | 0.02193 |
| x10:x14 | -0.62233 | 0.25907 | -2.4022 | 0.02479 |

### Valuation-Control Complex

#### Cocaine

Valuation-Control Complex Cocaine
|  | Estimate | Standard Error | t-stat | FDR-corrected p-values |
| --- | --- | --- | --- | --- |
| (Intercept) | 0.21801 | 0.092523 | 2.3562 | 0.02727 |
| x1 (VCC) | 0.0084324 | 0.028136 | 0.2997 | 0.79475 |
| x2 (Age) | 0.10037 | 0.020128 | 4.9865 | 2.47E-06 |
| x3 (Total Drinks 7 Days) | -0.10526 | 0.060086 | -1.7519 | 0.09684 |
| x4 (Drink Frequency 7 Days) | 0.064819 | 0.038244 | 1.6949 | 0.10836 |
| x5 (Drinks/Day 12 Months) | 0.074469 | 0.022767 | 3.2709 | 0.00224 |
| x6 (Drink Frequency 12 Months) | -0.013122 | 0.018856 | -0.69591 | 0.53021 |
| x7 (Total Tobacco 7 Days) | 0.0068708 | 0.021783 | 0.31542 | 0.7862 |
| x8 (Tobacco Frequency 7 Days) | 0.73355 | 0.092031 | 7.9707 | 4.72E-14 |
| x9 (Hallucinogens) | 0.46022 | 0.10924 | 4.213 | 7.50E-05 |
| x10 (Opiates) | -0.10223 | 0.11273 | -0.90693 | 0.4036 |
| x11 (Sedatives) | 0.42215 | 0.046208 | 9.1359 | 0 |
| x12 (Stimulants) | -0.021302 | 0.0084282 | -2.5275 | 0.01868 |
| x13 (Marijuana) | -0.024113 | 0.019626 | -1.2286 | 0.25296 |
| x14 (Father) | -0.021175 | 0.021218 | -0.99797 | 0.35535 |
| x15 (Mother) | -1.6435 | 0.4966 | -3.3095 | 0.00199 |
| x16_1 (Gender) | -0.074189 | 0.055707 | -1.3318 | 0.21321 |
| x1:x7 | 0.070669 | 0.023378 | 3.0228 | 0.00481 |
| x1:x9 | 0.18494 | 0.0346 | 5.345 | 4.43E-07 |
| x1:x10 | -0.19881 | 0.035036 | -5.6745 | 8.10E-08 |
| x1:x11 | -0.048278 | 0.021719 | -2.2228 | 0.03615 |
| x1:x12 | -0.0050319 | 0.0025167 | -1.9994 | 0.05903 |
| x1:x14 | -0.056556 | 0.019263 | -2.936 | 0.00622 |
| x2:x3 | 0.057609 | 0.023547 | 2.4466 | 0.02235 |
| x2:x8 | 0.14546 | 0.029644 | 4.9069 | 3.59E-06 |
| x2:x9 | 0.1923 | 0.028601 | 6.7236 | 1.90E-10 |
| x3:x5 | 0.074442 | 0.024544 | 3.0331 | 0.00469 |
| x3:x8 | 0.1322 | 0.039361 | 3.3586 | 0.0017 |
| x3:x9 | 0.091828 | 0.041076 | 2.2355 | 0.03533 |
| x3:x11 | -0.31607 | 0.036344 | -8.6967 | 0 |
| x3:x12 | 0.0084922 | 0.0025254 | 3.3627 | 0.00168 |
| x3:x13 | 0.09164 | 0.022421 | 4.0873 | 0.00012 |
| x4:x8 | -0.19438 | 0.047011 | -4.1348 | 0.0001 |
| x4:x9 | -0.15694 | 0.044449 | -3.5307 | 0.00094 |
| x4:x11 | 0.38012 | 0.044505 | 8.541 | 0 |
| x5:x7 | -0.054669 | 0.018642 | -2.9325 | 0.00628 |
| x5:x9 | 0.1659 | 0.035076 | 4.7297 | 8.09E-06 |
| x5:x11 | 0.12783 | 0.028815 | 4.4362 | 2.89E-05 |
| x6:x8 | 0.14028 | 0.025184 | 5.5701 | 1.39E-07 |
| x6:x11 | -0.19896 | 0.029957 | -6.6416 | 3.16E-10 |
| x7:x8 | 0.090095 | 0.022058 | 4.0846 | 0.00012 |
| x7:x9 | -0.084401 | 0.020769 | -4.0638 | 0.00013 |
| x7:x11 | 0.088186 | 0.018604 | 4.7401 | 7.74E-06 |
| x8:x11 | 0.066431 | 0.012139 | 5.4727 | 2.31E-07 |
| x8:x14 | 0.20564 | 0.024269 | 8.4733 | 1.70E-15 |
| x8:x15 | -3.4857 | 0.53358 | -6.5327 | 6.14E-10 |
| x9:x11 | 0.061953 | 0.016777 | 3.6926 | 0.00054 |
| x9:x14 | -0.31718 | 0.039307 | -8.0695 | 2.46E-14 |
| x9:x15 | -3.3182 | 0.54241 | -6.1176 | 7.21E-09 |
| x9:x16_1 | 0.21742 | 0.081011 | 2.6838 | 0.0127 |
| x10:x11 | -0.11906 | 0.014158 | -8.4095 | 1.70E-15 |
| x10:x13 | -0.1508 | 0.021665 | -6.9606 | 4.47E-11 |
| x10:x15 | 2.1964 | 0.52507 | 4.1831 | 8.43E-05 |
| x10:x16_1 | 0.27352 | 0.080828 | 3.384 | 0.00157 |
| x11:x13 | -0.067314 | 0.025331 | -2.6573 | 0.01359 |
| x11:x16_1 | -0.611 | 0.060297 | -10.133 | 0 |
| x12:x15 | 0.11112 | 0.043979 | 2.5266 | 0.01869 |
| x12:x16_1 | 0.012633 | 0.0049171 | 2.5692 | 0.01694 |

#### Hallucinogens

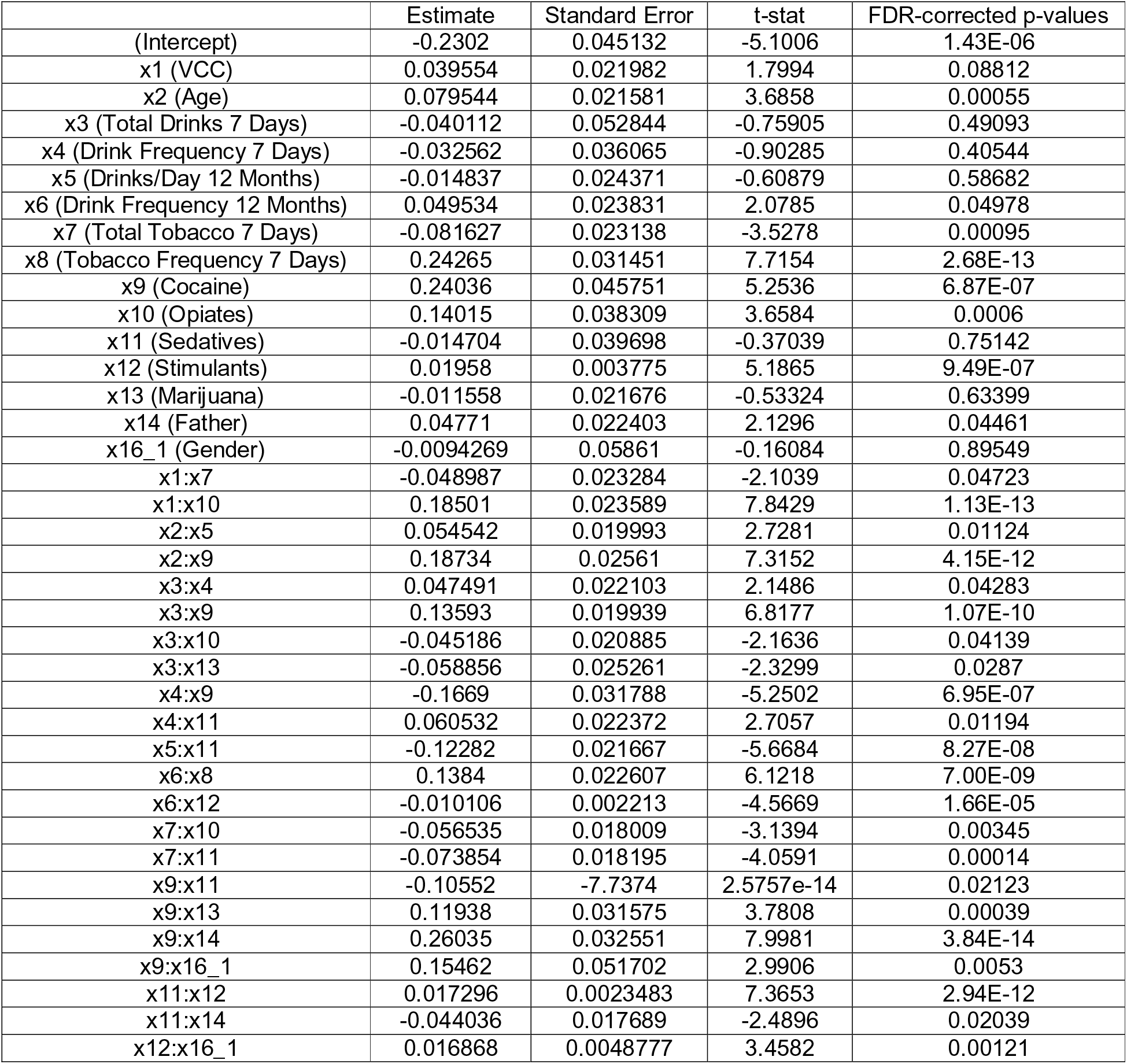

#### Opiates

|  | Estimate | Standard Error | t-stat | FDR-corrected p-values |
| --- | --- | --- | --- | --- |
| (Intercept) | -0.13254 | 0.072169 | -1.8365 | 0.08202 |
| x1 (VCC) | 0.040253 | 0.020747 | 1.9402 | 0.0666 |
| x2 (Age) | -0.047837 | 0.021001 | -2.2778 | 0.03225 |
| x3 (Total Drinks 7 Days) | -0.036696 | 0.050317 | -0.72928 | 0.5097 |
| x4 (Drink Frequency 7 Days) | 0.0079581 | 0.034373 | 0.23153 | 0.84409 |
| x5 (Drinks/Day 12 Months) | -0.05452 | 0.022952 | -2.3754 | 0.02624 |
| x6 (Drink Frequency 12 Months) | -0.0063606 | 0.019117 | -0.33272 | 0.77445 |
| x7 (Total Tobacco 7 Days) | 0.03682 | 0.022141 | 1.663 | 0.11547 |
| x8 (Tobacco Frequency 7 Days) | -0.11278 | 0.054631 | -2.0644 | 0.05135 |
| x9 (Cocaine) | 0.525 | 0.093189 | 5.6337 | 9.84E-08 |
| x10 (Hallucinogens) | 0.64991 | 0.044538 | 14.592 | 0 |
| x11 (Sedatives) | -0.60287 | 0.095112 | -6.3385 | 1.98E-09 |
| x12 (Stimulants) | 0.0079989 | 0.0026187 | 3.0546 | 0.00439 |
| x13 (Marijuana) | -0.046051 | 0.019696 | -2.338 | 0.02823 |
| x15 (Mother) | 0.060994 | 0.35664 | 0.17102 | 0.88857 |
| x16_1 (Gender) | 0.010387 | 0.044234 | 0.23482 | 0.84248 |
| x1:x5 | -0.045838 | 0.020606 | -2.2245 | 0.03611 |
| x1:x7 | 0.096573 | 0.022107 | 4.3685 | 3.91E-05 |
| x1:x10 | 0.08931 | 0.02511 | 3.5567 | 0.00086 |
| x2:x9 | 0.063258 | 0.028105 | 2.2508 | 0.03408 |
| x2:x11 | -0.21705 | 0.029353 | -7.3945 | 2.42E-12 |
| x3:x4 | 0.037971 | 0.020457 | 1.8561 | 0.07881 |
| x3:x7 | 0.044184 | 0.018199 | 2.4278 | 0.0234 |
| x3:x9 | 0.18642 | 0.022677 | 8.2207 | 8.35E-15 |
| x4:x9 | -0.17706 | 0.034732 | -5.098 | 1.45E-06 |
| x5:x7 | -0.0482 | 0.01983 | -2.4307 | 0.02326 |
| x5:x11 | -0.11275 | 0.021446 | -5.2575 | 6.75E-07 |
| x6:x9 | -0.056763 | 0.023394 | -2.4264 | 0.02345 |
| x6:x10 | -0.18749 | 0.027124 | -6.9124 | 5.91E-11 |
| x6:x11 | 0.20665 | 0.027502 | 7.514 | 1.08E-12 |
| x7:x9 | 0.10117 | 0.019503 | 5.1874 | 9.49E-07 |
| x8:x10 | -0.056695 | 0.0082332 | -6.8861 | 6.93E-11 |
| x8:x11 | 0.047575 | 0.0094879 | 5.0142 | 2.20E-06 |
| x8:x16_1 | 0.1865 | 0.062501 | 2.9839 | 0.00538 |
| x9:x15 | -2.2217 | 0.44545 | -4.9876 | 2.46E-06 |
| x9:x16_1 | -0.20784 | 0.062155 | -3.3439 | 0.00178 |
| x10:x13 | -0.094492 | 0.017929 | -5.2702 | 6.37E-07 |
| x10:x16_1 | -0.21276 | 0.064134 | -3.3174 | 0.00194 |
| x11:x15 | 3.3288 | 0.40243 | 8.2716 | 6.22E-15 |
| x11:x16_1 | 0.38016 | 0.074412 | 5.1088 | 1.38E-06 |

#### Sedatives

|  | Estimate | Standard Error | t-stat | FDR-corrected p-values |
| --- | --- | --- | --- | --- |
| (Intercept) | 0.024389 | 0.062107 | 0.39269 | 0.73685 |
| x1 (VCC) | 0.013669 | 0.017229 | 0.79342 | 0.47029 |
| x2 (Age) | -0.0066322 | 0.017301 | -0.38333 | 0.7422 |
| x3 (Total Drinks 7 Days) | 0.1868 | 0.072644 | 2.5715 | 0.01687 |
| x4 (Drink Frequency 7 Days) | -0.063683 | 0.029032 | -2.1936 | 0.03869 |
| x5 (Drinks/Day 12 Months) | 0.0020966 | 0.019542 | 0.10728 | 0.93122 |
| x6 (Drink Frequency 12 Months) | 0.02655 | 0.017838 | 1.4884 | 0.16114 |
| x7 (Total Tobacco 7 Days) | 0.010509 | 0.018515 | 0.56762 | 0.61196 |
| x8 (Tobacco Frequency 7 Days) | -0.64186 | 0.11007 | -5.8316 | 3.47E-08 |
| x9 (Cocaine) | 0.18519 | 0.080406 | 2.3032 | 0.03054 |
| x10 (Hallucinogens) | -0.12577 | 0.11793 | -1.0665 | 0.32389 |
| x11 (Opiates) | 1.284 | 0.13956 | 9.2003 | 0 |
| x12 (Stimulants) | -0.00034913 | 0.0022039 | -0.15841 | 0.89549 |
| x13 (Marijuana) | 0.17437 | 0.043753 | 3.9853 | 0.00018 |
| x14 (Father) | 0.0016467 | 0.017034 | 0.096672 | 0.93814 |
| x15 (Mother) | 0.30026 | 0.30469 | 0.98546 | 0.36168 |
| x16_1 (Gender) | 0.02026 | 0.037392 | 0.54183 | 0.62847 |
| x1:x5 | 0.039047 | 0.017251 | 2.2635 | 0.03322 |
| x1:x8 | 0.082159 | 0.022537 | 3.6454 | 0.00063 |
| x1:x10 | -0.13514 | 0.029037 | -4.6541 | 1.15E-05 |
| x1:x11 | 0.18292 | 0.022003 | 8.3134 | 4.74E-15 |
| x2:x9 | -0.16846 | 0.02444 | -6.893 | 6.79E-11 |
| x2:x11 | 0.18293 | 0.02869 | 6.376 | 1.60E-09 |
| x3:x4 | -0.042658 | 0.017883 | -2.3854 | 0.02571 |
| x3:x8 | -0.050011 | 0.020661 | -2.4205 | 0.02375 |
| x3:x9 | -0.1456 | 0.027275 | -5.3382 | 7.88E-08 |
| x3:x10 | -0.20886 | 0.027533 | -7.586 | 6.57E-13 |
| x3:x11 | 0.43029 | 0.031252 | 13.768 | 0 |
| x3:x15 | -0.68629 | 0.30292 | -2.2655 | 0.03319 |
| x4:x9 | 0.18042 | 0.030779 | 5.8618 | 2.96E-08 |
| x4:x11 | -0.46683 | 0.040112 | -11.638 | 0 |
| x5:x6 | 0.041472 | 0.014618 | 2.8371 | 0.00824 |
| x5:x7 | 0.031309 | 0.015692 | 1.9952 | 0.05939 |
| x5:x8 | -0.10872 | 0.030201 | -3.6 | 0.00074 |
| x5:x9 | -0.062916 | 0.027422 | -2.2943 | 0.03115 |
| x5:x10 | 0.38104 | 0.031854 | 11.962 | 0 |
| x5:x11 | -0.24011 | 0.026315 | -9.1248 | 0 |
| x6:x8 | -0.16926 | 0.027264 | -6.2079 | 4.32E-09 |
| x6:x10 | 0.07902 | 0.030093 | 2.6259 | 0.01467 |
| x7:x8 | 0.085114 | 0.017089 | 4.9806 | 2.54E-06 |
| x7:x11 | -0.087971 | 0.0177 | -4.9701 | 2.66E-06 |
| x8:x9 | 0.06194 | 0.012688 | 4.8817 | 4.07E-06 |
| x8:x10 | -0.12195 | 0.013914 | -8.7646 | 0 |
| x8:x11 | 0.069098 | 0.012761 | 5.4146 | 3.13E-07 |
| x8:x14 | -0.069737 | 0.018435 | -3.783 | 0.00039 |
| x8:x15 | 5.4962 | 0.52859 | 10.398 | 0 |
| x8:x16_1 | -0.2812 | 0.06243 | -4.5043 | 2.18E-05 |
| x9:x10 | 0.11883 | 0.015933 | 7.4584 | 1.60E-12 |
| x9:x11 | -0.11238 | 0.01855 | -6.0583 | 9.89E-09 |
| x9:x12 | -0.014053 | 0.0030793 | -4.5635 | 1.68E-05 |
| x9:x14 | 0.09909 | 0.022882 | 4.3305 | 4.57E-05 |
| x9:x16_1 | 0.24428 | 0.061687 | 3.96 | 0.0002 |
| x10:x12 | 0.015054 | 0.0026221 | 5.7414 | 5.70E-08 |
| x10:x13 | 0.30791 | 0.024809 | 12.411 | 0 |
| x10:x15 | 1.3966 | 0.58817 | 2.3746 | 0.02624 |
| x10:x16_1 | 0.35179 | 0.068092 | 5.1663 | 1.05E-06 |
| x11:x12 | -0.0058883 | 0.0032675 | -1.8021 | 0.08774 |
| x11:x13 | -0.16606 | 0.023821 | -6.9713 | 4.20E-11 |
| x11:x15 | -5.6098 | 0.6812 | -8.2352 | 7.52E-15 |
| x11:x16_1 | -0.16617 | 0.064809 | -2.564 | 0.0171 |
| x13:x15 | -0.56871 | 0.22808 | -2.4935 | 0.02024 |

#### Stimulants

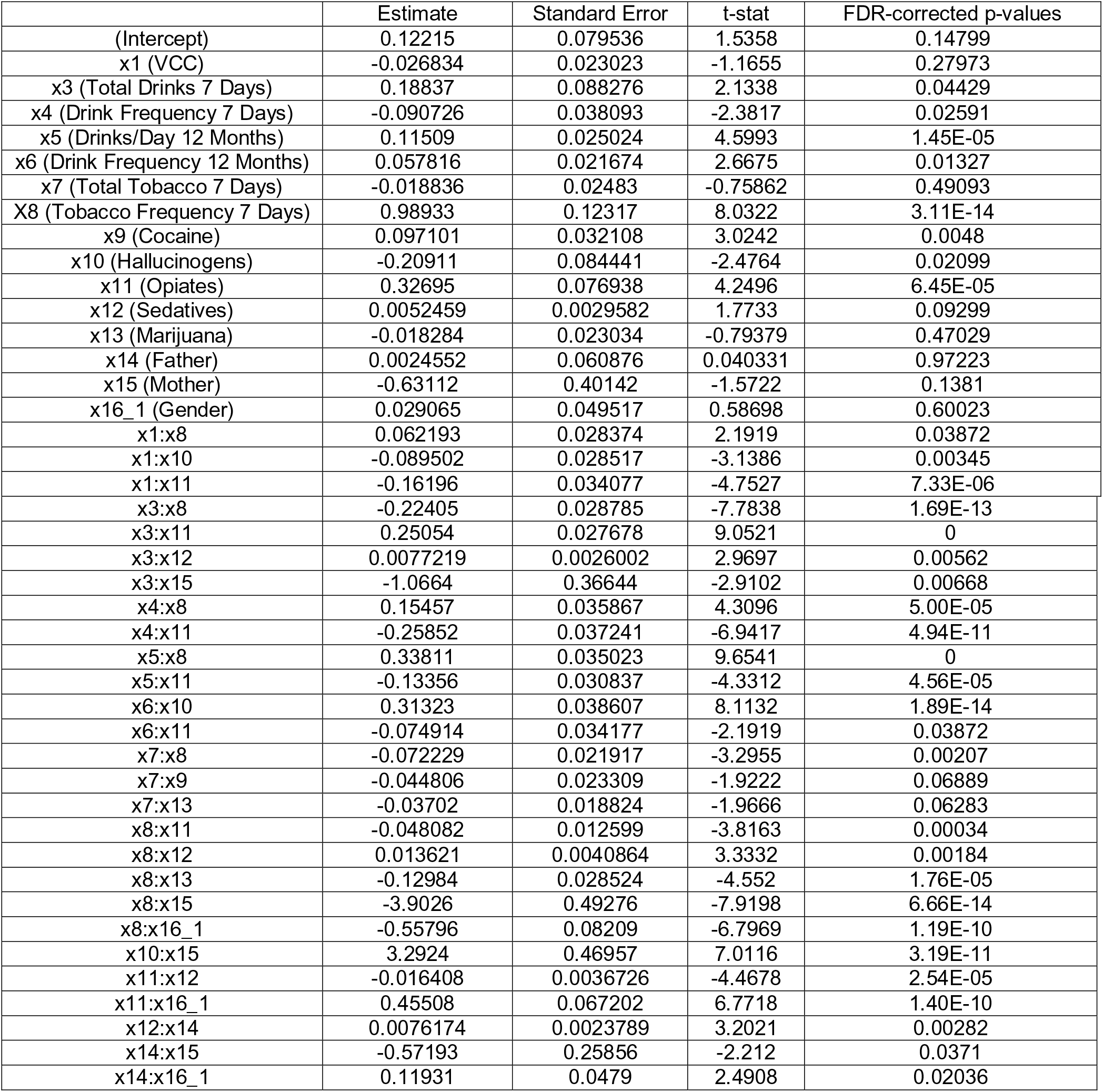

#### Marijuana

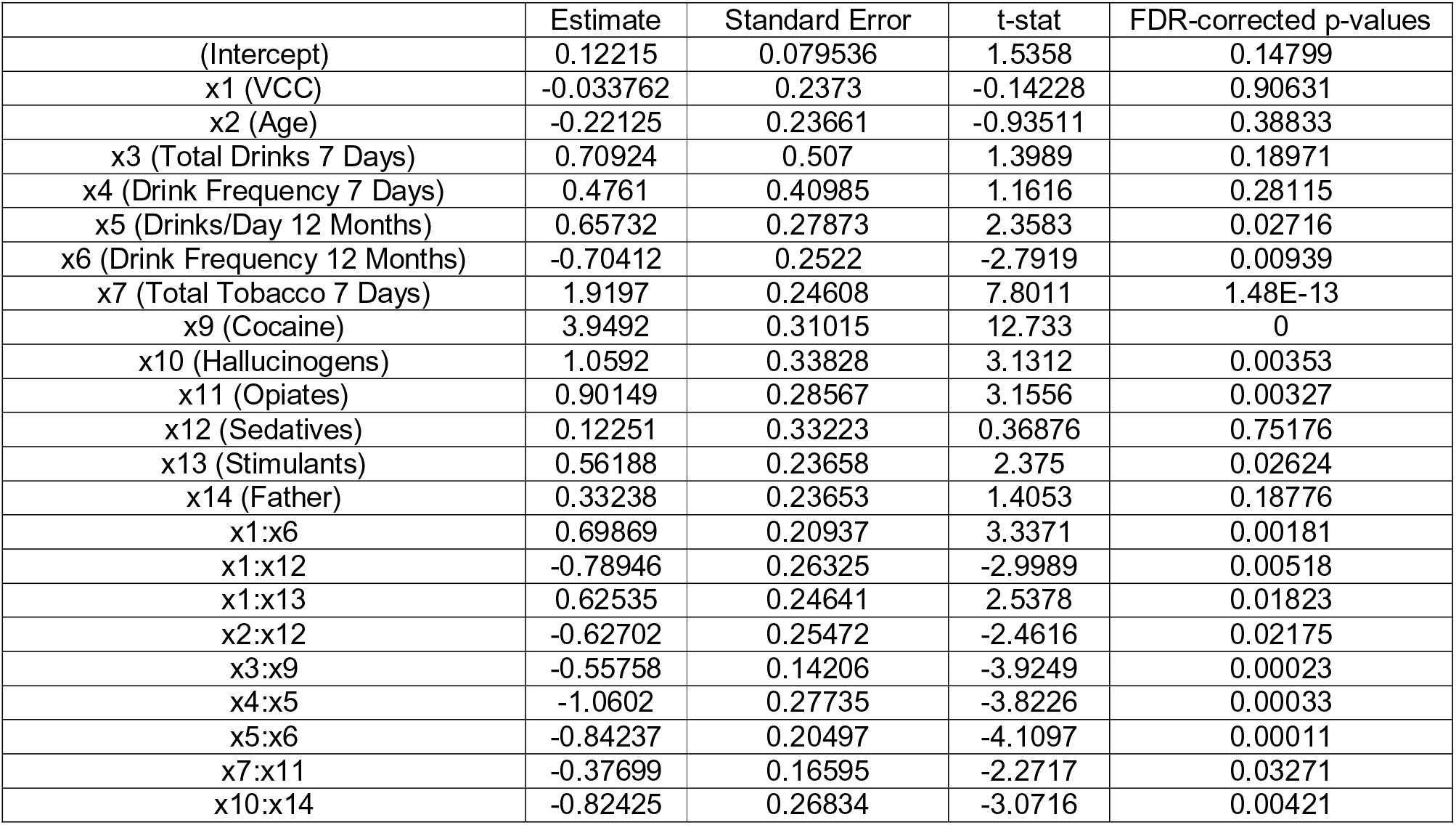

### Supplementary Analysis: Generalized Linear Models without Drink Frequency 7 Days

Given that the variables Total Drinks 7 Days and Drink Frequency 7 Days were highly correlated, we attempted to repeat the GLM analysis but after removing Drink Frequency 7 Days as a covariate. Results are shown in Supplementary Table 12. The most telling difference in results after the removal of Drink Frequency 7 Days was the complete absence of ECS connectivity correlated with Hallucinogen use. Seeing as how Drink Frequency 7 Days was originally a significant covariate in Hallucinogen use (via a two-way interaction with ECS connectivity), its removal may have something to do with the reduced relevance of ECS connectivity with regards to Hallucinogen use. Most other models also revealed reduced r^2^ values after the removal of Drink Frequency 7 Days as a covariate (except for VS and VCC models correlated with Hallucinogen use and VCC correlated with Marijuana use which all reported increased r^2^ values). The removal of a covariate that formed significant interactions with network connectivity from the modelling process may have resulted in the reduced effect sizes reported. We cannot ascertain why the r^2^ values increased in Hallucinogen use and VCC – Marijuana use. Despite these changes, many other interactions between network connectivity and substance use remained the same.

**Supplementary Table 12:**
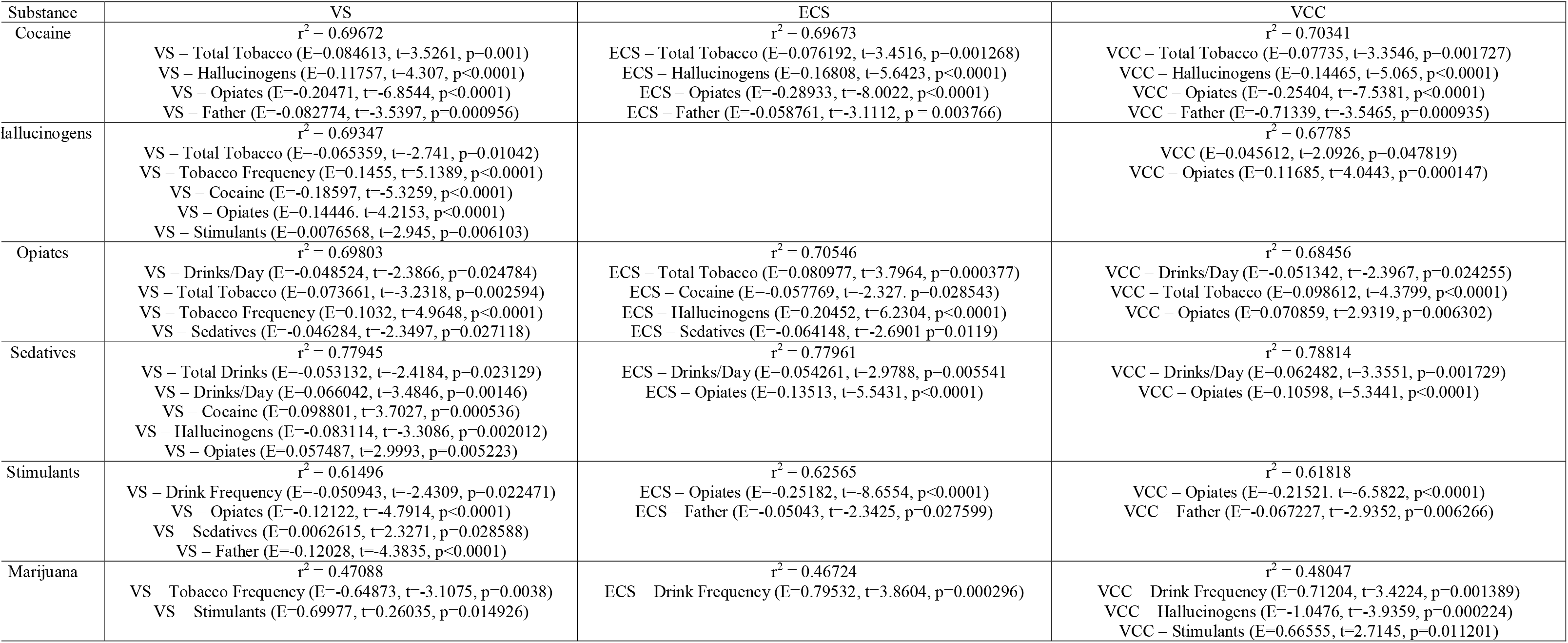
VS, ECS and VCC connectivity correlations with drug use after removing Drink Frequency 7 Days as a covariate. Effect size denoted by r^2^ values. Estimates displayed are standardized partial coefficients for each component of the GLM. *p*-values shown here are FDR-corrected. Exact p-values are displayed where possible. If the p-value is too small (less than 0.00001), then it is displayed as p<0.00001. Father, Fathers’ Use History; Mother, Mothers’ Use History; E, Estimate

## References

Anderson, J.S., Druzgal, T.J., Lopez-Larson, M., Jeong, E.-K., Desai, K., Yurgelun-Todd, D., 2011. Network anticorrelations, global regression, and phase-shifted soft tissue correction. Hum. Brain Mapp. 32, 919–934. https://doi.org/10.1002/hbm.21079

Anderson, R.E., Hruska, B., Boros, A.P., Richardson, C.J., Delahanty, D.L., 2018. Patterns of co-occurring addictions, posttraumatic stress disorder, and major depressive disorder in detoxification treatment seekers: Implications for improving detoxification treatment outcomes. J. Subst. Abuse Treat. 86, 45–51. https://doi.org/10.1016/j.jsat.2017.12.009

Bedi, G., Lindquist, M.A., Haney, M., 2015. An fMRI-Based Neural Signature of Decisions to Smoke Cannabis. Neuropsychopharmacol. Off. Publ. Am. Coll. Neuropsychopharmacol. 40, 2657–2665. https://doi.org/10.1038/npp.2015.135

Bellone, C., Loureiro, M., Lüscher, C., 2020. Drug-Evoked Synaptic Plasticity of Excitatory Transmission in the Ventral Tegmental Area. Cold Spring Harb. Perspect. Med. https://doi.org/10.1101/cshperspect.a039701

Breiter, H.C., Gollub, R.L., Weisskoff, R.M., Kennedy, D.N., Makris, N., Berke, J.D., Goodman, J.M., Kantor, H.L., Gastfriend, D.R., Riorden, J.P., Mathew, R.T., Rosen, B.R., Hyman, S.E., 1997. Acute effects of cocaine on human brain activity and emotion. Neuron 19, 591–611.

Bring, J., 1994. How to Standardize Regression Coefficients. Am. Stat. 48, 209–213. https://doi.org/10.1080/00031305.1994.10476059

Choi, N.G., DiNitto, D.M., Marti, C.N., Choi, B.Y., 2017. Association between Nonmedical Marijuana and Pain Reliever Uses among Individuals Aged 50. J. Psychoactive Drugs 49, 267–278. https://doi.org/10.1080/02791072.2017.1342153

Ciric, R., Rosen, A.F.G., Erus, G., Cieslak, M., Adebimpe, A., Cook, P.A., Bassett, D.S., Davatzikos, C., Wolf, D.H., Satterthwaite, T.D., 2018. Mitigating head motion artifact in functional connectivity MRI. Nat. Protoc. 13, 2801–2826. https://doi.org/10.1038/s41596-018-0065-y

Coffin, P.O., Galea, S., Ahern, J., Leon, A.C., Vlahov, D., Tardiff, K., 2003. Opiates, cocaine and alcohol combinations in accidental drug overdose deaths in New York City, 1990– 98. Addiction 98, 739–747.

Compton, W.M., Thomas, Y.F., Stinson, F.S., Grant, B.F., 2007. Prevalence, correlates, disability, and comorbidity of DSM-IV drug abuse and dependence in the United States: results from the national epidemiologic survey on alcohol and related conditions. Arch. Gen. Psychiatry 64, 566–576. https://doi.org/10.1001/archpsyc.64.5.566

Crane, N.A., Gorka, S.M., Weafer, J., Langenecker, S.A., de Wit, H., Phan, K.L., 2018. Neural activation to monetary reward is associated with amphetamine reward sensitivity. Neuropsychopharmacol. Off. Publ. Am. Coll. Neuropsychopharmacol. 43, 1738–1744. https://doi.org/10.1038/s41386-018-0042-8

Crean, R.D., Crane, N.A., Mason, B.J., 2011. An Evidence-Based Review of Acute and Long-Term Effects of Cannabis Use on Executive Cognitive Functions. J. Addict. Med. 5.

Cross, S.J., Lotfipour, S., Leslie, F.M., 2017. Mechanisms and genetic factors underlying co-use of nicotine and alcohol or other drugs of abuse. Am. J. Drug Alcohol Abuse 43, 171–185. https://doi.org/10.1080/00952990.2016.1209512

Dalley, J.W., Everitt, B.J., Robbins, T.W., 2011. Impulsivity, compulsivity, and top-down cognitive control. Neuron 69, 680–694. https://doi.org/10.1016/j.neuron.2011.01.020

Davis, W.R., Johnson, B.D., 2008. Prescription opioid use, misuse, and diversion among street drug users in New York City. Drug Alcohol Depend. 92, 267–276. https://doi.org/10.1016/j.drugalcdep.2007.08.008

Desikan, R.S., Ségonne, F., Fischl, B., Quinn, B.T., Dickerson, B.C., Blacker, D., Buckner, R.L., Dale, A.M., Maguire, R.P., Hyman, B.T., Albert, M.S., Killiany, R.J., 2006. An automated labeling system for subdividing the human cerebral cortex on MRI scans into gyral based regions of interest. Neuroimage 31, 968–980. https://doi.org/10.1016/j.neuroimage.2006.01.021

Destrieux, C., Fischl, B., Dale, A., Halgren, E., 2010. Automatic parcellation of human cortical gyri and sulci using standard anatomical nomenclature. Neuroimage 53, 1–15. https://doi.org/10.1016/j.neuroimage.2010.06.010

Devonshire, I.M., Mayhew, J.E.W., Overton, P.G., 2007. Cocaine preferentially enhances sensory processing in the upper layers of the primary sensory cortex. Neuroscience 146, 841–851. https://doi.org/10.1016/j.neuroscience.2007.01.070

Dlugos, A.M., Hamidovic, A., Hodgkinson, C., Shen, P.H., Goldman, D., Palmer, A.A., de Wit, H., 2011. OPRM1 gene variants modulate amphetamine-induced euphoria in humans. Genes. Brain. Behav. 10, 199–209. https://doi.org/10.1111/j.1601-183X.2010.00655.x

Downey, L.A., King, R., Papafotiou, K., Swann, P., Ogden, E., Boorman, M., Stough, C., 2013. The effects of cannabis and alcohol on simulated driving: Influences of dose and experience. Accid. Anal. Prev. 50, 879–886. https://doi.org/10.1016/j.aap.2012.07.016

Ersche, K.D., Meng, C., Ziauddeen, H., Stochl, J., Williams, G.B., Bullmore, E.T., Robbins, T.W., 2020. Brain networks underlying vulnerability and resilience to drug addiction. Proc. Natl. Acad. Sci. U. S. A. 117, 15253–15261. https://doi.org/10.1073/pnas.2002509117

Fisk, J.E., Murphy, P.N., Montgomery, C., Hadjiefthyvoulou, F., 2011. Modelling the adverse effects associated with ecstasy use. Addiction 106, 798–805. https://doi.org/10.1111/j.1360-0443.2010.03272.x

Floyd, L.J., Hedden, S., Lawson, A., Salama, C., Moleko, A.G., Latimer, W., 2010. The association between poly-substance use, coping, and sex trade among black South African substance users. Subst. Use Misuse 45, 1971–1987. https://doi.org/10.3109/10826081003767635

Fultz, E.K., Coelho, M.A., Lieberman, D., Jimenez-Chavez, C.L., Bryant, C.D., Szumlinski, K.K., 2021. Hnrnph1 is a novel regulator of alcohol reward. Drug Alcohol Depend. 220, 108518.

Geliebter, A., Benson, L., Pantazatos, S.P., Hirsch, J., Carnell, S., 2016. Greater anterior cingulate activation and connectivity in response to visual and auditory high-calorie food cues in binge eating: Preliminary findings. Appetite 96, 195–202. https://doi.org/10.1016/j.appet.2015.08.009

Givens, C.J., 2016. Adverse drug reactions associated with antipsychotics, antidepressants, mood stabilizers, and stimulants. Nurs. Clin. 51, 309–321.

Glasser, M.F., Sotiropoulos, S.N., Wilson, J.A., Coalson, T.S., Fischl, B., Andersson, J.L., Xu, J., Jbabdi, S., Webster, M., Polimeni, J.R., Van Essen, D.C., Jenkinson, M., Consortium, W.-M.H.C.P., 2013. The minimal preprocessing pipelines for the Human Connectome Project. Neuroimage 80, 105–124. https://doi.org/10.1016/j.neuroimage.2013.04.127

Helie, S., Shamloo, F., Novak, K., Foti, D., 2017. The roles of valuation and reward processing in cognitive function and psychiatric disorders. Ann. N. Y. Acad. Sci. 1395, 33–48. https://doi.org/10.1111/nyas.13327

Hellberg, S.N., Russell, T.I., Robinson, M.J.F., 2019. Cued for risk: Evidence for an incentive sensitization framework to explain the interplay between stress and anxiety, substance abuse, and reward uncertainty in disordered gambling behavior. Cogn. Affect. Behav. Neurosci. 19, 737–758. https://doi.org/10.3758/s13415-018-00662-3

Hernández-López, C., Farré, M., Roset, P.N., Menoyo, E., Pizarro, N., Ortuño, J., Torrens, M., Camí, J., de La Torre, R., 2002. 3,4-Methylenedioxymethamphetamine (ecstasy) and alcohol interactions in humans: psychomotor performance, subjective effects, and pharmacokinetics. J. Pharmacol. Exp. Ther. 300, 236–244. https://doi.org/10.1124/jpet.300.1.236

Hiebler-Ragger, M., Unterrainer, H.-F., 2019. The Role of Attachment in Poly-Drug Use Disorder: An Overview of the Literature, Recent Findings and Clinical Implications. Front. psychiatry 10, 579. https://doi.org/10.3389/fpsyt.2019.00579

Hogarth, L., 2020. Addiction is driven by excessive goal-directed drug choice under negative affect: translational critique of habit and compulsion theory. Neuropsychopharmacol. Off. Publ. Am. Coll. Neuropsychopharmacol. 45, 720–735. https://doi.org/10.1038/s41386-020-0600-8

Hogarth, L., Field, M., 2020. Relative expected value of drugs versus competing rewards underpins vulnerability to and recovery from addiction. Behav. Brain Res. 112815. https://doi.org/10.1016/j.bbr.2020.112815

Hood, L.E., Leyrer-Jackson, J.M., Olive, M.F., 2020. Pharmacotherapeutic management of co-morbid alcohol and opioid use. Expert Opin. Pharmacother. 21, 823–839. https://doi.org/10.1080/14656566.2020.1732349

Jan, R.K., Lin, J.C., McLaren, D.G., Kirk, I.J., Kydd, R.R., Russell, B.R., 2014. The effects of methylphenidate on cognitive control in active methamphetamine dependence using functional magnetic resonance imaging. Front. Psychiatry 5, 20.

Jiang, Y., Oathes, D., Hush, J., Darnall, B., Charvat, M., Mackey, S., Etkin, A., 2016. Perturbed connectivity of the amygdala and its subregions with the central executive and default mode networks in chronic pain. Pain 157, 1970–1978. https://doi.org/10.1097/j.pain.0000000000000606

Jongenelis, M., Pettigrew, S., Lawrence, D., Rikkers, W., 2019. Factors Associated with Poly Drug Use in Adolescents. Prev. Sci. 20, 695–704. https://doi.org/10.1007/s11121-019-00993-8

Kandel, D., Kandel, E., 2015. The Gateway Hypothesis of substance abuse: developmental, biological and societal perspectives. Acta Paediatr. 104, 130–137. https://doi.org/10.1111/apa.12851

Kishore, K., 2014. Monograph of tobacco (Nicotiana tabacum). Indian J. Drugs 2, 5–23.

Klega, A.E., Keehbauch, J.T., 2018. Stimulant and Designer Drug Use: Primary Care Management. Am. Fam. Physician 98, 85–92.

Knackstedt, L.A., Ettenberg, A., 2005. Ethanol consumption reduces the adverse consequences of self-administered intravenous cocaine in rats. Psychopharmacology (Berl). 178, 143–150. https://doi.org/10.1007/s00213-004-1996-2

Koob, G.F., Le Moal, M., 2008. Addiction and the brain antireward system. Annu. Rev. Psychol. 59, 29–53. https://doi.org/10.1146/annurev.psych.59.103006.093548

Kringelbach, M.L., 2005. The human orbitofrontal cortex: linking reward to hedonic experience. Nat. Rev. Neurosci. 6, 691–702. https://doi.org/10.1038/nrn1747

Lacy, R.T., Strickland, J.C., Brophy, M.K., Witte, M.A., Smith, M.A., 2014. Exercise decreases speedball self-administration. Life Sci. 114, 86–92. https://doi.org/10.1016/j.lfs.2014.08.005

Laursen, H.R., Henningsson, S., Macoveanu, J., Jernigan, T.L., Siebner, H.R., Holst, K.K., Skimminge, A., Knudsen, G.M., Ramsoy, T.Z., Erritzoe, D., 2016. Serotonergic neurotransmission in emotional processing: New evidence from long-term recreational poly-drug ecstasy use. J. Psychopharmacol. 30, 1296–1304. https://doi.org/10.1177/0269881116662633

Leknes, S., Tracey, I., 2008. A common neurobiology for pain and pleasure. Nat. Rev. Neurosci. 9, 314–320. https://doi.org/10.1038/nrn2333

Leurquin-Sterk, G., Ceccarini, J., Crunelle, C.L., Weerasekera, A., de Laat, B., Himmelreich, U., Bormans, G., Van Laere, K., 2018. Cerebral dopaminergic and glutamatergic transmission relate to different subjective responses of acute alcohol intake: an in vivo multimodal imaging study. Addict. Biol. 23, 931–944. https://doi.org/10.1111/adb.12542

Loganathan, K., Ho, E.T.W., 2021. Value, drug addiction and the brain. Addict. Behav. 116, 106816. https://doi.org/10.1016/j.addbeh.2021.106816

Loganathan, K., Lv, J., Cropley, V., Ho, E.T.W., Zalesky, A., 2020. Associations between delay discounting and connectivity of the valuation-control system in healthy young adults. Neuroscience. https://doi.org/10.1016/j.neuroscience.2020.11.026

McKetin, R., Chalmers, J., Sunderland, M., Bright, D.A., 2014. Recreational drug use and binge drinking: stimulant but not cannabis intoxication is associated with excessive alcohol consumption. Drug Alcohol Rev. 33, 436–445. https://doi.org/10.1111/dar.12147

Mello, N.K., Mendelson, J.H., Sellers, M.L., Kuehnle, J.C., 1980. Effects of heroin self-administration on cigarette smoking. Psychopharmacology (Berl). 67, 45–52. https://doi.org/10.1007/BF00427594

Moss, H.B., Chen, C.M., Yi, H., 2014. Early adolescent patterns of alcohol, cigarettes, and marijuana polysubstance use and young adult substance use outcomes in a nationally representative sample. Drug Alcohol Depend. 136, 51–62. https://doi.org/10.1016/j.drugalcdep.2013.12.011

Müller-Oehring, E.M., Jung, Y.-C., Sullivan, E. V, Hawkes, W.C., Pfefferbaum, A., Schulte, T., 2013. Midbrain-Driven Emotion and Reward Processing in Alcoholism. Neuropsychopharmacology 38, 1844–1853. https://doi.org/10.1038/npp.2013.102

Napoli, J.L., Camalier, C.R., Brown, A.-L., Jacobs, J., Mishkin, M.M., Averbeck, B.B., 2021. Correlates of Auditory Decision-Making in Prefrontal, Auditory, and Basal Lateral Amygdala Cortical Areas. J. Neurosci. 41, 1301–1316. https://doi.org/10.1523/JNEUROSCI.2217-20.2020

Nuechterlein, E.B., Ni, L., Domino, E.F., Zubieta, J.-K., 2016. Nicotine-specific and non-specific effects of cigarette smoking on endogenous opioid mechanisms. Prog. Neuropsychopharmacol. Biol. Psychiatry 69, 69–77. https://doi.org/10.1016/j.pnpbp.2016.04.006

Oken, B.S., Salinsky, M.C., Elsas, S.M., 2006. Vigilance, alertness, or sustained attention: physiological basis and measurement. Clin. Neurophysiol. 117, 1885–1901. https://doi.org/10.1016/j.clinph.2006.01.017

Peele, S., 2016. People Control Their Addictions: No matter how much the “chronic” brain disease model of addiction indicates otherwise, we know that people can quit addictions - with special reference to harm reduction and mindfulness. Addict. Behav. reports 4, 97–101. https://doi.org/10.1016/j.abrep.2016.05.003

Plakke, B., Ng, C.-W., Poremba, A., 2013. Neural correlates of auditory recognition memory in primate lateral prefrontal cortex. Neuroscience 244, 62–76. https://doi.org/10.1016/j.neuroscience.2013.04.002

Power, J.D., Barnes, K.A., Snyder, A.Z., Schlaggar, B.L., Petersen, S.E., 2012. Spurious but systematic correlations in functional connectivity MRI networks arise from subject motion. Neuroimage 59, 2142–2154. https://doi.org/10.1016/j.neuroimage.2011.10.018

Ronen, A., Chassidim, H.S., Gershon, P., Parmet, Y., Rabinovich, A., Bar-Hamburger, R., Cassuto, Y., Shinar, D., 2010. The effect of alcohol, THC and their combination on perceived effects, willingness to drive and performance of driving and non-driving tasks. Accid. Anal. Prev. 42, 1855–1865. https://doi.org/10.1016/j.aap.2010.05.006

Saad, Z.S., Gotts, S.J., Murphy, K., Chen, G., Jo, H.J., Martin, A., Cox, R.W., 2012. Trouble at Rest: How Correlation Patterns and Group Differences Become Distorted After Global Signal Regression. Brain Connect. 2, 25–32. https://doi.org/10.1089/brain.2012.0080

Sarter, M., Givens, B., Bruno, J.P., 2001. The cognitive neuroscience of sustained attention: where top-down meets bottom-up. Brain Res. Brain Res. Rev. 35, 146–160. https://doi.org/10.1016/s0165-0173(01)00044-3

Scott, D.J., Domino, E.F., Heitzeg, M.M., Koeppe, R.A., Ni, L., Guthrie, S., Zubieta, J.-K., 2007. Smoking modulation of mu-opioid and dopamine D2 receptor-mediated neurotransmission in humans. Neuropsychopharmacology 32, 450–457. https://doi.org/10.1038/sj.npp.1301238

Shurman, J., Koob, G.F., Gutstein, H.B., 2010. Opioids, pain, the brain, and hyperkatifeia: a framework for the rational use of opioids for pain. Pain Med. 11, 1092–1098. https://doi.org/10.1111/j.1526-4637.2010.00881.x

Smith, S.M., Beckmann, C.F., Andersson, J., Auerbach, E.J., Bijsterbosch, J., Douaud, G., Duff, E., Feinberg, D.A., Griffanti, L., Harms, M.P., Kelly, M., Laumann, T., Miller, K.L., Moeller, S., Petersen, S., Power, J., Salimi-Khorshidi, G., Snyder, A.Z., Vu, A.T., Woolrich, M.W., Xu, J., Yacoub, E., Ugurbil, K., Van Essen, D.C., Glasser, M.F., 2013a. Resting-state fMRI in the Human Connectome Project. Neuroimage 80, 144– 168. https://doi.org/10.1016/j.neuroimage.2013.05.039

Smith, S.M., Jenkinson, M., Woolrich, M.W., Beckmann, C.F., Behrens, T.E.J., Johansen-Berg, H., Bannister, P.R., De Luca, M., Drobnjak, I., Flitney, D.E., Niazy, R.K., Saunders, J., Vickers, J., Zhang, Y., De Stefano, N., Brady, J.M., Matthews, P.M., 2004. Advances in functional and structural MR image analysis and implementation as FSL. Neuroimage 23 Suppl 1, S208–19. https://doi.org/10.1016/j.neuroimage.2004.07.051

Smith, S.M., Vidaurre, D., Beckmann, C.F., Glasser, M.F., Jenkinson, M., Miller, K.L., Nichols, T.E., Robinson, E.C., Salimi-Khorshidi, G., Woolrich, M.W., Barch, D.M., Ugurbil, K., Van Essen, D.C., 2013b. Functional connectomics from resting-state fMRI. Trends Cogn. Sci. 17, 666–682. https://doi.org/10.1016/j.tics.2013.09.016

Solinas, M., Belujon, P., Fernagut, P.O., Jaber, M., Thiriet, N., 2019. Dopamine and addiction: what have we learned from 40 years of research. J. Neural Transm. 126, 481– 516. https://doi.org/10.1007/s00702-018-1957-2

Stark, M.J., Campbell, B.K., 1993. Cigarette smoking and methadone dose levels. Am. J. Drug Alcohol Abuse 19, 209–217. https://doi.org/10.3109/00952999309002681

Su, S., Fairley, C.K., Mao, L., Medland, N.A., Jing, J., Cheng, F., Zhang, L., 2019. Estimates of the national trend of drugs use during 2000-2030 in China: A population-based mathematical model. Addict. Behav. 93, 65–71. https://doi.org/10.1016/j.addbeh.2019.01.022

van den Bos, W., McClure, S.M., 2013. Towards a general model of temporal discounting. J. Exp. Anal. Behav. 99, 58–73. https://doi.org/10.1002/jeab.6

Walker, R.B., 1980. Medical aspects of tobacco smoking and the anti-tobacco movement in Britain in the nineteenth century. Med. Hist. 24, 391–402.

Wang, L., Min, J.E., Krebs, E., Evans, E., Huang, D., Liu, L., Hser, Y.-I., Nosyk, B., 2017. Polydrug use and its association with drug treatment outcomes among primary heroin, methamphetamine, and cocaine users. Int. J. Drug Policy 49, 32–40.

Wang, T., Ma, J., Wang, R., Liu, Z., Shi, J., Lu, L., Bao, Y., 2018. Poly-Drug Use of Prescription Medicine among People with Opioid Use Disorder in China: A Systematic Review and Meta-Analysis. Subst. Use Misuse 53, 1117–1127. https://doi.org/10.1080/10826084.2017.1400066

Weigl, M., Anzenberger, J., Grabenhofer-Eggerth, A., Horvath, I., Schmutterer, I., Strizek, J., Tanios, A., 2017. Bericht zur Drogensituation 2017. Gesundheit Österreich.

Wilson, T.D., Barry, K.L., Maust, D.T., Blow, F.C., 2020. Association between relationship quality and concurrent alcohol use and sedative-tranquilizer misuse in middle and later life. Aging Ment. Health 1–5. https://doi.org/10.1080/13607863.2020.1727850

WU-Minn, H.C.P., 2017. 1200 subjects data release reference manual. URL https://www.humanconnectome.org.

Xie, C., Shao, Y., Ma, L., Zhai, T., Ye, E., Fu, L., Bi, G., Chen, G., Cohen, A., Li, W., Chen, G., Yang, Z., Li, S.-J., 2014. Imbalanced functional link between valuation networks in abstinent heroin-dependent subjects. Mol. Psychiatry. https://doi.org/10.1038/mp.2012.169

Zhai, T., Shao, Y., Chen, Gang, Ye, E., Ma, L., Wang, L., Lei, Y., Chen, Guangyu, Li, W., Zou, F., Jin, X., Li, S.-J., Yang, Z., 2015. Nature of functional links in valuation networks differentiates impulsive behaviors between abstinent heroin-dependent subjects and nondrug-using subjects. Neuroimage 115, 76–84. https://doi.org/10.1016/j.neuroimage.2015.04.060

Zou, F., Wu, X., Zhai, T., Lei, Y., Shao, Y., Jin, X., Tan, S., Wu, B., Wang, L., Yang, Z., 2015. Abnormal resting-state functional connectivity of the nucleus accumbens in multi-year abstinent heroin addicts. J. Neurosci. Res. 93, 1693–1702. https://doi.org/10.1002/jnr.23608

